# The Impact of Non-Pharmaceutical Interventions on the First COVID-19 Epidemic Wave in South Africa

**DOI:** 10.1101/2021.06.29.21259625

**Authors:** Thabo Mabuka, Nesisa Ncube, Michael Ross, Andrea Silaji, Willie Macharia, Tinashe Ndemera, Tlaleng Lemeke

## Abstract

On the 5^th^ of March 2020, South Africa reported its first cases of COVID-19. This signalled the onset of the first COVID-19 epidemic wave in South Africa. The response by the Government of South Africa to the COVID-19 epidemic in South Africa was the use of non-pharmaceutical interventions (NPIs). In this study, a semi-reactive COVID-19 model, the ARI COVID-19 SEIR model, was used to investigate the impact of NPIs in South Africa to understand their effectiveness in the reduction of COVID-19 transmission in the South African population. This study also investigated the COVID-19 testing, reporting, hospitalised cases and excess deaths in the first COVID-19 epidemic wave in South Africa. The results from this study show that the COVID-19 NPI policies implemented by the Government of South Africa played a significant role in the reduction of COVID-19 active, hospitalised cases and deaths in South Africa’s first COVID-19 epidemic wave.

## 1. Introduction

The study of the occurrence of a disease is called epidemiology [1]. A disease is called a pandemic when there is a rapid increase of cases of the disease in a relatively short time and, a disease is endemic if it is within the population for a relatively long time. Diseases can be caused by various agents such as bacteria and viruses and transmitted by various modes, such as human to human contact, reservoir to vector to human such as in malaria. Every disease has a specific agent and mode of transmission [1]. COVID-19 is the disease caused by the SARS-CoV-2 (Severe acute respiratory syndrome coronavirus 2). The ‘CO’ stands for corona, ‘VI’ for virus, and ‘D’ for disease. Formerly, this disease was referred to as the ‘2019 novel coronavirus’ or ‘2019-nCoV.’ [2,3]. Coronaviruses are a large family of viruses that are attributed to causing mild respiratory infections such as the common cold to more severe diseases such as the Middle East Respiratory Syndrome (MERS) and Severe Acute Respiratory Syndrome (SARS) [4]. The SARS-CoV-2 virus is a zoonotic virus belonging to the Betacoronavirus 2B lineage, similar to Coronaviruses found in bats and pangolins. Bats and pangolins are common reservoirs for Coronaviruses however, SARS-CoV-2 has not been isolated from bats or pangolins and there is a suspected inconclusive intermediate host in the initial transmission of the SARS-CoV-2 to humans [4,5]. In December 2019 and January 2020, there was a cluster of cases of respiratory illnesses in the province of Wuhan City, Hubei Province, China. This cluster of cases was later determined to be caused by the SARS-CoV-2. The SARS-CoV-2 was identified and isolated on the 7^th^ of January in China [6]. On January 30, 2020, the WHO declared the COVID-19 outbreak a global health emergency and on March 11, 2020, a global pandemic [7].

The SARS-COV-2’s main mechanism for host entry is through interactions with the host angiotensin-converting enzyme II (ACE2) located on the host cell’s surface. In humans, these enzymes are most abundant in the epithelia of the lungs and small intestines [8]. Hence an infection with SARS-COV-2 is characterised by respiratory illness/disease. The immune response to SARS-COV-2 in humans is thought to be both innate and adaptive. The innate response attributing to the early symptoms of COVID-19 which are fevers and muscle aches [9]. SARS-COV-2 infections cause various symptoms such as coughing/sore throat, fever, myalgia or fatigue, respiratory symptoms, pneumonia. In moderate disease, pneumonia is reported and becomes severe in severe cases. In critical cases, it can cause acute respiratory distress syndrome (ARDS), dyspnoea, respiratory failure, sepsis, septic shock, acute thrombosis and multiple organ failure [10–12]. It has been hypothesised that there might be an over-response by the immune system (Cytokine storm syndromes). The result is the generation of fluids and inflammation, damage to respiratory cells especially in severe and critical cases [13]. Severe and critical COVID-19 cases need assisted/mechanical breathing. Most COVID-19 patients who recovered developed antibodies to the SARS-CoV-2 virus within 1 to 3 weeks [9]. Even though recovered cases show the presence of late antibodies (IgG), there is still uncertainty in their titre levels and neutralisation for an effective immune response to a secondary SARS-CoV-2 infection. Some studies have shown the disappearance of neutralising antibodies reacting to SARS-COV-2 after 3 months [14,15]. However, the adaptive immune response’s ability to produce memory cells (remaining T-cells and B-cells after primary infection) is unknown for COVID-19 [9]. SARS-CoV-2 is an RNA virus and RNA genetic material is prone to mutations however coronaviruses have the capacity for proofreading during replication which results in relatively lower rates of mutation [16]. Regardless of the capacity to proofread, there have been several reported variants of SARS-CoV-2 that have been identified since the outbreak. Some variants have raised concern about their impacts on transmissibility, clinical characteristics particularly in COVID-19 disease severity, diagnostics, therapeutics, and vaccines. Currently of note, are the SARS-CoV-2 variants B.1.1.7 (SARS-CoV-2 VOC 202012/01 first identified in the UK), B.1.351 (501Y.V2 first identified in South Africa) and P.1 (first identified in Brazil) [17]. There have been reports of increased transmissibility and reduced vaccine efficacy with some of the mentioned variants [18].

On the 5^th^ of March 2020, South Africa reported its first cases of COVID-19 [19]. This signalled the onset of the first wave of the COVID-19 epidemic in South Africa where the numbers of the reported cases started to increase exponentially. According to the data provided by the National Institute of Communicable Diseases (NICD), the first wave of the pandemic in South Africa lasted from 05 March 2020 to 1 October 2020. From 05 March to mid-June, the positivity rate stood at 0.02 cases per 100 000 people in the population per week and increased rapidly between mid-June and mid-July signalling the peak of the wave where the positivity rate had risen to 138.1 per 100 000 people in the population. During the first epidemic wave period, 676 084 confirmed COVID-19 cases had been reported and 16 866 reported deaths. More than 4.2 million tests had been conducted. Western Cape, KwaZulu-Natal and Gauteng provinces had the highest number of cases with the confirmed cases in these provinces accounting for more than 66.5 % of the total reported cases in the country. From the reported cases, 609 854 had been reported to have recovered from COVID-19 which translated to a recovery rate of 90.2 % [20].

The response of governments in the wake of the COVID-19 pandemic was the use of non-pharmaceutical interventions (NPIs) [21]. Non-Pharmaceutical Interventions (NPIs) are actions taken by a population to slow down the transmission of disease, apart from vaccination and medicinal treatment [22]. In an African context, a region with a relatively high disease burden, NPIs have played a significant role in controlling disease in the population. For example, in the case of the Cholera outbreak in Africa, citizens were informed to boil all naturally sourced water before drinking, constant washing of hands and the proper disposal of human faecal matter. Sanitation played a significant role in containing the Cholera outbreak in Africa [23]. In the case of the HIV epidemic, a disease with a relatively high prevalence in the Southern African region, HIV epidemic needle and syringe programmes to prevent the sharing of needles were implemented in conjunction with increased condom distribution and use (Avert.org, 2020). The Ebola outbreak in 2014 which was mostly reported in the Western African region, also saw the implementation of widespread encouragement of frequent hand washing, the avoidance of contact with infected individuals, screening and testing at borders, in addition to the wearing of full-body Personal Protective Equipment (PPE) when dealing with infected patients as well as with the deceased who had succumb to Ebola (Mayo Clinic, 2020). On a global scale, the use of NPIs can be referenced in the Influenza (H1N1) Pandemic of 1918. During this pandemic, worldwide, populations were confined to their residences, public gatherings were banned, schools and public institutions were closed, infected individuals were quarantined and widespread mask wearing became the norm within a distanced public (Billing, 2005). Significant parallels can be drawn between the implementation of NPIs in the COVID-19 pandemic and the Influenza Pandemic of 1918 due to the similar modes of transmission of the two diseases. Similar to the Influenza pandemic, in the early stages of the COVID-19 pandemic conclusions were reached that the “prevention of contact” was paramount in the management the outbreak.

Of interest in this study is the response to the COVID-19 epidemic in South Africa. In South Africa, the Government of South Africa declared a state of disaster and created a National Coronavirus Command Council to oversee the COVID-19 outbreak [24]. There were several National Lockdown Alert Levels declared by the council to try to curb the rate of infection to avoid colossally overwhelming the South African health care system. These policies focused on limiting contact in the population through movement restrictions, curfews, limiting services, restriction of business and trade, isolation and quarantine of infected persons and use of PPE. The understanding of the COVID-19 policy response in South Africa and its impact can aid in the development of NPI policies in future epidemics, particularly in an African context. Epidemiological modelling can be a powerful tool to assist in understanding the scale of transmission, disease severity, and the effectiveness of NPIs for policy development, disease control and prevention. Epidemiological models provide the ability to predict the macroscopic behaviour of diseases using microscopic descriptions. One of the many ways used in modelling an epidemic is through deterministic (based on average characteristics of the population characteristics under study) and/or stochastic modelling (based on the randomness of the elements of the population) [25]. Stochastic modelling appears to be more accurate in evaluating real-life epidemic propagation, hence the most used and preferred [25]. Stochastic models can be classified into 3 main groups: The SI, SIS and SIR model [1]. These compartmentalise a population into classes as a function of time with the rates determined by the clinical and social characteristics of the disease in the population: The Susceptible class-(S) (these are individuals who have no effective immunity and have not been infected yet); Exposed class (E) (Individuals who have contracted the virus but are still not yet infectious); Infective class (I) (these are individuals who are infected and are infectious that is transmitting the disease to others) and Removed/Recovered (R) (these are individuals who have recovered from the disease with immunity, isolated or died). In the SI model infected individuals do not recover whilst in the SIS model individuals recover with no immunity and in the SIR model individuals recover with immunity [1].

COVID-19 has been widely modelled with variations of the SEIR model [26–32]. One of the earliest Global COVID-19 transmission models to be published was the Imperial College London COVID-19 Model [28,33]. The Imperial College London COVID-19 Model had a great influence in the early policy response to COVID-19 in the United Kingdom and many other countries including Africa [28]. Another COVID-19 Model of note was the model produced by the Center for Disease Dynamics, Economics & Policy (CDDEP) [27,29]. The CDDEP COVID-19 Models tried to understand the impact of Country-Wise Lockdowns (Frost, Craig, et al., 2020) and Health Care system preparedness in African countries [27]. South Africa received much attention with regards to COVID-19 Modelling with several models being published and noted by the Government of South Africa [34]. Of note, is the National COVID-19 Epi Model (NCEM) and the National COVID-19 Cost Model (NCCM) by the South African COVID-19 Modelling Consortium, 2020. The NCEM is an SEIR stochastic compartmental transmission model that was developed to estimate the total and reported incidence of COVID-19 cases in South Africa up to November 2020. While the NCCM was a model developed to determine the COVID-19 response budget in South Africa. The NCEM and NCCM played a key role in South Africa’s early policy and planning response to COVID-19. While most of the mentioned COVID-19 models have been pro-active, there is a need for semi-reactive models to help assess the post-COVID-19 epidemic with parameters derived from real reported case data. This allows for improvement in the accuracy of modelling parameters and outputs.

In this study, a semi-reactive COVID-19 model, the ARI COVID-19 SEIR model, was used to investigate the impact of NPIs in South Africa to understand their effectiveness in the reduction of transmission of COVID-19 in the South African population. This study also investigated the COVID-19 testing, reporting, hospitalised cases and excess deaths in the first wave of the COVID-19 epidemic in South Africa. The understanding of the NPIs developed in this study is aimed at assisting with early NPI policy development in South Africa and Africa for current and future epidemics.

## 2. Methodology

### 2.1 Model Structure

To model the transmission dynamics of COVID-19 in the first COVID-19 epidemic wave in South Africa a stochastic compartmental transmission SEIR model was used hereafter called the “ARI COVID-19 SEIR” model. Figure 1 shows the structure of the ARI COVID-19 SEIR model. The ARI COVID-19 SEIR Model was constructed in a Macro-Enabled Microsoft (Ms) Excel File for user-friendliness (visual interaction with parameters) and Database Query Support. The Model had a Visual Basic Application (VBA) code for Sensitivity and Variable Analysis.

**Figure 1:**
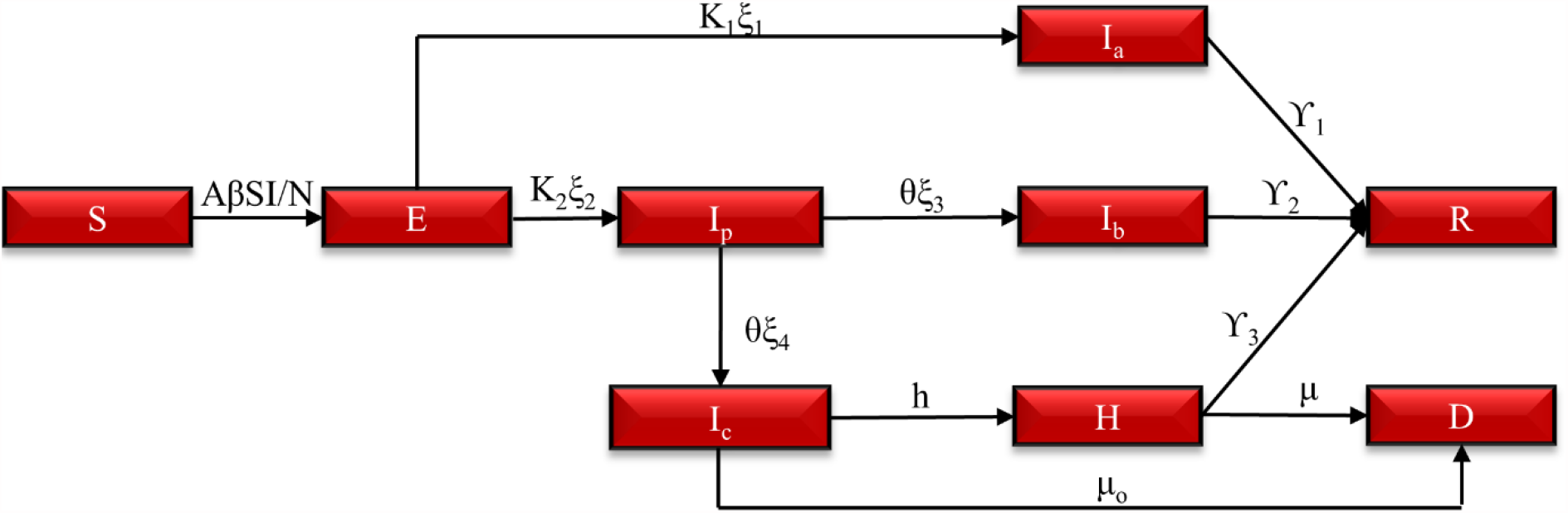
The ARI COVID-19 SEIR Model structure

Figure 1 shows that the ARI COVID-19 SEIR model had the following population classes based on the assumed clinical diagnosis of COVID-19 within the population:

#### Susceptible (S)

Individuals within the population of the model who can incur the disease however have not been infected yet.

#### Exposed (E)

Individuals within the population of the model in an incubation period who are not yet infectious.

#### Asymptomatic (I_a_)

Individuals within the population of the model who are infected and are infectious that is transmitting the disease to others however are not showing any symptoms throughout their infectiousness.

#### Pre-symptomatic Infectious (I_p_)

Individuals within the population of the model that are transmitting the disease during their incubation period.

#### Infected with Mild and Moderate Symptoms (I_b_)

Individuals within the population of the model with mild and moderate symptoms who are infectious.

#### Infected with Severe and Critical Symptoms (I_c_)

Individuals within the population of the model with severe and critical symptoms who are infectious however have not yet been hospitalised.

#### Hospitalised COVID-19 Cases (H)

Individuals within the population of the model with severe and critical symptoms who have been hospitalised.

#### Death due to COVID-19 (D)

Individuals who have died due to COVID-19 or indirect consequences of the COVID-19 epidemic.

#### Recovered (R)

Individuals within the population of the model who have recovered from the disease with immunity or partial immunity.

### 2.2 Model Equations

The transmission and severity of COVID-19 within-population classes in the model were simulated using ordinary differential equations (ODEs). The differentiation in the ODEs was conducted using Euler’s method with 1-day estimation steps. The total population (N) in the model is represented by Equation 1 based on the conservation of mass. Vitals (new births and non-COVID-19 deaths) were not considered in the model due to the relatively small annual growth rate of 1.40 % in the South African population [36] and the low incident of COVID-19 in neonatal [37]. The ARI COVID-19 SEIR model instead used the 2020 South African population estimates from the United Nations (UN) World Population Prospects [38]. The total infections are given by Equation 2.

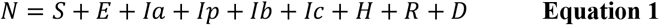

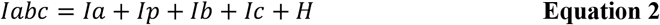

The change in the susceptible population class is given by Equation 3 where β is the Effective daily contact rate, this is the average number of adequate contacts per infective per day. The product of S and I in Equation 3 is referred to as the mass incident term.

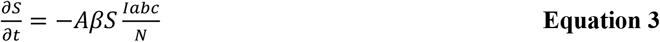

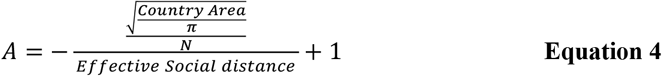

A is the Population density factor given by Equation 4. Where the Effective Social distance is the minimum distance between infector and infectee which prevents infection. For COVID-19 a distance of 2 m was assigned [39]. As the average distance between individual tends towards the effective social distance, the Population Density Factor (A) tends towards 0. The Population Density Factor assumes a uniform distribution of population within a confined area. The change in the exposed class is given by Equation 5.

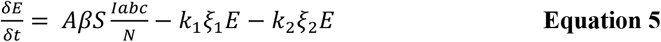

Where K_1_ and K_2_ are the rates in which an exposed individual move to the infected class. K_1_ is inversely proportional to the average incubation period of COVID-19 Asymptomatic Cases (T_inc,1_) and K_2_ is inversely proportional to the average incubation period of COVID-19 Symptomatic Cases (T_inc,2_) in the population. ξ_1_, ξ_2_, ξ_3,_ ξ_4_ are the proportions of the exposed and pre-symptomatic who will be Asymptomatic (ξ_1_), Symptomatic (ξ_2_), Develop Mild and Moderate Symptoms (ξ_3_) and Severe and Critical Symptoms (ξ_4_), respectively.

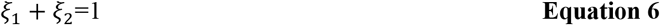

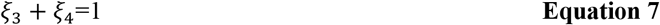

The change in the infectious class is given by Equation 8-12.

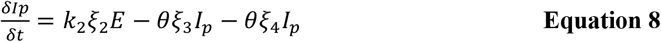

Equation 8 reduces to Equation 9 by substituting Equation 7.

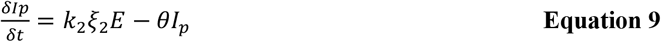

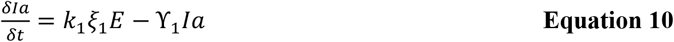

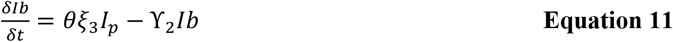

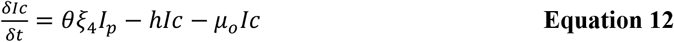

The change in the Hospitalised, Death and Recovered class is given by Equation 13, 14, 15, respectively.

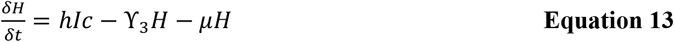

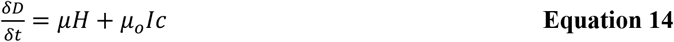

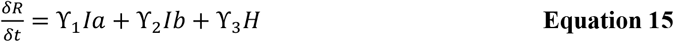

Where, ϒ_1_, ϒ_2_ and ϒ_3_ are the daily recovery rates of individuals with Asymptomatic, Mild and Moderate Symptoms and Severe and Critical Symptoms, respectively. θ is the rate at which pre-symptomatic individuals develop symptoms. h is the rate at which individuals who have developed severe and critical cases are hospitalised. μ_o_ is the daily death rate due to direct and indirect effects of COVID-19 in individuals with severe and critical symptoms who have not been hospitalised. *μ* is the daily death rate due to COVID-19 in hospitalised individuals.

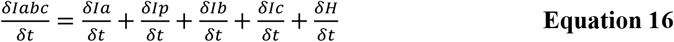

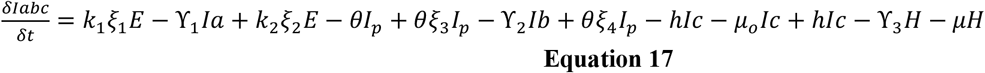

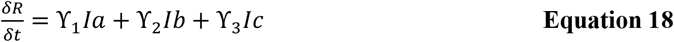

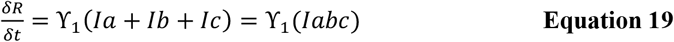

### 2.3 Model Basic Reproductive Number and Herd Immunity

For the feasible region in the octant of the mathematic model, there exists an equilibrium in which there is no disease called the Disease-Free Equilibrium (DFE). This condition is satisfied by the stability of the rate of change of the population. Thus:

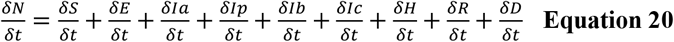

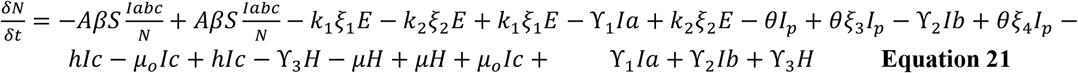

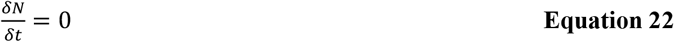

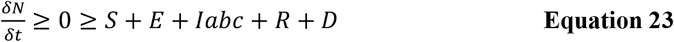

At Disease Free Equilibrium (DFE), E=0, I=0, R=0, D=0. Therefore, substituting DFE values into Equation 23 gives Equation 24.

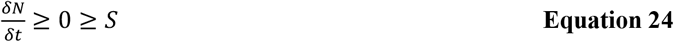

At each point in time in the model, there also exists an equilibrium in which there is a maximum/minimum for each class. This equilibrium is called the Endemic Equilibrium (EE). Thus, taking Equation 3 and Equation 9-15 where the rate of change is 0 gives Equation 25-33

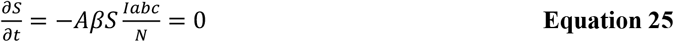

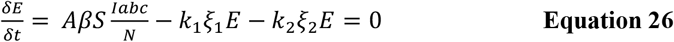

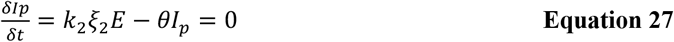

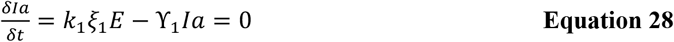

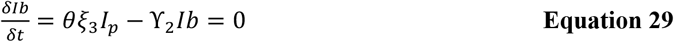

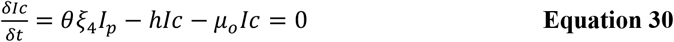

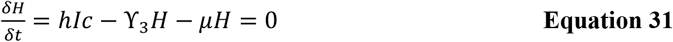

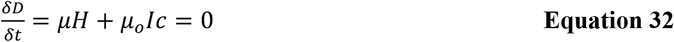

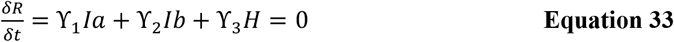

Substituting Equation 27-31 into Equation 2 gives Equation 34.

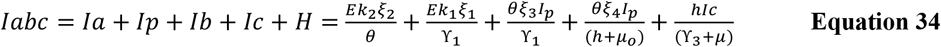

Substituting Equation 27 and 30 into Equation 34 gives Equation 35.

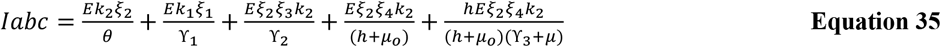

From Equation 26, making S the subject of the equation gives Equation 36:

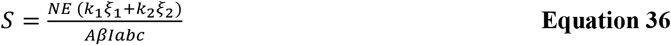

Substituting Equation 35 into Equation 36 gives Equation 37.

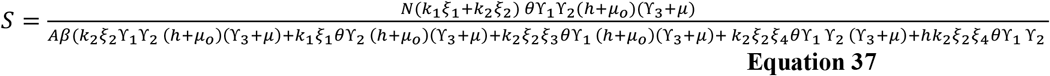

The relative critical point for the model is when DFE=EE. Using Equation 24 and Equation 36 we derive Equation 38-40.

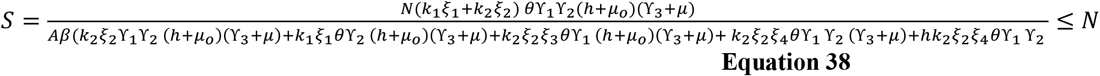

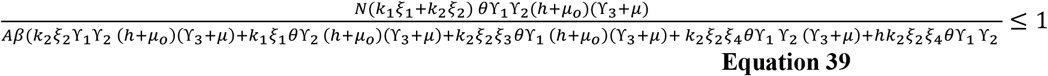

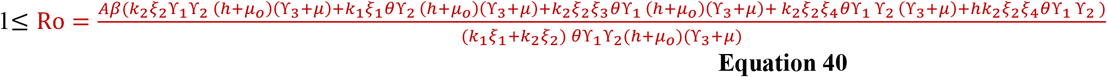

Equation 40 is what is defined as the basic reproductive number (Ro) for the ARI COVID-19 SEIR model. The basic reproductive number is the number of secondary infections that one infected person would produce in a fully susceptible population through the entire duration of the infectious period. Ro provides a threshold condition for the stability of the disease-free equilibrium point [1]. If Ro is greater than 1 then there is an endemic equilibrium thus there will be an epidemic. If Ro is less than 1 then the disease will die out and remain at a relatively low level to the population size. As can be seen from Equation 40, Ro can be summarised as a ratio of the daily contact number over the daily recovery rate. It can also be defined by Equation 41:

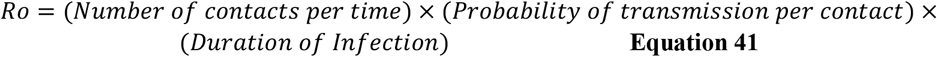

The basic reproductive number assumes a completely susceptible population however the infection productiveness of the population changes as infections increase. The effective reproductive number, Re, is the number of secondary infections that one infected person would produce through the entire duration of the infectious period. It can be estimated based on the susceptible class given by Equation 42:

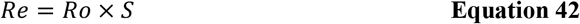

Herd immunity is an important concept in epidemiology. Herd immunity is when the population has enough people immune such that the disease will not spread if it was suddenly introduced randomly in the population. Consider if α is the fraction immune due to vaccination/acquire immunity. Then Heard Immunity is given by **Equation 43:**

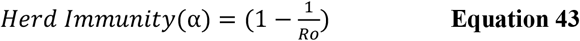

### 2.4 Model Parameters

#### 2.4.1 Determining the Hospital Discharge Rate in South Africa

The average Daily Hospital Discharge Rate (ϒ_3_) in South Africa was calculated based on clinical information from admitted patients with laboratory-confirmed COVID-19 in selected hospitals in South Africa under the National Institute for Communicable Diseases (NICD) DATCOV surveillance system. The NICD sentinel hospital surveillance system was designed to monitor and describe trends of COVID-19 hospitalizations and the epidemiology of hospitalized patients in South Africa [40]. The number of hospitals reporting in the NICD DATCOV surveillance system increased in the reporting period. Initially, 204 Facilities were reporting, and this increased to 434 Facilities by 4 September 2020 [41]. Therefore, caution was taken when taking averages between reporting case dates. The average Daily Hospital Discharge Rate (ϒ_3_) for COVID-19 patients was calculated using the Number of Discharged Alive and Admitted patients Data in the NICD DATCOV surveillance system from 24 May to 01 October 2020. Equation 44 was used to calculate the Daily Discharge:

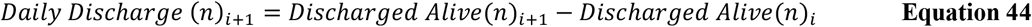

Where n is the number of patients and i is the reported case date.

The average Daily Hospital Discharge Rate (ϒ_3_) was then calculated using Equation 45.

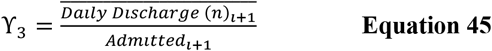

#### 2.4.2 Determining the Death Rate in Hospitalised Cases in South Africa

The average Daily Death Rate or Daily Case Fatality Rate (CFR) for COVID-19 patients (μ_1_) was calculated using the Number of Daily Deaths and Admitted patients Data in the NICD DATCOV surveillance system from 24 May-01 October 2020. The WHO guideline in estimating the CFR in [42] was followed. Equation 46 was used to calculate the Daily Deaths:

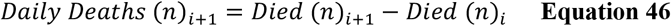

Where n is the number of patients and i is the reported case date.

The Daily Case Fatality Rate (CFR) for COVID-19 patients (μ_1_) was then calculated using Equation 47.

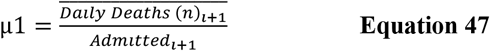

#### 2.4.3 Determining the Death Rate of Unreported Severe and Critical Cases in South Africa

Excess mortality is a count of deaths from all causes relative to what would normally have been expected. Excess mortality/deaths allow for accounting for miscounted or underreported COVID-19 Deaths and indirect Deaths related to the COVID-19 pandemic. National statistical agencies publish weekly deaths and averages of past ‘normal’ deaths [43]. In South Africa, the South African Medical Research Council (SAMRC) published the Excess deaths from 29 December 2019 to 01 October 2020 using information obtained from the National Population Register [44,45]. The Unreported Excess Deaths (Natural) to COVID-19 Death Ratio was calculated for the period, 25 March to 01 October 2020 with data from Bradshaw et al. (2020) using Equation 48:

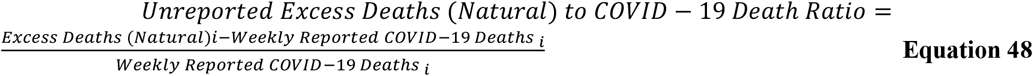

Where i is the Weekly Reported Date. The daily death rate due to direct and indirect effects of COVID-19 in individuals with severe and critical symptoms who have not been hospitalised (μ_o_) was then calculated using Equation 49:

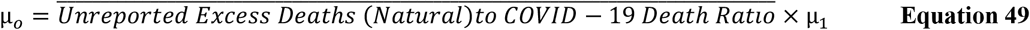

#### 2.4.4 Determining the COVID-19 Patient Admission Status in South Africa

The average admission status for COVID-19 patients was calculated using the Number of patients Currently in Hospital (n), General Ward (n), High Care (n), Intensive Care Unit (n), Isolation Ward (n), On Oxygen (n) and On Ventilator (n) Data in the NICD DATCOV surveillance system from 24 May-1 October 2020. The average admission Status was calculated using Equation 50-55:

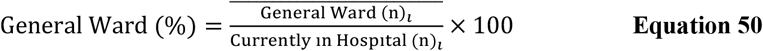

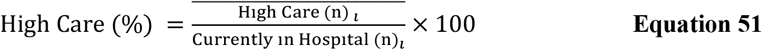

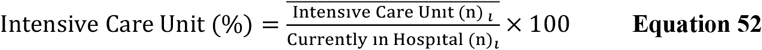

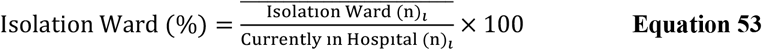

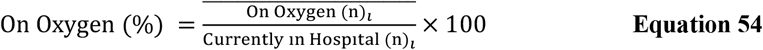

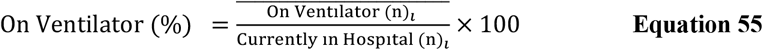

The admission status was then calculated using the Hospitalised Cases (H) in the model with Equation 56:

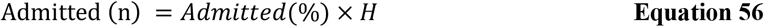

Where the Admitted (%) is the General Ward (%), High Care (%), Intensive Care Unit (%), Isolation Ward (%), On Oxygen (%) and Ventilator (%) respectively. A summary of the model parameters used in the ARI-COVID-19 Model are given in Table 1:

**Table 1:** ARI COVID-19 SEIR model parameters for the South African first COVID-19 epidemic wave.

| Model Parameters | Value Used | Source |
| --- | --- | --- |
| $\beta$ | 0.14-0.50 day<sup>-1</sup> | Model defined using Statical Regression Analysis |
| $\xi_1$ | 0.75 | 40-80 % [47-49] |
| $\xi_3$ | 0.98 | 80-99 % [5,50,51] |
| $T_{inc,1}$ | 1 Day | 1-4 days [5,52,53] |
| $T_{inc,2}$ | 1 Day | 1-4 days [5,26,52,53] |
| $T_p$ | 3 Days | 1-4 days [5,52,53] |
| $T_{inf,1}$ | 11 Days | 6.5-9.5 days [52] |
| $T_{inf,2}$ | 2 Days | 2-5 days [26] |
| $T_{testing}$ | 10 Days | Model defined from data from [41] |
| $T_{disch}$ | 12 Days | Model defined from data from [41] |
| $T_h$ | 5 Days | [31] |
| $\mu_1$ | 0.0195 day<sup>-1</sup> | Model defined from data from [41] |
| $\mu_o$ | 0.0315 day<sup>-1</sup> | Model defined from data from [41] and [44,45] |
| N | 59 308 690 people | [38] |
| A | 0.995 | Model defined |
| Ro | 1.37-4.73 | Model defined |
| $\alpha$ | 27-79 % | Model defined |

### 2.5 Model NPI Scenarios and Seeding

#### 2.5.1 Modelling Periods and Reported Case Data

To model the impact of NP1s in South Africa, the National Lockdown Alert Level 5, 4, 3 policies implemented by the South African government were modelled as scenarios in the ARI COVID-19 SEIR model, respectively. A “No lockdown” scenario where there was no policy response from the South African government was also modelled in the ARI COVID-19 SEIR model.

In each scenario, the ARI COVID-19 SEIR model was seeded using COVID-19 deaths with the Unreported Excess Deaths (Natural) to COVID-19 Death Ratio used to account for excess deaths in the National Lockdown Alert Level 3 scenario. For the “No lockdown” Scenario, reported active cases were used to seed the model due to no reported COVID-19 and Excess (Natural) Deaths in this period. Table 2 shows the model classification of the South African COVID-19 policy and the seeding period used in the model.

**Table 2:** South Africa COVID-19 Policy response and Period Implemented, Model Classification and Model Seeding Period

| COVID-19 Policy Response | Model Classification | Model Seeding Period |
| --- | --- | --- |
| No Lockdown | No lockdown | 2020/03/14-2020/03/27 |
| National Lock down Alert Level 5 | Hard-Lockdown | 2020/04/14-2020/04/30 |
| National Lock down Alert Level 4 | Moderate-Lockdown | 2020/05/01-2020/05/31 |
| National Lock down Alert Level 3 | Soft-Lockdown | 2020/06/01-2020/08/17 |

Cumulative Daily Reported COVID-19 Case, Recovery and Death Data for South Africa were obtained from the Johns Hopkins University (JHU) Center for Systems Science and Engineering (CSSE) COVID-19 Database [54] for the period of 22 January 2020 to 1 October 2020. Since reported case data come after a period of clinical diagnosis, reported case dates thus are lagged from the “real-time” date of infection. Therefore, an Average Date of infection date was estimated using Equation 57:

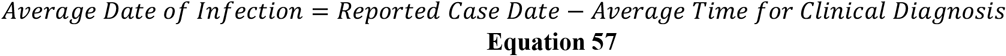

Where the Average time for Clinical diagnosis (T_testing_) is the average time taken for an infected person to be diagnosed and the diagnosis outcome to be classified and reported as a COVID-19 case. The COVID-19 Active Cases were determined using Equation 58:

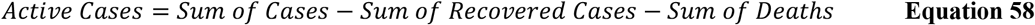

#### 2.5.1 Regression and Statistical Analysis

To seed the model, a non-linear regression analysis was conducted between the Reported COVID-19 Deaths and Death due to COVID-19 (D) from the model for the model seeding periods stated in Table 2. Seeding the Models with points that are oversensitive results in large deviations between modelled data versus reported case data. This deviation or noise introduced by “over-sensitive” data points creates a significant error in the model results. Thus, it was important to decide which Data points can be used in the regression analysis. For the modelling of NPIs, this was particularly important at the start of the pandemic where data values are relatively small. To decide seeding data points, a Data Point Sensitivity term described by Equation 59 was used:

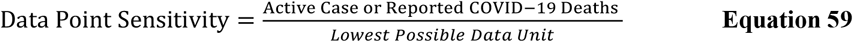

Where the Lowest Possible Data unit is the lowest possible unit of measurement for that data. In this case, it is 1 COVID-19 Reported Case. Data points with a Data Point Sensitivity greater than 5 % were ignored in the regression analysis. To conduct the regression analysis, the Residual and Normalised Errors were determined using Equation 60 and Equation 61:

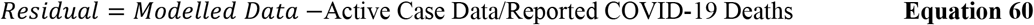

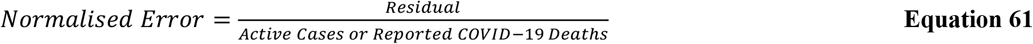

To allow the goodness of fit of Modelled data to Active Case or Reported COVID-19 Deaths Data, the Average Normalised Error of all Data points used in the Regression Analysis was reduced to 0 by changing the Effective Daily Contact Number (β) using the What-If Analysis Function in MS Excel. The pooled sample variance (s^2^) was then calculated using Equation 62:

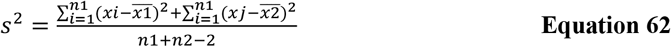

Where xi is the Reported Case Data points, 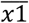 is the Reported Case Data Points Mean, xj is the Model Data points, 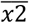 is the Model Data Points Mean, n1 and n2 are the samples sizes. The T-value: Two-Sample Assuming Equal Variance was used to calculate the t-value using Equation 63

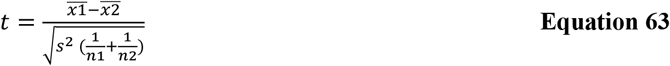

For the No lockdown scenario, since reported active cases were used to seed this model scenario, a sensitivity analysis was run to determine the fraction of reported cases that are pre-symptomatic, mild & moderate, asymptomatic, and critical & severe. This was done using a VBA code following the computational steps outlined in Figure 2.

**Figure 2:**
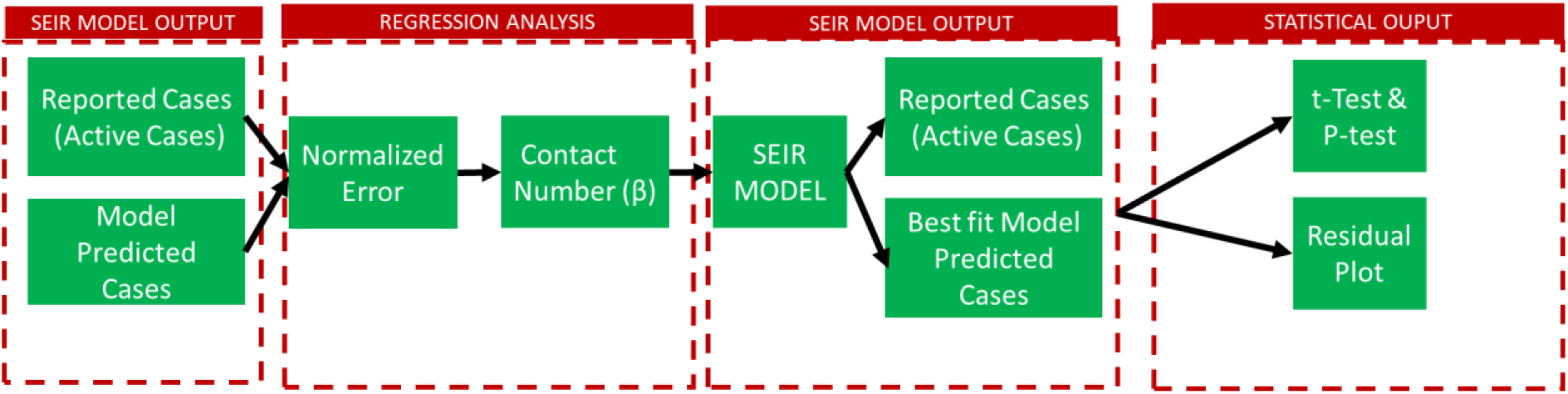
ARI COVID-19 SEIR Model Scenario Statistical Analysis Computational Steps

The combination of the Φ_1_ Φ_2_ Φ_3_ Φ_4_ that resulted in the lowest T-value (most significance) between model data and reported case data for the No Lockdown Scenario are shown in Table 3 and were chosen to seed the No Lockdown Scenario.

**Table 3:** Fraction of Pre-symptomatic, Mild & Moderate, Asymptomatic and Critical & Severe in Reported Cases

|  |  |
| --- | --- |
| $\Phi_1$ (Fraction of Mild & Moderate Cases Reported) | 0.5 |
| $\Phi_2$ (Fraction of Pre-symptomatic Cases Reported) | 1 |
| $\Phi_3$ (Fraction of Asymptomatic Cases Reported) | 0.1 |
| $\Phi_4$ (Fraction of Critical & Severe Cases Reported) | 1 |

For other Model Scenarios (South Africa National Lockdown Alert Level 5, 4 and 3), Reported COVID-19 Deaths were used to seed the Models. However, for the National Lockdown Alert Level 3 the reported COVID-19 deaths were adjusted to include unreported COVID-19 Deaths using Equation 64:

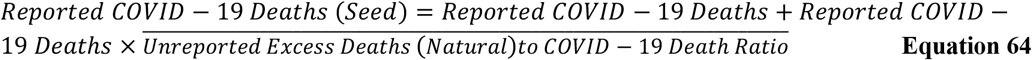

### 2.6 Validation of Model

The functionality and data produced by the ARI COVID-19 SEIR model were validated using the following:

- For model functionality, the model ODEs were validated by setting the model scenario reproductive number (Ro) to 1 by changing the Effective Daily Contact Number (β) using the What-If Analysis Function in MS Excel. At Ro=1, there should be no transmission of the population between model classes and model class values should be either at initial values or 0.
- For model data, comparison between COVID-19 Admitted data in NICD DATCOV surveillance system and Hospitalised COVID-19 Cases (H) in the model. Comparison between Excess (Natural) deaths data and Death due to COVID-19 (D) in the model. Comparison between the reported date of peak Active cases and Total infections (Iabc) in the model. Comparison between the ARI COVID-19 SEIR Model results and results from the South Africa National COVID-19 Epi Model (NCEM), the Center for Disease Dynamics, Economics & Policy (CDDEP) COVID-19 Model and the Imperial College London COVID-19 Model.

## 3. Results and Discussions

### 3.1 Impact of South African COVID-19 Policy on Movement, Contact Rate and Reproductive Number

Table 4 shows a summary of the National Lockdown Alert Level policies implemented by the South African government in the duration of the first COVID-19 epidemic wave in South Africa. The National Lockdown Alert Levels were implemented under South Africa’s Disaster Management Act, 2002 (Act NO. 57 of 2002) [55]. The first COVID-19 case in South Africa was reported on the 5^th^ of March 2020 [19]. For the first 20 days after the first reported case in South Africa, there was no NPI COVID-19 policy implemented however the country geared towards policy implementation by declaring a state of emergency and establishing a National Coronavirus Command Council to oversee the COVID-19 pandemic in South Africa. The first stringent measure initiated by the Council was a National Lockdown Level Alert 5 to try to curb the rate of infection to avoid colossally overwhelming the South African health care system. The National Lockdown Level Alert 5 was declared on the 26th of March 2020 to the 15th of April 2020 and then extended to the 30th of April 2020. The South African National Lockdown Level Alert 5 as shown in Table 4 was predominantly movement restrictions and limitation of services to essential services. Under this level the South African borders and air space were closed, there was an enforcement of strict non-movement of non-essential personal and a ban on some of the industries such as the alcohol and tobacco industry. To enforce this the South African Defence Force was deployed to oversee the compliance of this measure (Government of South Africa, 2020b). A screening and testing program for COVID-19 was initiated under the South African Department of Health. The initial testing was conducted by the National Institute of Communicable Disease (NICD) and this was expanded to a larger network of private and National Health Laboratory Services (NHLS) (NICD, 2020b). Mobile testing units were also deployed particularly to the hardest-hit provinces of Gauteng, Western Cape and Kwa-Zulu Natal (Government of South Africa, 2020c). The South African economy like in many other countries with similar COVID-19 measures was negatively affected due to limitations in business and trade (Arndt et al., 2020). South Africa gradually eased restrictions to the National Lockdown Level Alerts 4,3,2 and 1 by permitting businesses to trade, easing curfews, gathering capacity and movement restrictions as shown by the difference in the Alert Level policy summaries provided in Table 4. On the 17^th^ of August 2020, the national lockdown alert level was adjusted to level 1 allowing for normal activities to resume with the strict condition of hygiene protocols being followed.

**Table 4:** South African National Lockdown Alert Level Policy Implemented in the period of the First National COVID-19 Epidemic Wave [24,56–60]

|  |
| --- |
| **No Lockdown Summary (2020/03/05-2020/03/25), (21 Days) [24]:** |
| Declaration of State of Emergency and establishment of National Coronavirus Command Council to oversee the COVID-19 outbreak in South Africa. |
| **Alert Level 5 Summary (2020/03/26-2020/04/30), (34 Days) [56]:** |
| South African border and air space were closed except for ports for the transportation of essential goods. Entry and exit screening at borders. Restriction on the movement of persons and goods. Persons confined to their residence with limitation to essential services. To enforce this South African Defence Force was deployed to oversee the compliance of this measure. Gatherings prohibited except for funerals. Movement between provinces, metropolitan and districts areas prohibited. Business Activities restricted to essential services and goods. Essential services must be provided with social distancing and hygienic measures. Public transport prohibited except bus (no more than 50 % capacity), taxi (no more than 70 % capacity) and e-hailing/private services (no more than 60 % capacity) for essential services. Identified Essential goods were the following: Food, Cleaning and Hygiene Products, Medical, Basic goods (ie electricity), Fuel, Hardware. Prohibition on evictions. Establishment of "COVID-19 Tracing Database" under the South African Department of Health. Establishment of Screening and Testing program under the South African Department of Health. Isolation, Quarantine of potentially infected persons and contact tracing protocols through testing programmes. Use of PPE for healthcare workers (high type variation including Isolation PPE), essential services (moderate type variation) and the general population (low type variation in general masks). |
| **Alert Level 4 Summary (2020/05/01-2020/05/31), (30 Days) [57]:** |
| Similar to Level 5 regarding Movements of persons except walk, run, cycle between 06h00 to 09h00 within 5 km radius of residence permitted, non-essential services in Table 1 of [57] permitted, a curfew was issued from 20:00 until 05:00 with most travel being restricted to the essential services. Movement between provinces, metropolitan areas and districts are prohibited with additional exception of learner's commute to higher education institutions permitted, return of dislocated persons to residences permitted, movement of children permitted. Attendance of funerals limited to 50 people. Sale, dispensing or transportation of liquor prohibited except for industries producing sanitisers and disinfectants. The sale of tobacco, tobacco products, e-cigarettes and related products prohibited. Table 1 services: 1) Agriculture, Hunting, Forestry and Fishing 2) Electricity, Gas and Water Supply 3) Manufacturing 4) Construction 5) Retail trade 6) ICTS 7) Media 8) Financial and Business 9) Accommodation and Food 10) Transport and Storage 11) Mining 12) Repair and Related Emergency 13) Supply Chain 14) Household Employment 15) Public Administration and Government Services 16) Health, Social and Personal Services 17) Education. Table 1 Services must have COVID-19 compliance officer, develop plan for phased return on employees and health protocols. |
| **Alert Level 3 Summary (2020/06/01-2020/08/17), (77 Days) [58]:** |
| Similar to Level 4 regarding Movements of persons except curfew was issued from 23:00 until 04:00. Movement between provinces, metropolitan areas and districts are prohibited with the additional exception of interprovincial travel. All business activities permitted with closing time for some services restricted to 22:00. Mandatory protocols when in a public place including wearing of face mask. Face mask defined as 'a cloth face mask or a homemade item that covers the nose and mouth, or another appropriate item to cover the nose and mouth.' Gatherings limited to 50 persons for indoor facilities and 100 persons for outdoor facilities. For public transport, bus and taxi services (no more than 70 % capacity if short distance travel and 100 % capacity permitted for long-distance travel) permitted. Long-distance travel defined as 200 km or more. Sale, dispensing or transportation of liquor permitted with sales from 10:00 to 18:00 from Mondays to Thursdays for off-site consumption and 10:00 to 22:00 for on-site consumption and transportation of liquor permitted. |
| **Alert Level 2 Summary (2020/08/18-2020/09/20), (32 Days) [59]** |
| Similar to Level 3 regarding Movements of person except curfew was issued from 22:00 until 04:00. Specific economic exclusions: 1) Night clubs. 2) International passenger air travel for leisure purposes. 3) Passenger ships for international leisure purposes 4) Attendance of any sporting event by spectators. 5) International sports events. |
| **Alert Level 1 Summary (2020/08/17-2020/09/20), (11 Days) [60]** |
| Most normal activity can resume, with precautions and health guidelines followed at all times. Curfew was issued from 00:00 until 04:00. Gatherings limited to 250 persons for indoor facilities and 500 persons for outdoor facilities. |

The following are NPIs that can be identified from South Africa’s COVID-19 policy response implemented through the National Lockdown Alert Levels:

- Entry and exit screening at borders.
- Limitations of movements in the form of national, provincial, district lockdowns and curfews.
- Ban/limitation of mass gatherings.
- Closure/Limitations of institution and business activities which included the closure of entertainment establishments, schools, higher tertiary institution, non-essential services.
- Ban/limiting of alcohol and tobacco industries, later banning/limiting liquor licence operating hours.
- Isolation, Quarantine of potentially infected persons and contact tracing protocols through testing programmes.
- Use of PPE for healthcare workers (high type variation including isolation PPE), essential services (moderate type variation) and the general population (low type variation in general masks).
- Hygienic protocols including social distancing, widespread use of sanitiser and frequent hand washing.

The COVID-19 NPIs implemented by the South African government were similar to those implemented globally in the Influenza (H1N1) Pandemic of 1918 (Billing, 2005). They show a focus on restricting contact between individuals within the population. The ban/limiting of the alcohol and tobacco industries were meant to reduce the pressure in trauma wards (by reducing the incident of accidents reporting to these wards) particularly car accident cases and also improving general social behaviour adherence to NPIs [61].

Figure 3 shows the South African Google Community Mobility Report in retail and recreation, grocery and pharmacy, parks, transit stations, workplaces and residences during the period of 2020/02/15 to 2020/10/01. The Google Community Mobility Reports are aimed at providing an understanding of the change in community movement in response to COVID-19 policies. The reports are generated using Google account information from people’s devices who have a location history turned on [62]. In 2017, South Africa had 61.8 % of their households with at least one member who had access to or used the internet [63]. As of 2020, there were 36.54 million internet users, 22 million social media users and 103.5 million mobile connections in South Africa [64]. The high penetration of the internet in South Africa shows the potential of using information gathered from the internet in understanding the South African community as done by the Google Community Mobility reports.

**Figure 3:**
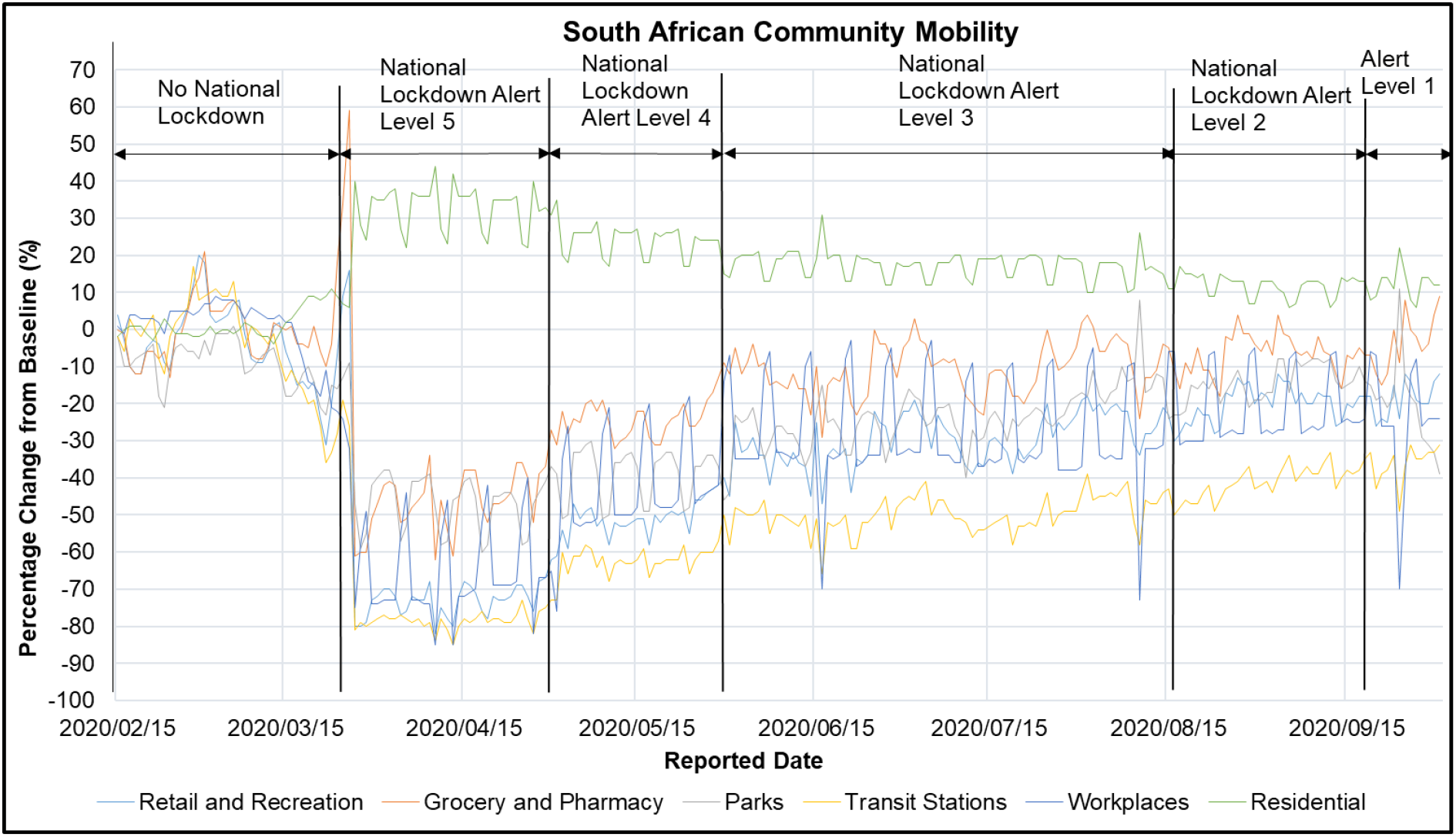
South African Community Google Mobility in Retail and Recreation, Grocery and Pharmacy, Parks, Transit Stations, Workplaces and Residences during the period of 2020/02/15 to 2020/10/01 [62]

Figure 3 shows that there was a spike in the South African grocery and pharmacy locations of 34 % and 53 % from baseline on the 25^th^ and 26^th^ of March 2020 respectively. These dates correspond to 1 day prior and a day into the implementation of the National Lockdown Alert Level 5. The spike was due to panic buying of groceries and medication by South Africans in anticipation of the COVID-19 pandemic and the lockdown implementation [65,66]. Figure 3 shows that implementation of the National Lockdown Alert Level 5 resulted in an increase in the South African residential location by 33±6 % from baseline while the retail and recreation, grocery and pharmacy, parks, transit stations, workplaces locations decreased by -73±4 %, -46±9 %,-47±7 %,-78±2 % and - 66±12 % respectively. These results suggest that the National Lockdown Alert Level 5 was effective in reducing the movement of communities in these locations in South Africa. Implementation of the National Lockdown Alert Level 4 resulted in a decrease in the South African residential location compared to the Alert Level 5 by 10 % (23±5 % from baseline). This is due to exceptions of walking, running, cycling and the introduction of a curfew system in the National Lockdown Alert Level policies. The National Lockdown Alert Level 4 resulted in a decrease in the retail and recreation, grocery and pharmacy, parks, transit stations, workplaces locations by -50±5 %, -23±5 %,-39±7 %,-62±4 % and -41±15 % from baseline respectively. These results reflect the impact on movement due to the relaxed policies in the National Lockdown Alert Level 4 from the Alert Level 5. Figure 3 shows that implementation of the National Lockdown Alert Level 3 resulted in a decrease in the South African residential location compared to the Alert Level 4 by 6 % (17±4 % from baseline). The National Lockdown Alert Level 3 resulted in a decrease in the retail and recreation, grocery and pharmacy, parks, transit stations, workplaces locations by -30±7 %, -11±7 %,-23±7 %,-50±5 % and -28±14 % from baseline respectively. In general, these results reflect the impact on movement due to the relaxed policies such as permitting all businesses to operate under strict hygienic protocols, allowance of interprovincial travel and a decreased curfew period in the National Lockdown Alert Level 3 from the Alert Level 4. The grocery and pharmacy and workplaces locations were the most impacted. The implementation of the National Lockdown Alert Level 2 resulted in a decrease in the South African residential location compared to the Alert Level 3 by 5 % (12±3 % from baseline). The National Lockdown Alert Level 2 resulted in a decrease in the retail and recreation, grocery and pharmacy, parks, transit stations, workplaces locations by -19±4 %, -6±5 %,-14±4 %,-40±4 % and -22±10 % from baseline respectively. The implementation of the National Lockdown Alert Level 1 resulted in a decrease in the retail and recreation, grocery and pharmacy, parks, transit stations, workplaces locations by -19±5 %, -3±8 %,-21±13 %,-37±5 % and -26±17 % from baseline respectively. The results in Figure 3 show that the National Lockdown Alert Level 1 and 2 had a similar impact on the movement in the South African communities. The results in Figure 3 also show that the impact of the National Lockdown Alert Level 3 on movement in South Africa was 32±9 % more than that in Alert Level 2 for all locations studied. Similar results were observed between the National Lockdowns Alert Level 5 and 4 and 4 with 3.

Table 5 shows the ARI COVID-19 SEIR Model effective daily contact number (β), observations (Obs), pooled variance, degree of freedom (df), t-statistical value (t Stat), P-value (P(T<=t) two-tail), t-Critical value, reduction in β for hypothesized mean difference of 0 between reported case data and ARI COVID-19 Model Data at a P-value of 0.05 for South Africa No Lockdown, National Lock Down Alert Level 5,4 and 3 Scenarios. The results in Table 5 were determined using non-linear regression analysis. The ARI COVID-19 SEIR Model Residual (Normalised Error) and Statistical Regression Plots between Model Data and Seeding Reported Case Data are shown in Figure A. 1 to Figure A. 2 in Section 8:Appendix. A non-linear regression analysis could not be conducted for the National Lockdown Alert Levels 2 and 1 due to these policies being implemented in the negative exponential phase of the first COVID-19 epidemic wave in South Africa. The limitation of non-linear regression analysis in the negative exponential phase of an epidemic is that the decrease in cases is not only due to reduction in contact but also due to the reduction in the susceptibles and increase in the recovered cases in the population (decrease in the mass incident term) at disease-free equilibrium. The t Stat, P(T<=t) two-tail and t Critical two-tail values in Table 5 show that Model and Reported Case Data used in the seeding period was significantly similar.

**Table 5:** ARI COVID-19 SEIR Model Effective Daily Contact Number (β), Observations (Obs), Pooled Variance, Degree of Freedom (df), t-statistical value (t Stat), P-Value (P(T<=t) two-tail), t-Critical value, Reduction in β for hypothesized mean difference of 0 between Reported Case Data and ARI COVID-19 Model Data at P-value=0.05 for South Africa No Lockdown, National Lock Down Alert Level 5,4 and 3 Scenarios.

| COVID-19 Policy Response | $\beta(\text{day}^{-1})$ | Obs | Pooled Variance | df | t Stat | $P(T \leq t)$ two-tail | t Critical two-tail | Reduction in $\beta$ (%) |
| --- | --- | --- | --- | --- | --- | --- | --- | --- |
| No Lockdown | 0.498 | 21 | 56895 | 40 | -0.112 | 0.911 | 2.02 |  |
| National Lock down Alert Level 5 | 0.144 | 35 | 1041 | 68 | -0.0986 | 0.922 | 2.00 | 71.1 |
| National Lock down Alert Level 4 | 0.196 | 21 | 53840 | 40 | 0.275 | 0.785 | 2.02 | 60.6 |
| National Lock down Alert Level 3 | 0.208 | 22 | 512005891 | 42 | -3.06 | 0.00382 | 2.02 | 58.1 |

Table 5 shows that the effective SARS-CoV-2 daily contact number (β) in South Africa was 0.498, 0144, 0.196, 0.208 day^-1^ for the no lockdown, National Lock down Alert Level 5, 4 and 3 model scenarios respectively. These results translate into a reduction of 71.1 %, 60.6 %, 58.1 % in the effective daily contact number from having no lockdown in South Africa to the implementation of the National Lockdown Alert Level 5, 4 and 3 respectively. Table 5 shows that the difference between the reduction in the effective daily contact number in the National Lock down Alert Level 4 and 3 was 4.13 % while that between Alert Level 5 and 4 was 14.6 %. These results suggest that the difference between movements in all locations studied and the National Lockdown Alert Levels implemented (which was approximately 31 % to 36 %) only had a 4.13 % to 14.6 % impact on the COVID-19 effective daily contact number. These results also suggest that the National Lockdown Alert Level 3 was as effective as the Alert Level 4 in reducing the COVID-19 effective daily contact number in South Africa.

Several factors contributed to the South African population’s preparedness to follow the COVID-19 policies implemented by the South African government. These factors include socio-economic status, age, education, and whether or not families care for vulnerable individuals like children or the elderly [67]. People who lived in informal dwelling settlements found it particularly difficult to isolate adequately during the National lockdown Alerts Level 5 and 4 when movement restrictions were strict. Due to South Africa’s history of racial segregation as well as apartheid, other race groups were more prepared to self-isolate compared to the Black population. Another challenge that was faced was a shortage of PPE for health workers resulting in workers either using torn PPE or working without them [68]. There was an increase in home deaths by those who are critically ill with Covid-19 or other diseases because they were afraid of going to public hospitals [44,69]. The South African public has a grown a large sense of trust in private hospitals which led to private hospital reaching their maximum capacity resulting in patients being transferred to public hospitals [69]. There was limited access to COVID-19 patients and other patients in hospitals during the early policy response. However, these policies were eased to allowing one visitor at a time for fifteen minutes whilst observing NPIs [70].

Covid-19 and the policies formulated to help with reducing the spread of the SARS-CoV-2 had adverse effects on the South African economy, more especially on Micro Small and Medium Enterprises (MSMEs) and informal workers and their households. The largest impact was the sudden loss of demand and revenue for Small and Medium Enterprises (SMEs) causing liquidity shortages [71,72]. Additionally, since SMEs are labour-intensive they were exposed to disruption during lockdowns where their workforces are required to quarantine [71]. In efforts to keep the economy from crumbling, the Government of South Africa presented the government’s Economic Reconstruction and Recovery Plan (ERRP) to help restore the economy [73]. This COVID-19 stimulus package, which was announced on April 21, 2020, amounted to 10 per cent of the country’s GDP ($26 Billion). The stimulus package would be directed to help the health sector municipalities that provide basic services, wage protection through the Unemployment Insurance Fund (UIF), financial supports for SMEs, and the credit guarantee scheme [73].

### 3.2 South Africa COVID-19 First Epidemic Wave Testing Data

COVID-19 testing was an important tool in the isolation, quarantine of potentially infected persons and contact tracing protocols in South Africa. COVID-19 tests can be classed into two categories either viral (Genome sequencing/reverse transcriptase PCR (rRT-PCR/ antigen) or serological (antibody) [74]. Viral tests can detect the genetic material of the virus and thus can determine if a person is currently infected with SARS-CoV-2. Samples or specimens for testing are usually taken through nasopharyngeal and oropharyngeal swab (upper respiratory specimens) and sputum, endotracheal aspirate, bronchoalveolar lavage (lower respiratory specimens). Specimens must be swiftly transferred to laboratories, stored and shipped between 2°C to 8°C or they may be frozen at -20°C with recommendations for freezing at -70°C. Viral test for SARS-Cov-2 in the specimen is then conducted in laboratories through the real-time reverse transcriptase-polymerase chain reactions (rRT-PCR). The COVID-19 rRT-PCR is a nucleic acid amplification test (NAAT) [75]. A positive COVID-19 rRT-PCR indicates that the specimen collected has SARS-CoV-2 thus the person from which the specimen is collected has a SARS-CoV-2 infection. It must be noted a negative rRT-PCR does not rule out the possibility of a SARS-CoV-2 infection. Negative tests can also be caused by the poor quality specimen, cross-contamination, specimen collected too early or late into the infection, specimen not handled appropriately and technical limitations such as viral mutation and PCR inhibition [75]. A positive COVID-19 rRT-PCR is considered accurate and usually not repeated. The time from sampling to result in a report for this test can take from less than 24 hours to up to a week depending on the laboratory testing demand and resources.

Serological tests detect antibodies in the blood generated by the immune response to an infection. They include a lateral-flow antibody, bead-based, enzyme-linked immunosorbent assays, and automated serology platforms. These essays assess the presence of Immunoglobulin M (IgM) and Immunoglobulin G (IgG). Antibodies can take up to 1 to 3 weeks to develop after infection and may stay in the blood for several days after recovery [9]. However, in acute infections, there is a potential waning of the antibodies post-infection [76]. IgM develops in the innate phase of the immune response and IgG in the later stages of the infection [9]. For COVID-19, the development of IgM has been observed to occur 5 to 7 days after the onset of symptoms and IgG, 10 to 14 days after symptom onset [76]. With the incubation period of COVID-19 being between 4 to 5 days [5], the use of serological testing is not recommended in this period [76]. Specimens for the COVID-19 serological test are obtained through a finger stick (using a bloodletting set) or blood draw. Results can be obtained from less than 24 hours to 1 to 3 days after the test. COVID-19 serological tests have a limitation in sensitivity and specificity with the sensitivity of the tests ranging from 33.3 % to 65.5 % [76]. A positive COVID-19 serological test usually requires repetition for confirmation. A negative COVID-19 serological test does not exclude past or current infection due to potential waning or low levels of antibodies [76]. The National Health Laboratory Service (NHLS) and National Institute for Communicable Diseases (NICD) were the national laboratories conducting the COVID-19 testing in South Africa and for other Southern African countries such as Lesotho, Namibia and Eswatini in their earlier COVID-19 epidemics. Major private laboratories involved in COVID-19 testing in South Africa included Abbott, Ampath, Pathcare and Lancet Laboratories [77].

Table 6 shows the South African average COVID-19 daily testing capacity, reported test positivity, cumulative tests, tests per million for the period reported of 2020/02/07 to 2020/10/01. Table 6 shows that the cumulative COVID-19 tests for the period reported in 2020/02/07 to 2020/10/01 was 5 383 078 COVID-19 tests. According to the NICD, in the period reported of 2020/03/01 to 2020/10/03, there were 3 705 951 laboratory tests for SARS-COV-2 conducted nationally [78]. Laboratory testing in South Africa was conducted for persons under investigation (PUI) which included community screening and testing programmes that were initiated in April 2020 and discontinued in the week of 17^th^ of May 2020. Testing was performed using rRT-PCR and laboratories used in-house and/or commercial PCR assays to conduct testing for the presence of SARS-CoV2 RNA [78]. The difference in the cumulative COVID-19 tests in Table 6 and those reported by the NICD, 2020d can be attributed to: (i) difference in sources reporting, (ii) cumulative COVID-19 tests in Table 6 including serological tests. According to the NICD, South African public and private sector laboratories conducted 45.9 % and 54.1 % of the cumulative COVID-19 tests respectively in the period reported in 2020/03/01 to 2020/10/03. Table 6 shows that the average COVID-19 testing capacity per day in South Africa for the first COVID-19 epidemic wave was 18 069±13760 COVID-19 tests. Table 6 also shows that the average reported COVID-19 positive cases to testing in South Africa for the first COVID-19 epidemic wave was 14.7 %. It must be noted that the COVID-19 Positive cases to Testing ratio particularly if the majority of the testing is through rRT-PCR are not an indicator of the seroprevalence in the population. According to an NICD report, the seroprevalence in the Cape Town metropolitan sub-districts (Western Cape Province) after the peak of the first epidemic infections was 39 % [79].

**Table 6:**
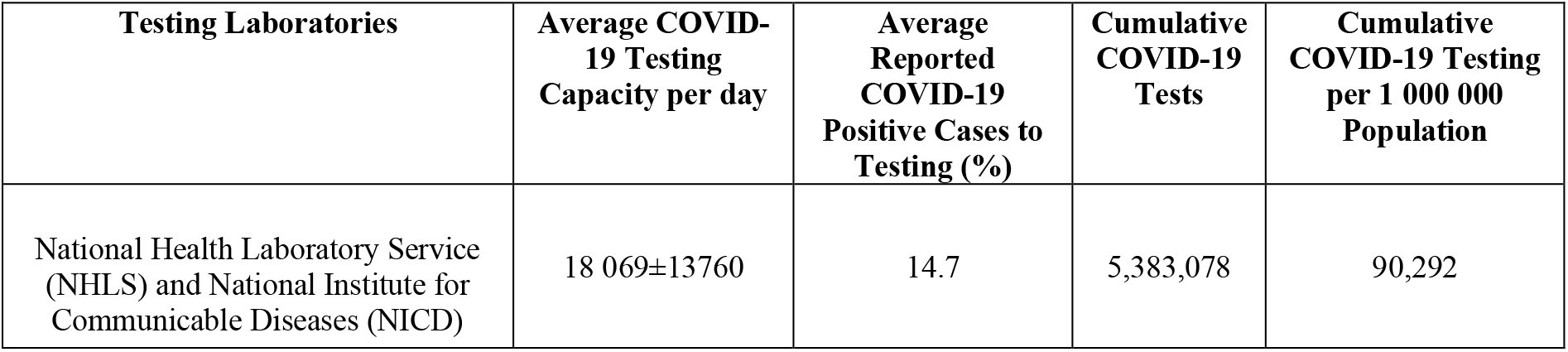
South African Average COVID-19 Daily Testing Capacity, Reported Test Positivity, Cumulative Tests, Test per Million for the period 2020/02/07 to 2020/10/01 [80]

| **Testing Laboratories** | **Average COVID-19 Testing Capacity per day** | **Average Reported COVID-19 Positive Cases to Testing (%)** | **Cumulative COVID-19 Tests** | **Cumulative COVID-19 Testing per 1 000 000 Population** |
| --- | --- | --- | --- | --- |
| National Health Laboratory Service (NHLS) and National Institute for Communicable Diseases (NICD) | 18 069±13760 | 14.7 | 5,383,078 | 90,292 |

Figure 4 shows the South African COVID-19 daily testing and cases for the period reported in 2020/02/07 to 2020/10/01. Figure 4 shows that the COVID-19 testing in South Africa fluctuated daily during the first COVID-19 epidemic wave. The general trend shows a positive correlation between COVID-19 testing and cases in the first COVID-19 epidemic wave in South Africa. COVID-19 testing was limited and challenging in South Africa in the initial stages of the COVID-19 pandemic. This was due to the limited supply and global competition for the resources to perform COVID-19 testing such as reagents, equipment and assays [81]. COVID-19 testing increased up to 57 000 tests per day in South Africa in the period of 2020/03/13 to 2020/07/17. The correlation of COVID-19 testing and cases shows that testing has an impact on how the epidemic is observed/reported. Increasing testing increases the accuracy of case reporting however this is limited by the testing approach. Random testing can aid in increasing the accuracy of the viro-prevalence and seroprevalence of COVID-19. However, most COVID-19 testing in South Africa has been targeted (non-random) which includes contact tracing efforts [78]. Another limitation in reported COVID-19 cases was that asymptomatic COVID-19 cases were difficult to identify in the population due to a lack of symptoms. Studies have shown a high proportion of asymptomatic COVID-19 cases in COVID-19 reported cases [47–49]. The result is that there is a probability that some COVID-19 cases in South Africa were not reported.

**Figure 4:**
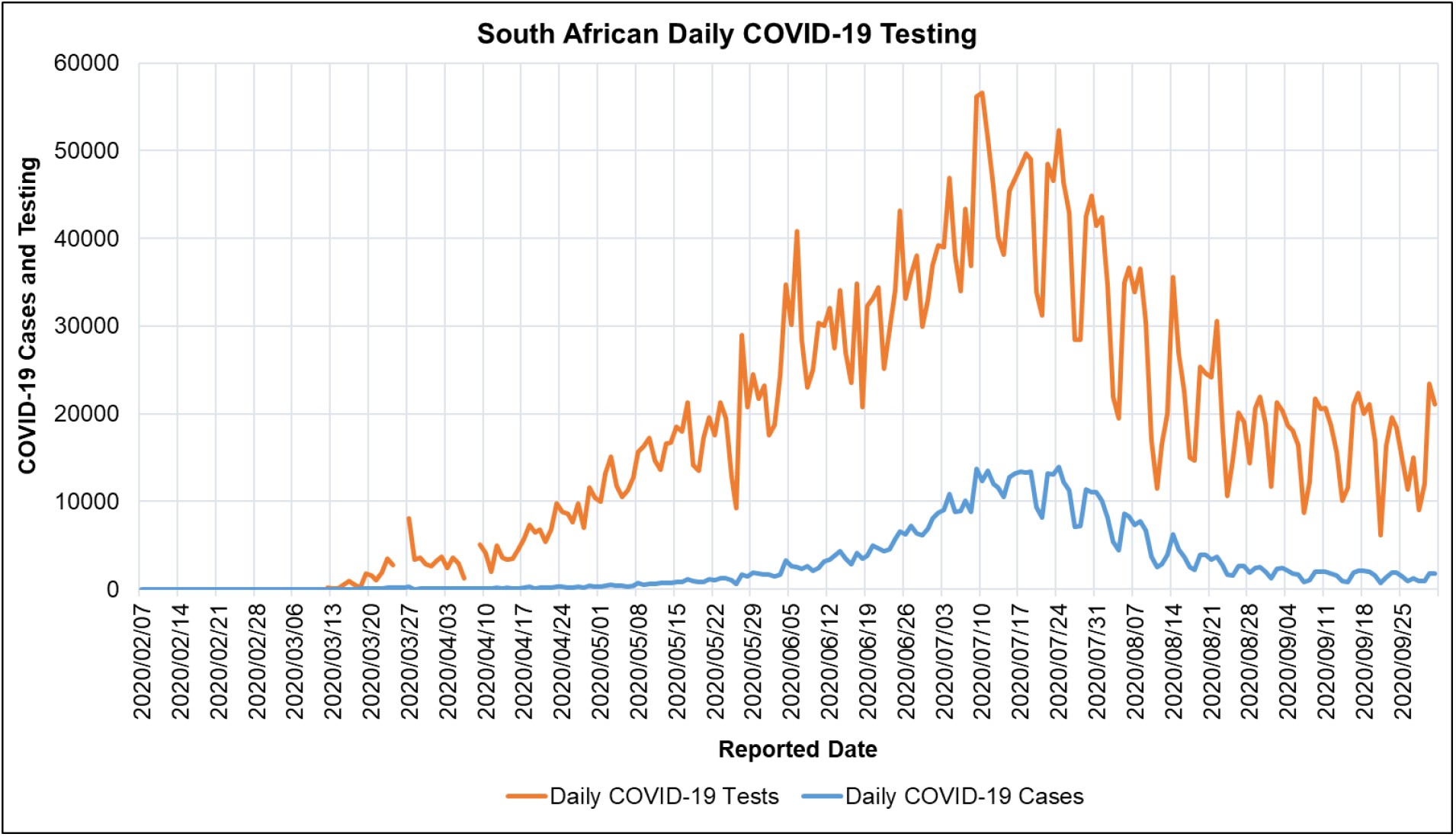
South African COVID-19 Daily Testing and Cases for the period 2020/02/07 to 2020/10/01 [80]

### 3.3 South Africa COVID-19 First Epidemic Wave Reported Case Data

Figure 5 shows the cumulative, recovered and active South African COVID-19 cases and deaths for the period reported in 2020/01/22 to 2020/10/01. Figure 5 shows that the cumulative, recovered COVID-19 cases and deaths in South Africa in the respective reported period were 676 084, 609 584 and 16 866 respectively. 90.2 % of the COVID-19 cases reported in the respective reported period recovered. Figure 5 shows that the National Lockdowns Alert Level 5 and 4 were implemented at the start of the epidemic however the COVID-19 policy in South Africa was then eased to Lockdown Alert Level 3 (2020/06/01-2020/08/17) where the majority of the positive exponential phase of the first epidemic wave was observed. The COVID-19 policy was further eased to the National Lock down Alert Level 2 and 1 in the negative exponential phase of the first epidemic wave. The first COVID-19 epidemic wave in South Africa lasted for 205 days from the first reported case. According to the Network for Genomics Surveillance in South Africa (NGS-SA), at least 101 introductions of SARS-CoV-2 were estimated in South Africa, with the bulk of the important introductions occurring before lockdown from Europe. South African genomes in the period of 2019/12/24-2020/08/26 were assigned to 42 different lineages with 16 South African specific lineages. The largest monophyletic linear clusters that spread in South Africa during the lockdown and then grew into large transmission cluster during the peak of infections during the first COVID-19 epidemic wave were the C.1, B.1.1.54, B.1.1.56 lineage clusters. These main lineages accounted for 42 % of all sampled South African sequences (1365 South African genomes). Genomes belonging to these lineages were sampled in five provinces in South Africa (North-West, Limpopo, KwaZulu-Natal, Gauteng and Free State) [82]. Spatiotemporal phylogeographic analysis suggests that the variant of concern, SARS-CoV-2 501 Y.V2 lineage (B.1.351) emerged in early August after the peak and in the period of the negative exponential phase of the first COVID-19 epidemic wave in South Africa [83]. These results suggest that the C.1, B.1.1.54, B.1.1.56 lineage clusters were major drives of the first COVID-19 epidemic wave in South Africa.

**Figure 5:**
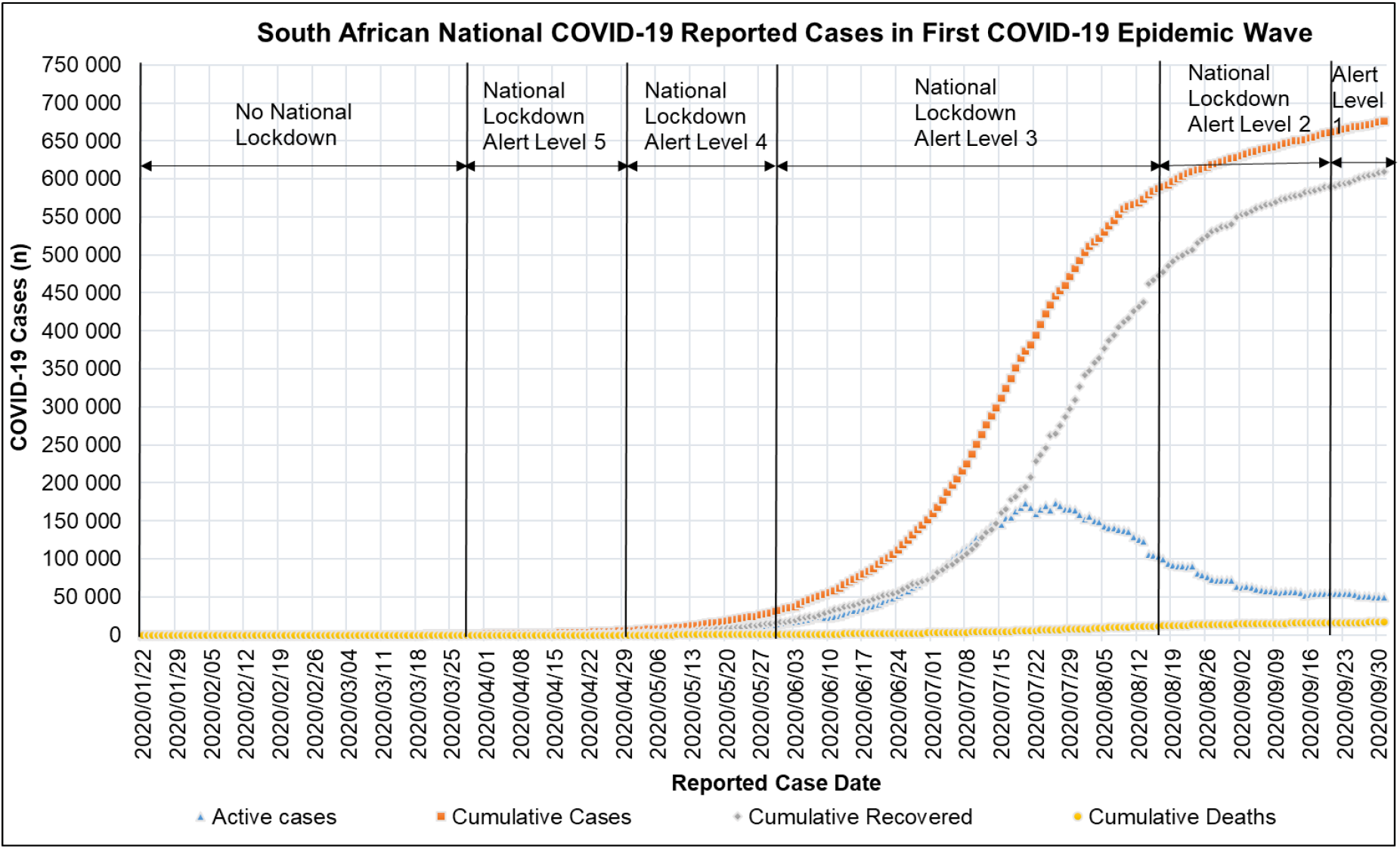
National Cumulative, Recovered and Active COVID-19 cases and deaths in South Africa for the period 2020/01/22 to 2020/10/01 [54]

Figure 6 shows the active provincial COVID-19 cases in South Africa for the period reported in 2020/01/22 to 2020/10/01. Figure 6 shows that the first COVID-19 epidemic wave in South African provinces had different amplitudes and periods. Epidemiologically this result can be explained by the district and provincial confinement of the South African population due to the National Alert Level Lockdowns, the difference in testing capacity, population, population distribution, residential settings and business activities in the provinces. Table 7 shows the provincial South African population, COVID-19 date of peak and peak active cases for the period reported in 2020/01/22 to 2020/10/01. Table 7 shows that the total peak COVID-19 active cases in South Africa’s first COVID-19 epidemic wave was 173 587 COVID-19 cases. Table 7 also shows that Gauteng, KwaZulu Natal, Eastern Cape, Western Cape provinces had 26.0 %, 19.3 %, 11.3 %, 11.8 % of the total first peak COVID-19 active cases respectively. This represents 68.4 % of the total peak COVID-19 active cases observed in South Africa’s first COVID-19 epidemic wave. The Western Cape province was the first province to observe a peak active cases on the 10^th^ of July 2020. While the Northern Cape province was the last province to observe a peak in Active Cases on the 5^th^ of September 2020. The South African national average date of peak active cases in the first COVID-19 epidemic wave was on the 26^th^ of July 2020.

**Table 7:** Provincial South African Population, Peak COVID-19 Date of Peak and Active Cases for the period 2020/01/22 to 2020/10/01 [84]

| Province | Observed Date of Peak Active COVID-19 Cases | Peak Active COVID-19 Cases (n) | Peak Active COVID-19 Cases (%) | Population (n) | Population (%) |
| --- | --- | --- | --- | --- | --- |
| Northern Cape | 2020/09/05 | 4000 | 2.30 | 1292786 | 2.2 |
| Limpopo | 2020/07/29 | 4136 | 2.38 | 5852553 | 9.8 |
| Mpumalanga | 2020/07/31 | 7169 | 4.13 | 4679786 | 7.8 |
| North West | 2020/07/23 | 11834 | 6.82 | 4108816 | 6.9 |
| Free State | 2020/07/30 | 18066 | 10.41 | 2928903 | 4.9 |
| Western Cape | 2020/07/10 | 18230 | 10.50 | 7005741 | 11.8 |
| Eastern Cape | 2020/07/20 | 19638 | 11.31 | 6734001 | 11.3 |
| KwaZulu Natal | 2020/08/02 | 44298 | 25.52 | 11531628 | 19.3 |
| Gauteng | 2020/07/20 | 77368 | 44.57 | 15488137 | 26.0 |
| **South Africa** | **2020/07/26** | **173587** | **100** | **59622350** | **100** |

**Figure 6:**
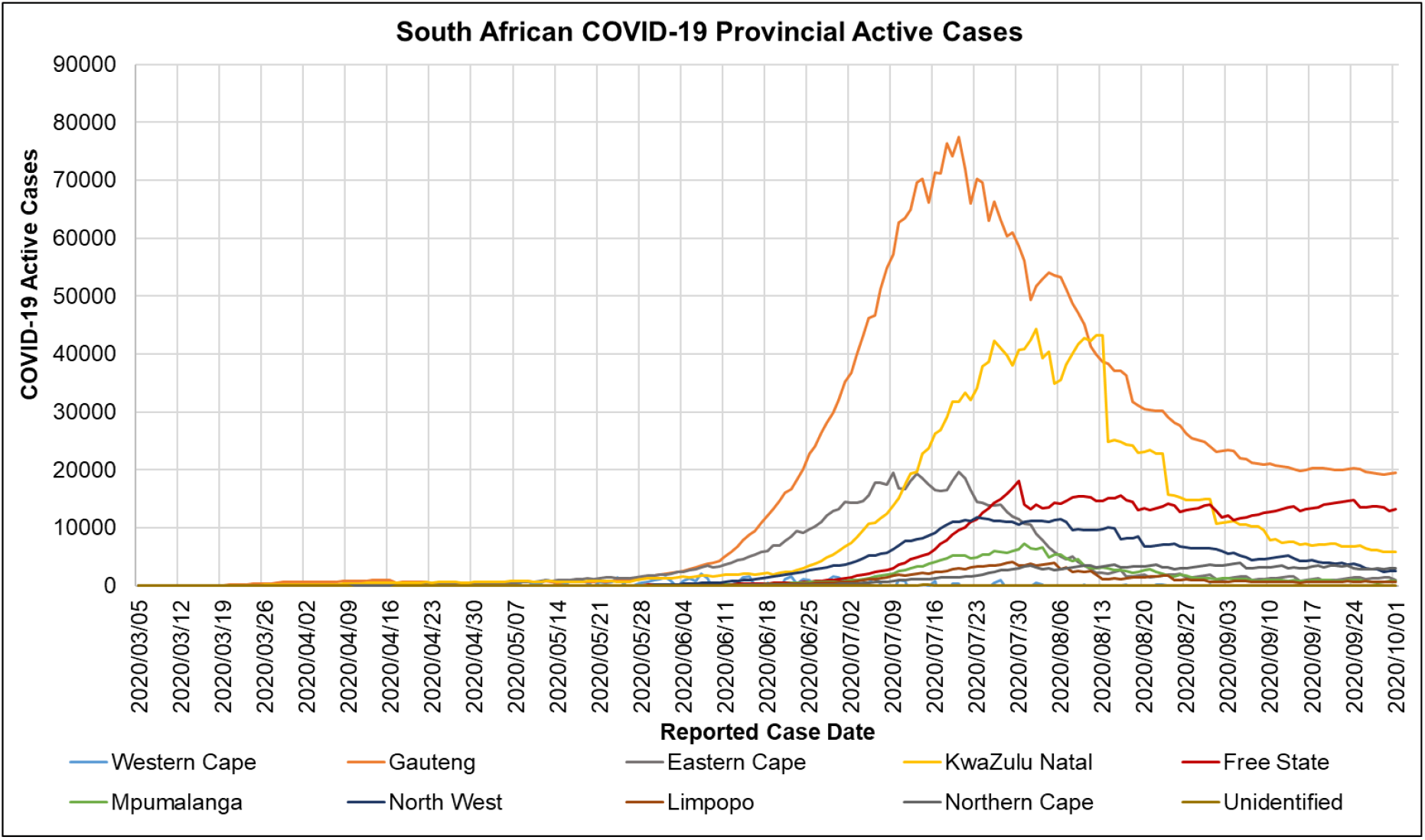
Active Provincial COVID-19 cases in South Africa for the period 2020/01/22 to 2020/10/01 [84]

### 3.4 South Africa COVID-19 First Epidemic Wave Hospitalised Cases

Figure 7 shows the admission status of COVID-19 patients in South African hospitals reported in the NICD DATCOV surveillance system during the period of 2020/05/24 to 2020/10/01. Figure 7 shows that the number of admitted patients increased from the period of 2020/05/24 to 2020/08/01 reaching a peak and then decreased thereafter. The peak of admitted COVID-19 patients in South African hospitals corresponds with the peak of active COVID-19 cases observed in South African provinces shown in Table 7. The general trend in Figure 7 shows that most COVID-19 patients were admitted to the general ward. At the peak of admitted COVID-19 patients, there was a total of 8319 patients with 5745, 1520, 989, 799, 763, 442 patients being admitted in the general ward, intensive care unit, on oxygen, on ventilators, in high care and the isolation ward respectively. Although the severity of COVID-19 in South Africa cannot be conclusively drawn from hospital admissions, the result of the trend observed in Figure 7 corresponds with the severity of COVID-19 described in the [12,85].

**Figure 7:**
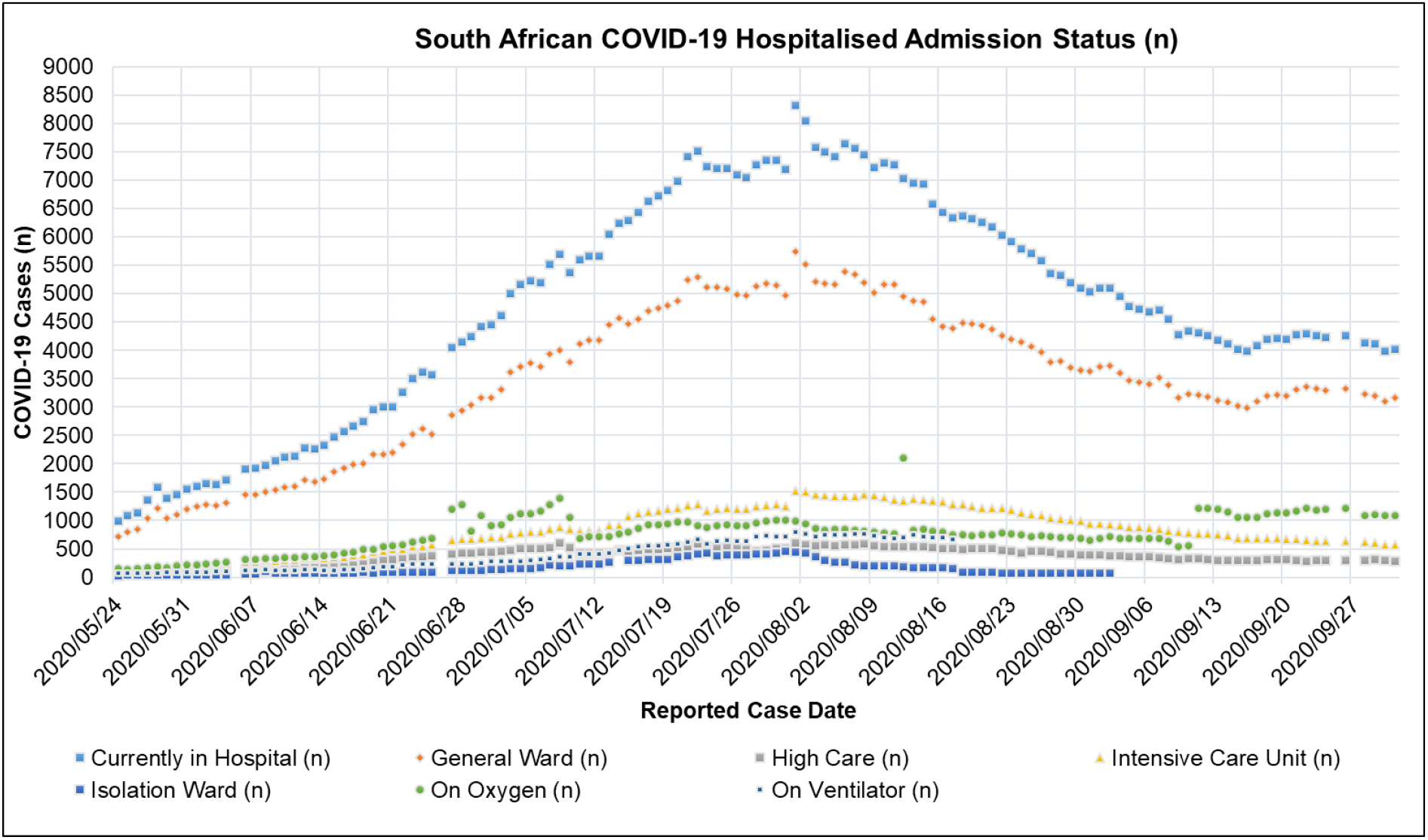
South African COVID-19 Hospitalised Admission Status: Currently in Hospital, General Ward, High Care, Intensive Care Unit, Isolation Ward, On Oxygen, On Ventilator for the period of 2020/05/24 to 2020/10/01 [41]

Table 8 shows the mean, standard deviation (STDev), standard error of mean (SE), lower and upper confidence interval at P-value of 0.05 for the South African COVID-19 discharge rate, hospitalised and the un-hospitalised fatality rate for the period of 2020/05/24 to 2020/10/01. Table 8 shows that the mean of the South African COVID-19 patient discharge rate was 11.9 days per patient. The mean of the South African COVID-19 patient case fatality rate (CFR) in hospital and outside the hospital was 2.06 %, 95% CI [1.86,2.25] (deaths per admitted patients) and 2.30 %, 95% CI [1.12,3.83](deaths per severe and critical cases) respectively. The COVID-19 CFR outside the hospital was observed to be higher than in the hospital.

**Table 8:** Mean, Standard Deviation (STDev), Standard Error of Mean (SE), Lower and Upper Confidence Interval at P-Value=0.05 for South African COVID-19 Discharge (T_disch)_, Hospitalised (μ_1_) and un-hospitalised Fatality Rate (μ_o)_ for the period of 2020/05/24 to 2020/10/01

| Parameter | Mean | STDev | SE | CI-Lower | CI-Upper |
| --- | --- | --- | --- | --- | --- |
| $\mu_1$ (CFR) | 0.0206 | 0.0110 | 0.0010 | 0.0186 | 0.0225 |
| $T_{disch}$ (Day) | 11.9 | 2.31E-04 | 1.05E-05 | 11.90 | 11.9 |
| $\mu_o$ (CFR) | 0.0230 | 0.0123 | 0.0011 | 0.0112 | 0.0383 |

Figure 8 shows a linear regression analysis done on the daily cumulative COVID-19 deaths and discharged patients in South African hospitals in the NICD DATCOV surveillance system for the period of 2020/05/24 to 2020/10/01. Figure 8 shows that cumulative COVID-19 deaths and discharged patients had a positive linear correlation with the reported case date. The correlation coefficient (R^2^) of the COVID-19 deaths and discharged patients with the reported case date was 0.9708 and 09675 respectively. These results show a constant positive gradient in the cumulative COVID-19 deaths and discharged patients per reported case date indicating a constant CFR and discharge rate in the respective period. A plot of the daily CFR and discharge rate in South African hospitals in the period of 2020/05/24 to 2020/10/01 is shown in Figure A. 4. The constant daily hospital CFR and discharge rate in South African hospitals indicate good clinical management in the face of adversity where the increasing number of cases towards the peak of the first epidemic wave did not influence the hospital COVID-19 discharge and death rate.

**Figure 8:**
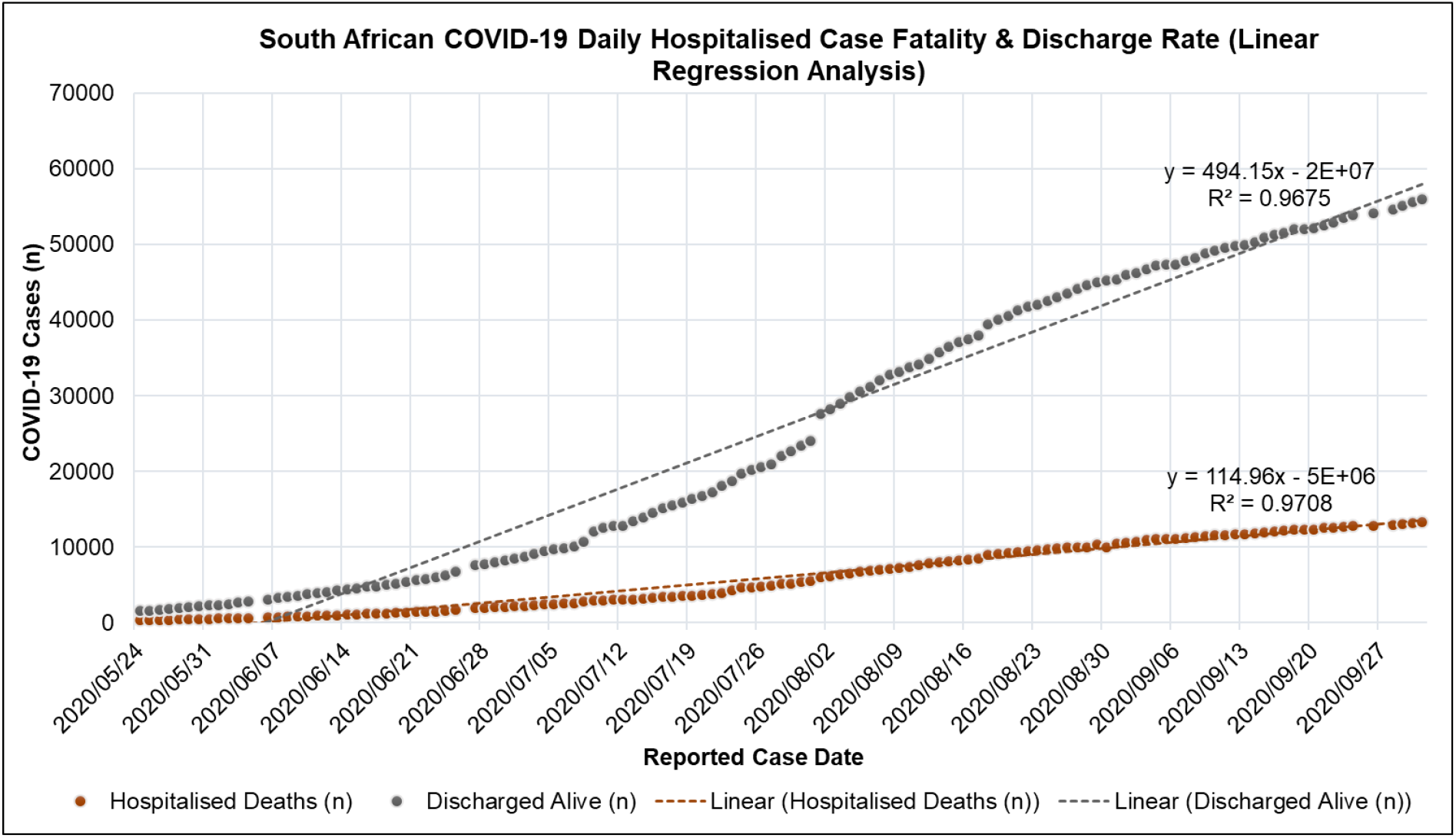
Linear regression of South African COVID-19 Hospitalised Case Fatality and Discharge Rate for the period of 2020/05/24 to 2020/10/01 [41]

Table 9 shows the mean, standard deviation (STDev), standard error of mean (SE), lower and upper confidence interval at P-value=0.05 for the South African proportion of COVID-19 admission status for the period of 2020/05/24 to 2020/10/01. Table 9 shows that the mean for the COVID-19 general ward, intensive care unit, on oxygen, high care, on ventilator, in isolation ward admission status in South African hospital was 58.5 %, 95% CI [58.1,59.0], 13.4 %, 95% CI [13.1,13.7], 13.3 %, 95% CI [12.6,14.0], 6.37 %, 95% CI [6.23,6.51], 6.29 %, 95% CI [6.02,6.55], 2.13 %, 95% CI [1.87,2.43] respectively. These results suggest that most COVID-19 patients reporting to South African hospitals were admitted into the general wards. The proportion reporting to intensive care units and on oxygen were similar regarding confidence intervals. A relatively low proportion of patients were admitted to the isolation ward. A plot of the daily proportion of the COVID-19 admission status in South African hospitals in the period of 2020/05/24 to 2020/10/01 is shown in Figure A. 3.

**Table 9:** Mean, Standard Deviation (STDev), Standard Error of Mean (SE), Lower and Upper Confidence Interval at P-Value=0.05 for the South African COVID-19 Hospital Admission Status for the period of 2020/05/24 to 2020/10/01

| Parameter | Mean | STDev | SE | CI-Lower | CI-Upper |
| --- | --- | --- | --- | --- | --- |
| General Ward (%) | 58.5 | 2.68 | 0.239 | 58.1 | 59.0 |
| High Care (%) | 6.37 | 0.81 | 0.073 | 6.23 | 6.51 |
| Intensive Care Unit (%) | 13.4 | 1.83 | 0.163 | 13.1 | 13.7 |
| Isolation Ward (%) | 2.13 | 1.47 | 0.131 | 1.87 | 2.43 |
| On Oxygen (%) | 13.3 | 3.91 | 0.348 | 12.6 | 14.0 |
| On Ventilator (%) | 6.29 | 1.50 | 0.134 | 6.02 | 6.55 |

Table 10 shows the mean, standard deviation (STDev), standard error of mean (SE), lower and upper confidence Interval at P-value=0.05 for the South African COVID-19 hospitalised case age profile for the period of 2020/05/24 to 2020/10/01. Table 10 shows that children in the age groups of 0 to 9 years and 10 to 19 years made up 2.32 %, 95% CI [2.21,2.4] and 1.75 %, 95% CI [1.71,2.4] of the COVID-19 hospitalised cases in South African hospitals respectively. This was relatively lower than other age groups reporting in South African hospitals indicating low case incident in the severe and critical COVID-19 disease in children. According to WHO, the COVID-19 disease in children is relatively rare with a small proportion of individual under 19 developing severe or critical symptoms [5]. In the case of South Africa, this is indeed the case, this phenomenon also being noted by the NICD [86]. People in the age groups of 40 to 49 years and 50 to 59 years made up 20.4 %, 95% CI [19.9,20.9] and 25.3 %, 95% CI [24.5,26.0] of the COVID-19 hospitalised cases in South African hospitals respectively. People in the age groups over 40 years accounted for 78.9 % of the COVID-19 hospitalised cases in South African hospitals. People in the age groups of 20 to 29 years and 30 to 39 years made up 8.04 %, 95% CI [7.65,8.44] and 18.1 %, 95% CI [17.5,18.7] of the COVID-19 hospitalised cases in South African hospitals respectively. A plot of the daily proportion of COVID-19 case age group profile in South African hospitals in the period of 2020/05/24 to 2020/10/01 is shown in Figure A. 5.

**Table 10:** Mean, Standard Deviation (STDev), Standard Error of Mean (SE), Lower and Upper Confidence Interval at P-Value=0.05 for the South African COVID-19 Hospitalised Case age profile for the period of 2020/05/24 to 2020/10/01

| Parameter | Mean | STDev | SE | CI-Lower | CI-Upper |
| --- | --- | --- | --- | --- | --- |
| Hospitalised Cases Age Group 0-9 (%) | 2.32 | 0.5785 | 0.0552 | 2.21 | 2.43 |
| Hospitalised Cases Age Group 10-19 (%) | 1.75 | 0.2190 | 0.0209 | 1.71 | 1.79 |
| Hospitalised Cases Age Group 20-29 (%) | 8.04 | 2.1040 | 0.2006 | 7.65 | 8.44 |
| Hospitalised Cases Age Group 30-39 (%) | 18.1 | 3.2549 | 0.3103 | 17.5 | 18.7 |
| Hospitalised Cases Age Group 40-49 (%) | 20.4 | 2.6277 | 0.2505 | 19.9 | 20.9 |
| Hospitalised Cases Age Group 50-59 (%) | 25.3 | 4.0561 | 0.3867 | 24.5 | 26.0 |
| Hospitalised Cases Age Group 60-69 (%) | 17.3 | 3.1225 | 0.2977 | 16.8 | 17.9 |
| Hospitalised Cases Age Group 70-79 (%) | 10.2 | 1.9700 | 0.1878 | 9.79 | 10.5 |
| Hospitalised Cases Age Group 80+ (%) | 5.66 | 1.1664 | 0.1112 | 5.44 | 5.88 |

Table 11 shows mean, standard deviation (STDev), standard error of mean (SE), lower and upper confidence interval at P-Value=0.05 for the South African hospitalised COVID-19 death age profile for the period of 2020/05/24 to 2020/10/01. Table 11 shows that the proportion of COVID-19 deaths in South African hospitals increased with an increase in age groups up to the age group 60 to 69 years. COVID-19 deaths of children in South African hospitals was relatively low with age groups 0 to 9 years and 10 to 19 years making up 0.21 %, 95% CI [0.19,0.2] and 0.28 %, 95% CI [0.27,0.4] of the COVID-19 deaths in South African hospitals respectively. People in the age groups of 50 to 59 years and 60 to 69 years had the highest proportion of COVID-19 deaths in South African hospitals. Both respective age groups made up 24.3 %, 95% CI [23.9,24.6] and 26.0 %, 95% CI [25.8,26.3] of the COVID-19 deaths in South African hospitals. A plot of the daily proportion of the COVID-19 death age group profile in South African hospitals in the period of 2020/05/24 to 2020/10/01 is shown in Figure A. 6.

**Table 11:**
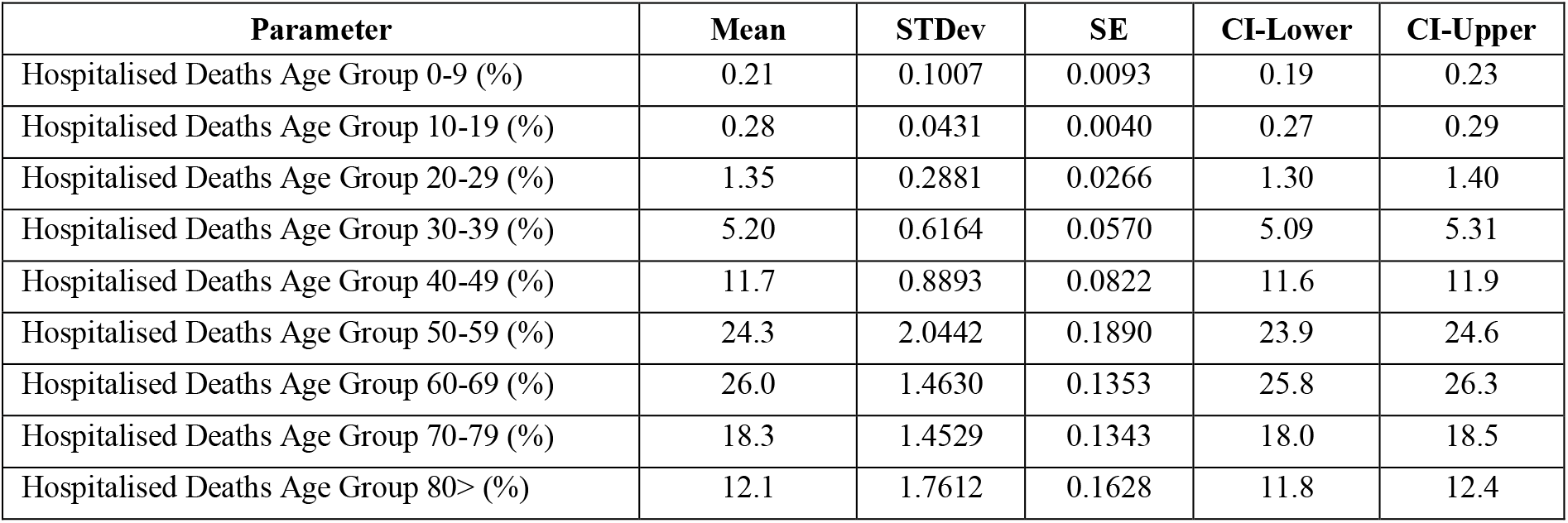
Mean, Standard Deviation (STDev), Standard Error of Mean (SE), Lower and Upper Confidence Interval at P-Value=0.05 for the South African hospitalised COVID-19 Death Age profile for the period of 2020/05/24 to 2020/10/01

Table 12 shows the cumulative COVID-19 death risk ratio for South Africa hospitalised age groups (Age Group 0-9 as reference (Ref)) at P-value of 0.05 for the period of 2020/05/24 to 2020/10/01. Table 12 shows that the risk of COVID-19 deaths in South African hospitals increased with increasing age groups. People in the age groups over 80 years had the highest risk of dying from COVID-19 in South African hospitals with a cumulative COVID-19 death risk ratio of 23.7, 95% CI [22.6,25.1] times more than the age group 0 to 9 years. Table 12 shows the risk of dying from COVID-19 in South African hospitals for age groups over 20 years approximately doubled with an increase in age of 10 years.

**Table 12:** Cumulative COVID-19 Death Risk Ratio for South Africa hospitalised age groups (Age Group 0-9 as reference (Ref)) at P-Value=0.05 for the period of 2020/05/24 to 2020/10/01

| Parameter | Cumulative Death Risk Ratio (Ref) | Cumulative Death Risk Ratio CI-Lower (Ref) | Cumulative Death Risk Ratio CI-Upper (Ref) |
| --- | --- | --- | --- |
| Hospitalised Cases Age Group 0-9 (%) | 1.00 | 1.00 | 1.00 |
| Hospitalised Cases Age Group 10-19 (%) | 1.77 | 1.71 | 1.84 |
| Hospitalised Cases Age Group 20-29 (%) | 1.87 | 1.78 | 1.97 |
| Hospitalised Cases Age Group 30-39 (%) | 3.19 | 3.03 | 3.37 |
| Hospitalised Cases Age Group 40-49 (%) | 6.39 | 6.09 | 6.75 |
| Hospitalised Cases Age Group 50-59 (%) | 10.7 | 10.1 | 11.3 |
| Hospitalised Cases Age Group 60-69 (%) | 16.7 | 15.7 | 17.8 |
| Hospitalised Cases Age Group 70-79 (%) | 20.0 | 18.8 | 21.3 |
| Hospitalised Cases Age Group 80> (%) | 23.7 | 22.6 | 25.1 |

Another cofounding factor in COVID-19 deaths in South African hospitals were disease comorbidities. In the period of 5 March to18 July 2020, in Western Cape, South Africa COVID-19 comorbidity with Diabetes was accounted for in most hospitalized cases followed by HIV then Hypertension at 38.5 %, 37.4 %, and 36.4 % respectively. While, chronic kidney, pulmonary, and tuberculosis (TB) accounted for 6.8 %, 12.3 %, and 11.8 % respectively. COVID-19 comorbidity with diabetes accounted for most reported deaths in Western Cape, South Africa followed by Hypertension at 55.1 % and 47.2 % respectively. While, HIV, Chronic Kidney disease, Asthma, and TB accounted for 16.2 %, 14.8 %, 11.5 %, and 3.2 % respectively [87].

### 3.5 Excess (Natural) Deaths in South Africa COVID-19 First Epidemic Wave

Figure 9 shows the South African weekly excess (natural) and COVID-19 reported deaths for the period of 2019/12/29 to 2020/10/01. Figure 9 shows that the first reported COVID-19 death in South Africa was on the 15^th^ of March 2020. Excess (Natural) deaths in South Africa started to exceed reported COVID-19 deaths in the weekly report of the 14^th^ June 2020. Figure 9 shows that weekly excess natural and COVID-19 reported deaths were characterised by positive exponential growth in the period of 2020/03/15 to 2020/07/26 and negative exponential decline thereafter. The peak of weekly excess natural and COVID-19 reported deaths observed on the 26^th^ of July 2020 in Figure 9 coincides with the peak active COVID-19 cases in South Africa observed in Section 3.3. The peak of weekly excess natural and COVID-19 reported deaths in South Africa’s first COVID-19 epidemic wave was 6676 and 2057 deaths respectively. A plot of the South African Cumulative Excess (Natural) Deaths and COVID-19 Reported Deaths for the period of 2019/12/29-2020/10/01 is shown in Figure A. 7.

**Figure 9:**
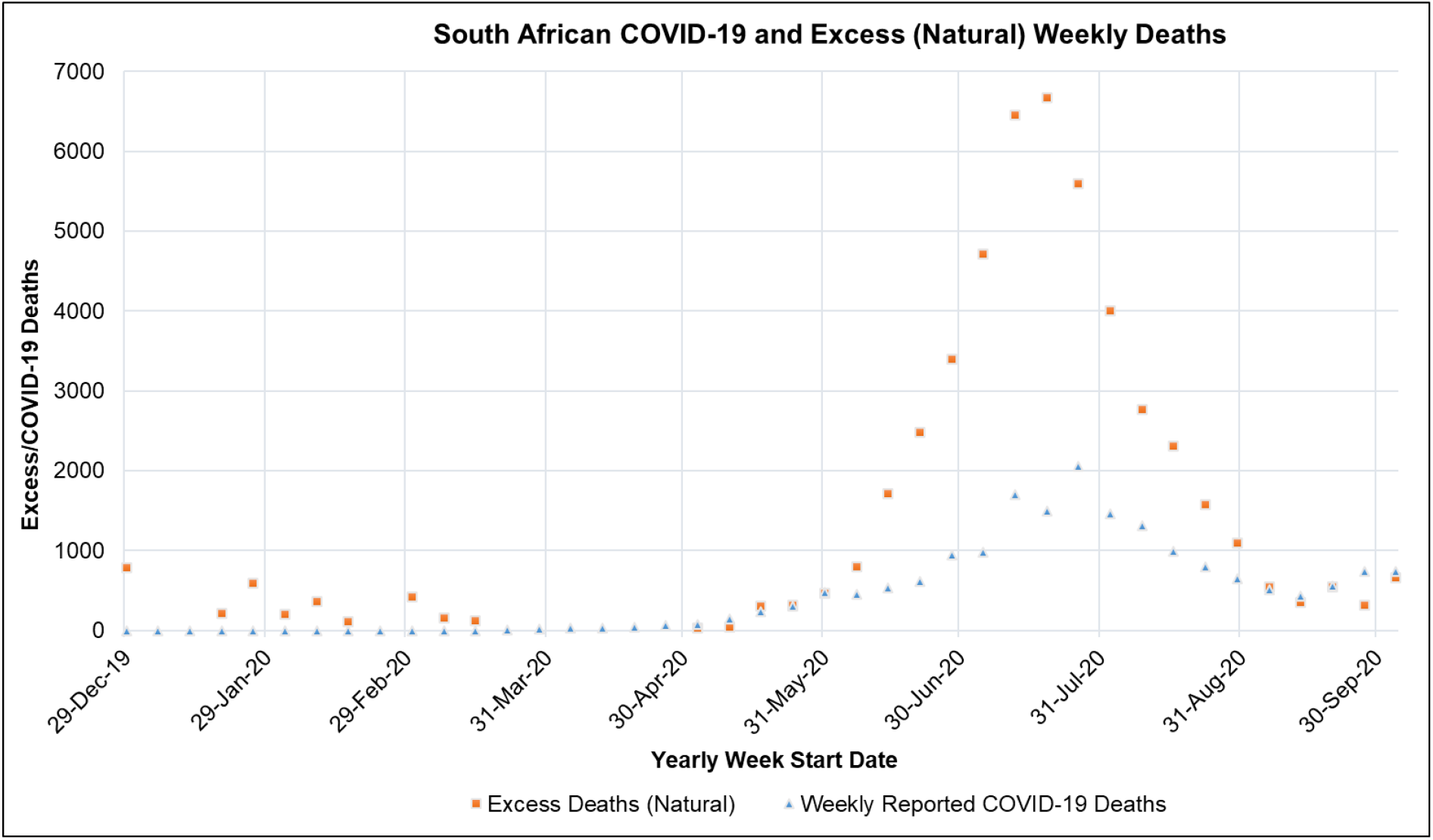
South African Weekly Excess (Natural) and COVID-19 Reported Deaths for the period of 2019/12/29-2020/10/01 [44,45]

Table 13 shows the mean, standard deviation (STDev), standard error of the mean (SE), lower and upper confidence interval at P-Value of 0.05 for South African weekly, excess deaths, excess (natural) to natural deaths and excess deaths (natural) to COVID-19 death ratio for the period from 2019/12/29 to 2020/10/01. Table 13 shows that the weekly excess (natural) deaths, excess (natural) to natural deaths (%), excess deaths (natural) to COVID-19 death ratio was 2114, 95% CI [1239,2990], 16.7 %, 95% CI [10.9,22.5] and 1.12, 95% CI [0.55,1.68] respectively.

**Table 13:** Mean, Standard Deviation (STDev), Standard Error of Mean (SE), Lower and Upper Confidence Interval at P-Value=0.05 for South African Weekly, Excess Deaths, Excess (Natural) to Natural Deaths and Excess Deaths (Natural) to COVID-19 Death Ratio for the period from 2019/12/29 to 2020/10/01

| Parameter | Mean | STDev | SE | CI-Lower | CI-Upper |
| --- | --- | --- | --- | --- | --- |
| Weekly Excess (Natural) Deaths | 2114 | 2095.9 | 446.842 | 1239 | 2990 |
| Excess (Natural) to Natural Deaths (%) | 16.7 | 13.8 | 2.940 | 10.9 | 22.5 |
| Excess Deaths (Natural) to COVID-19 Death Ratio | 1.12 | 1.346 | 0.287 | 0.55 | 1.68 |

The relatively low value of the excess (natural) to natural deaths in South Africa shows that the COVID-19 disease or epidemic did not account for the majority of the natural deaths occurring in South Africa in the respective period reported. This is reflective of the high disease burden in South Africa. The estimated crude death rate in South Africa in 2020 (*calculated in 2019*) was 9.5 per 1000 of the population [88]. In 2016, non-communicable diseases (NCD) in South Africa accounted for 51 % of the total deaths in South Africa [89]. Figure 10 shows the South African excess to COVID-19 death ratio for the period from 2020/03/29 to 2020/10/01. Figure 10 shows that the excess to COVID-19 death ratio increased as the COVID-19 active cases in South Africa increased reaching a peak in the peak of the first COVID-19 epidemic wave and then decreasing thereafter. Figure 10 indicates that COVID-deaths were under-reported during the epidemic wave with the accuracy decreasing in the positive exponential phase of the epidemic wave.

**Figure 10:**
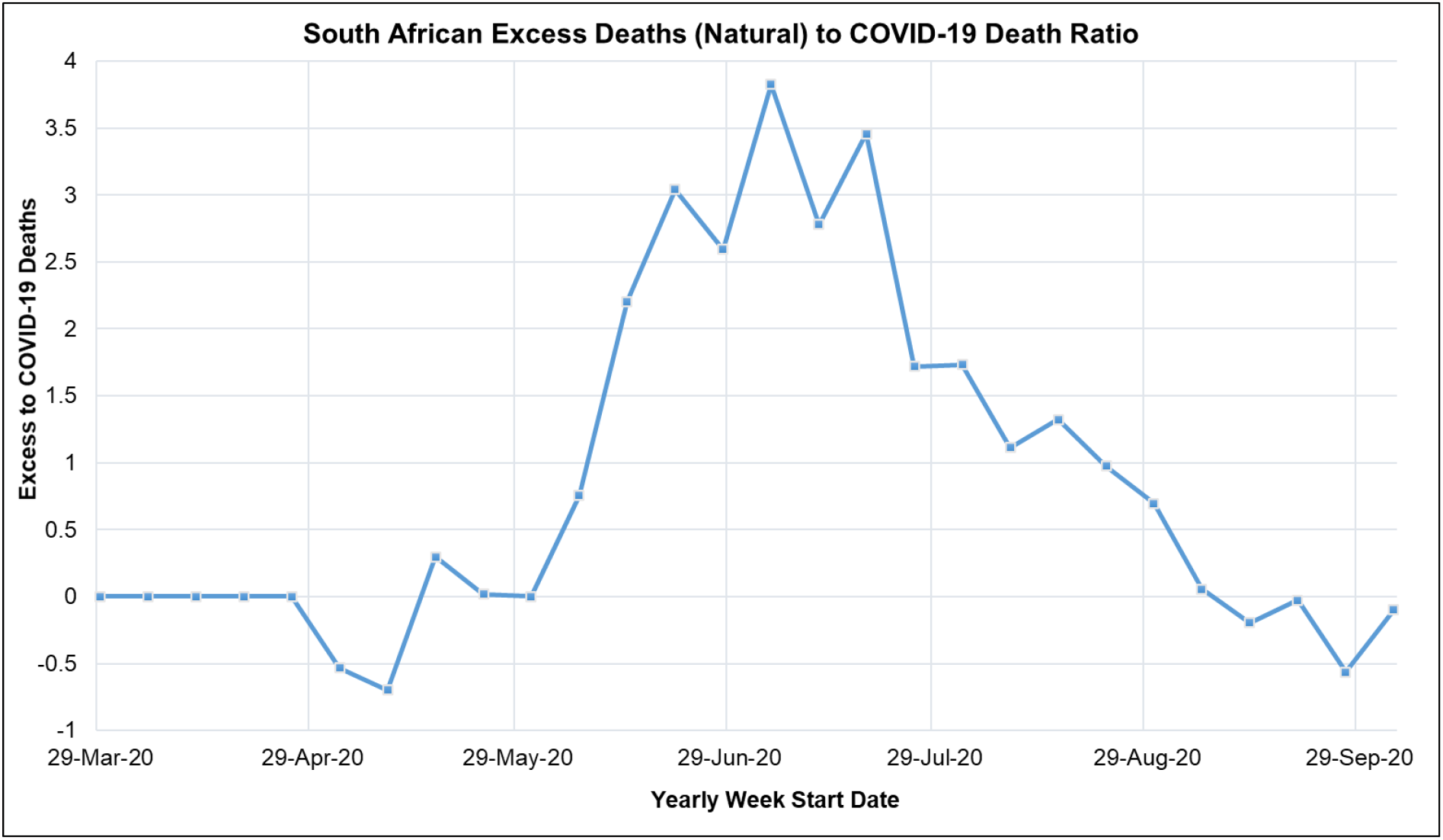
South African Excess to COVID-19 Death Ratio for the period from 2020/03/29 to 2020/10/01

### 3.6 Estimated COVID-19 Cases

Figure 11 shows the ARI COVID-19 SEIR model total COVID-19 cases in the South African first COVID-19 epidemic wave for the no lockdown, hard lockdown (National Lockdown Alert Level 5), moderate lockdown (National Lockdown Alert Level 4) and soft lockdown (National Lockdown Alert Level 3) scenarios. Figure 11 shows that if no COVID-19 NPI policies were implemented in South Africa, 25 391 522 COVID-19 active cases would have occurred at the peak of South Africa’s first COVID-19 epidemic wave. This corresponds to almost 42.6 % of the South African population. Figure 11 shows that if the National Lockdown Alert Level 5 policy had been continued for the duration of the South African fist COVID-19 epidemic wave, the peak active COVID-19 cases would have been reduced to 2 028 381 cases. While if the National Lockdown Alert Level 4 policy had been continued 4 621 066 peak active COVID-19 cases would have occurred. According to the ARI COVID-19 SEIR Model, as shown in Figure 11, the impact of the adjustment of NPI policies in South Africa up to the National Lockdown Alert Level 3 resulted in the peak active COVID-19 cases in South Africa being reduced to 8 057 754 cases. The results shown in Figure 11 indicate that the COVID-19 NPI policies implemented by the South African government in the form of national lockdown alert levels played a significant role in the reduction of active COVID-19 cases in South Africa.

**Figure 11:**
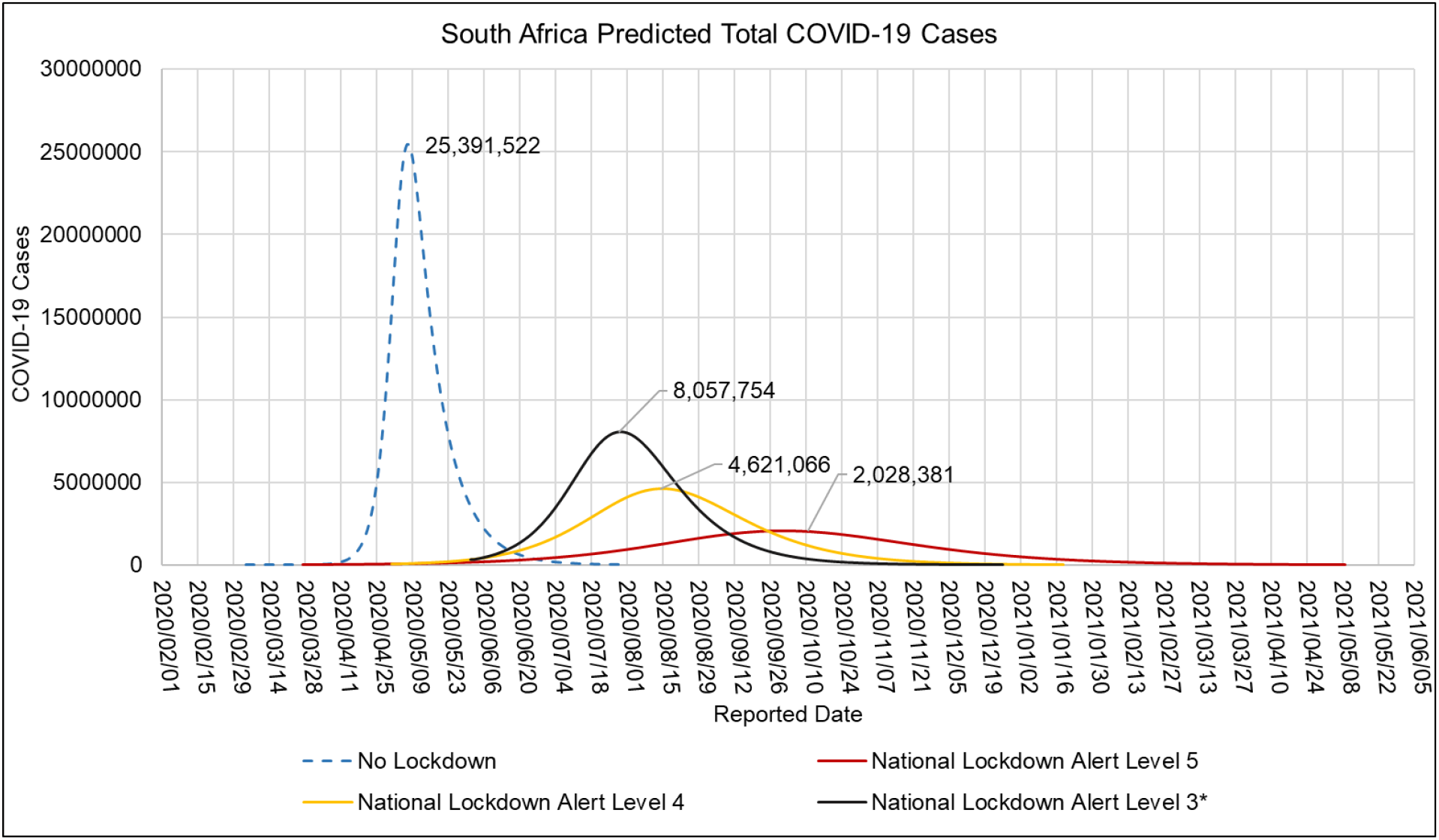
ARI COVID-19 SEIR Model Total COVID-19 Cases in the South African First COVID-19 Epidemic Wave for the No Lockdown, Hard Lockdown (National Lockdown Alert Level 5), Moderate Lockdown (National Lockdown Alert Level 4) and Soft Lockdown (National Lockdown Alert Level 3) scenarios

Figure 12 shows the ARI COVID-19 SEIR model symptomatic COVID-19 cases in the South African first COVID-19 epidemic wave for the no lockdown, hard lockdown (National Lockdown Alert Level 5), moderate lockdown (National Lockdown Alert Level 4) and soft lockdown (National Lockdown Alert Level 3) scenarios. Symptomatic COVID-19 cases are infectious individuals within the population of the model with either mild, moderate, severe or critical symptoms. Symptomatic COVID-19 cases have a high probability of being identified due to the awareness and visibility of COVID-19 symptoms or individuals seeking treatment. Figure 12 shows that if no COVID-19 NPI policies were implemented in South Africa, 1 964 568 peak symptomatic COVID-19 cases would have occurred in the peak of South Africa’s first COVID-19 epidemic wave. This value represents 7.74 % of the active COVID-19 cases. Implementation of the National Lockdown Alert Level 5 policy for the during of the South African first COVID-19 epidemic wave would have resulted in the symptomatic cases being reduced to 123 359 cases. While the National Lockdown Alert Level 4 would have resulted in the symptomatic cases being reduced to 280 325 cases. Figure 12 show that the impact of the adjustment of NPI policies in South Africa up to the National Lockdown Alert Level 3 resulted in the peak symptomatic COVID-19 cases in South Africa being reduced to 506 402 cases. The peak reported active COVID-19 cases in South Africa’s first COVID-19 epidemic wave was 173 587 cases as shown in Figure 5. The lower peak active COVID-19 cases observed relative to that reported in the ARI COVID-19 SEIR model indicates that a large number of symptomatic cases were not reported. With the model symptomatic COVID-19 cases being 2.91 times more than the observed peak active COVID-19 cases.

**Figure 12:**
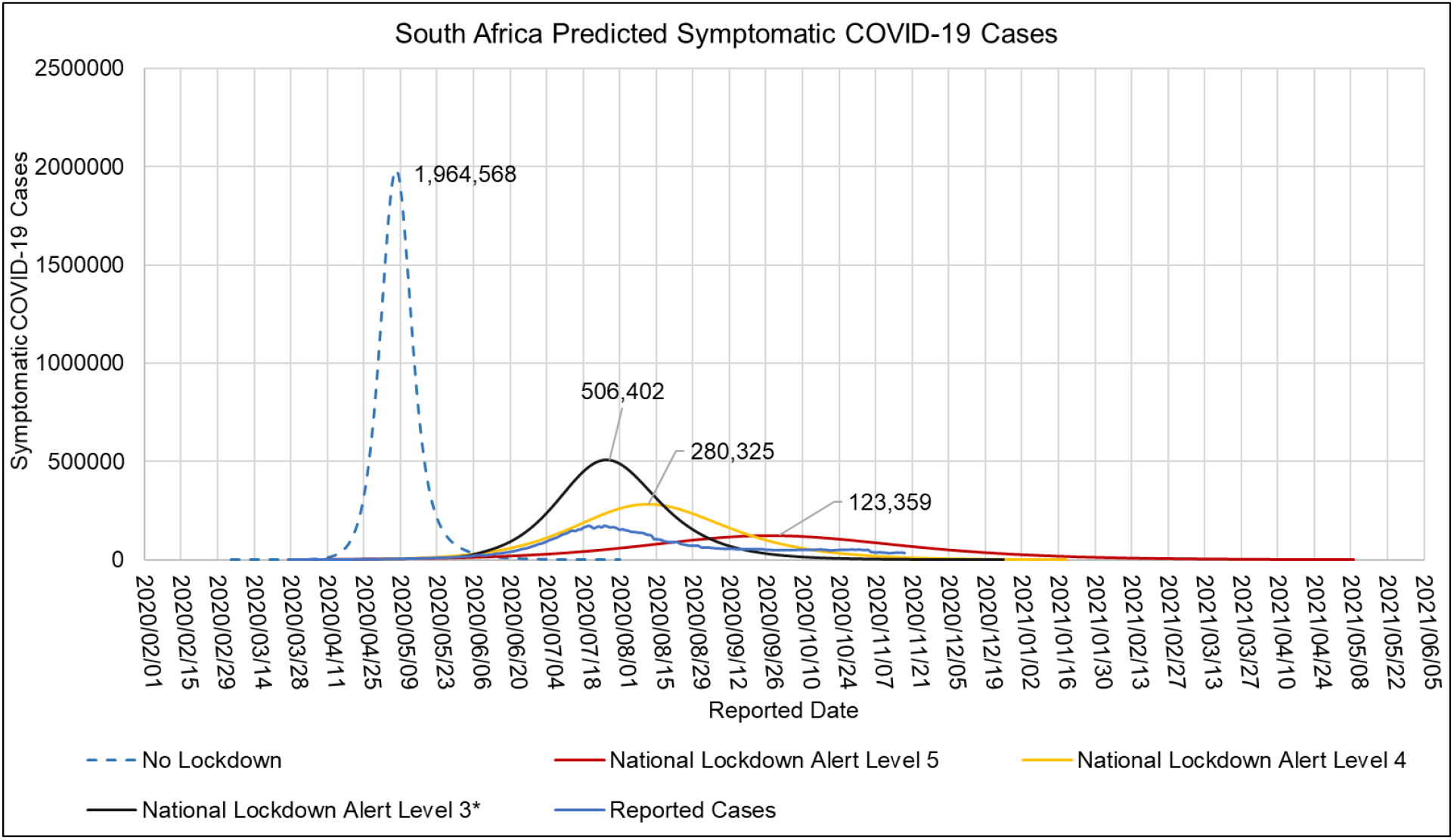
ARI COVID-19 SEIR Model Symptomatic COVID-19 Cases in the South African First Epidemic Wave for the No Lockdown, Hard Lockdown (National Lockdown Alert Level 5), Moderate Lockdown (National Lockdown Alert Level 4) and Soft Lockdown (National Lockdown Alert Level 3) scenarios

Table 14 shows the ARI COVID-19 SEIR model basic productive number, herd immunity, peak date, total infections, hospitalised cases and total deaths in the South African first COVID-19 epidemic wave for the no lockdown, hard lockdown (National Lockdown Alert Level 5), moderate lockdown (National Lockdown Alert Level 4) and Soft Lockdown (National Lockdown Alert Level 3) scenarios. Table 14 shows that if no COVID-19 NPI policies were implemented in South Africa, the initial basic reproductive number would have been 4.73 with estimated peak hospitalised cases at 100 653 and total deaths due to total COVID-19 deaths at 79 631. The total COVID-19 deaths estimated by the ARI COVID-19 SEIR model for the no national lockdown scenario is a conservative estimate. This estimate is based on the upper confidence intervals of the CFR shown in Table 8 which were derived from periods in which COVID-19 NPI policies were implemented. The impact of the higher number of active COVID-19 cases in the no lockdown scenario would have had an impact on the CFR. However, there was a limitation in determining the CFR via regression analysis in this period in South Africa due to the low number of reported COVID-19 and excess deaths. The CFR could have been adjusted using functions that relate active COVID-19 cases and the CFR however such functions are limited. This is due to lack of data as well as the complexity brought about by the multifactor that contribute towards the CFR.

**Table 14:**
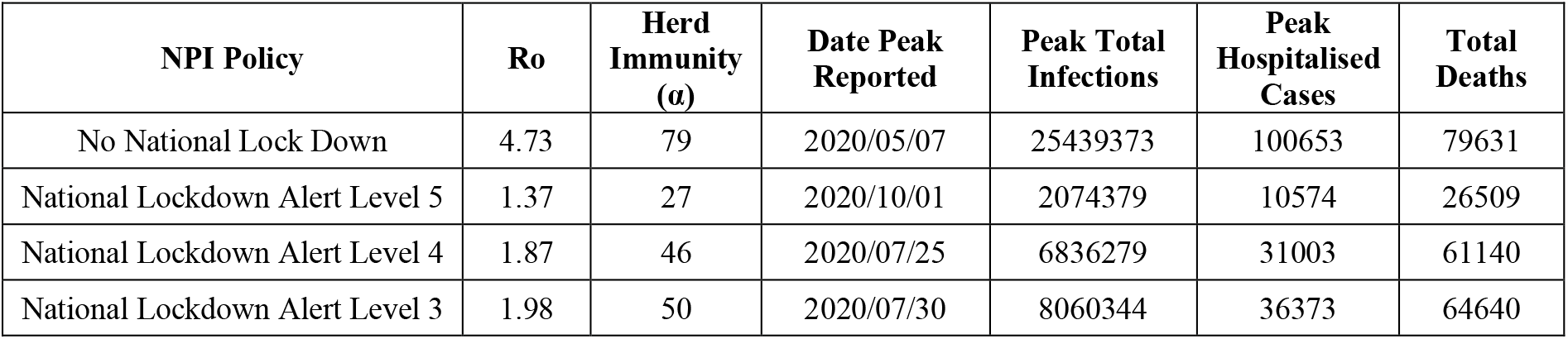
ARI COVID-19 SEIR Model Basic Reproductive Number, Herd Immunity, Peak Date, Total Infections, Hospitalised Cases and Total Deaths in the South African First COVID-19 Epidemic Wave for the No Lockdown, Hard Lockdown (National Lockdown Alert Level 5), Moderate Lockdown (National Lockdown Alert Level 4) and Soft Lockdown (National Lockdown Alert Level 3) scenarios

Table 14 shows that the implementation of the National Lockdown Alert Level 5 policy resulted in a decrease in the initial reproductive number by 71 % to 1.37. The decrease in the initial reproductive number can be attributed to the decrease in the daily effective contact number due to the NPI policy. If the National Lockdown Alert Level 5 policy was implemented for the duration of South Africa’s first COVID-19 epidemic wave, the peak total COVID-19 infections would have been reduced by 91.8 % to 2 074 379 cases. While peak COVID-19 hospitalised cases would have reduced by 89 % to 10 574 cases and total COVID-19 deaths by 67 % to 26 509 deaths. The peak of the first epidemic wave would have been delayed by 147 days to the 1^st^ of October 2020. Implementation of the National Lockdown Alert Level 4 resulted in an initial reproductive number of 1.87. If the National Lockdown Alert Level 4 policy was implemented for the duration of South Africa’s first COVID-19 epidemic wave, the peak total COVID-19 infections would have been reduced by 73.1 % to 6 836 279 cases. While peak COVID-19 hospitalised cases would have been reduced by 69 % to 31 003 cases and total COVID-19 deaths to 61 140 deaths. The peak of the first epidemic wave would have been delayed by 79 days to the 25^th^ of July 2020. Implementation of the National Lockdown Alert Level 3 resulted in an initial reproductive number of 1.98. The adjustment to the National Lockdown Alert Level 3 policy resulted in a reduction in the peak total COVID-19 infections by 68.3 % to 8 060 344 cases. While peak COVID-19 hospitalised cases were reduced by 69 % to 31 003 cases and total COVID-19 deaths to 64 640 deaths. The peak of the first epidemic wave was delayed by 84 days to the 30^th^ of July 2020. Table 14 shows that if no COVID-19 NPI policies were implemented in South Africa, the COVID-19 herd immunity required in South Africa would be 79 %. Table 14 shows that implementation of NPI policies results in a decrease in the required herd immunity with the condition that the NPI policy is maintained.

### 3.7 Estimated COVID-19 Effective Reproductive Number

Figure 13 shows the ARI COVID-19 SEIR Model effective reproductive number in the South African COVID-19 first epidemic wave for the no Lockdown, hard Lockdown (National Lockdown Alert Level 5), moderate Lockdown (National Lockdown Alert Level 4) and soft Lockdown (National Lockdown Alert Level 3) scenarios. Figure 13 shows that for the implemented NPI policies in South Africa up to the National Lockdown Alert Level 3 the effective reproductive number was between 1.98 to 0.40 in the first COVID epidemic wave. According to the NICD, the nationally average reproductive number during the period of the National Lockdown Alert Level 5 was 1.29 (95%CI: 1.9601.58)) and rose to 1.5 by end of April. While the average reproductive number in National Lockdown Alert 3 was 1.05 (95%CI:1.01-1.09) between 1 June and 1 August dropping below 1 during the last week of July [90]. These results are similar to what was obtained by the ARI COVID-19 SEIR Model as shown in Figure 13.

**Figure 13:**
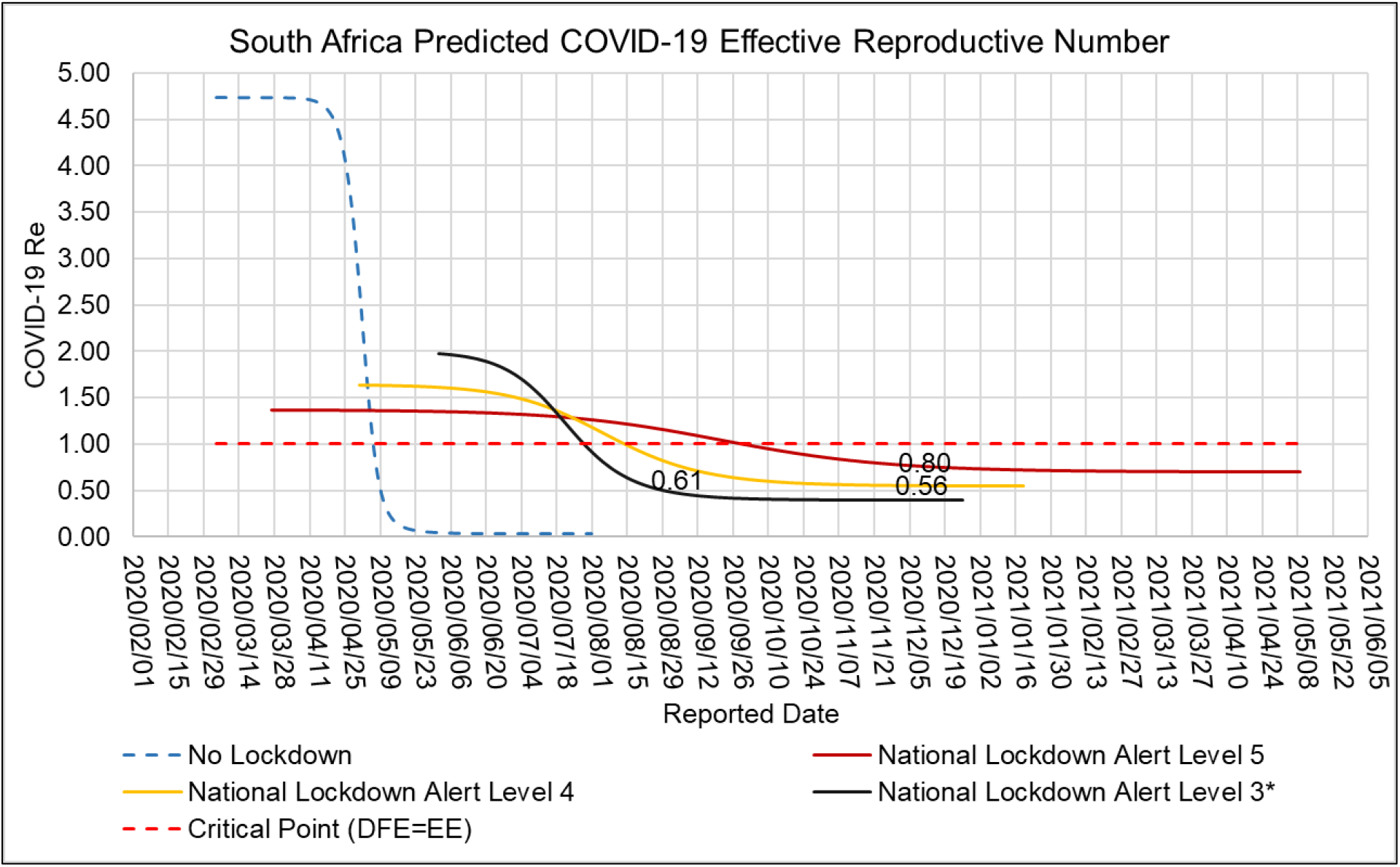
ARI COVID-19 SEIR Model Effective Reproductive Number in the South African COVID-19 First Epidemic Wave for the No Lockdown, Hard Lockdown (National Lockdown Alert Level 5), Moderate Lockdown (National Lockdown Alert Level 4) and Soft Lockdown (National Lockdown Alert Level 3) scenarios

### 3.8 Impact of NPIs on hospitalised COVID-19 cases, COVID-19 deaths and healthcare system preparedness

Figure 14 shows the ARI COVID-19 SEIR Model admission status in the South African first COVID-19 epidemic wave. The admission status was calculated based on the admission status means in Table 9 developed from the NICD DATCOV surveillance system data. Figure 14 shows that the ARI COVID-19 SEIR Model estimated that there would be 26 279, 5 847, 5 630, 2 947, 2 719, 1 334 COVID-19 patients in the general ward, intensive care unit, on oxygen, in high care, on the ventilator, in the isolation ward in South African hospitals. The estimated peak for hospital cases in South Africa’s first COVID-19 epidemic wave by the ARI COVID-19 SEIR Model was the 6^th^ of August 2020. Figure 15 shows the ARI COVID-19 SEIR Model ICU Bed and Ventilator Occupied Capacity in South Africa’s first COVID-19 epidemic wave. According to Houreld et al., 2020 there were 3 200 ventilators and 3 300 ICU beds in South Africa by May 2020. Based on these estimates of the available ventilators and ICU beds in South Africa at the time, the ICU occupied capacity in South Africa was breached by the 17^th^ of July 2020 with the occupied capacity at the peak at 167 % as shown in Figure 15. While the ventilator capacity was not breached. Figure 15 shows that at the peak the occupied capacity was 77 % indicating that South Africa had sufficient ventilators and insufficient ICU beds in the first COVID-19 epidemic wave. The results obtained in this section indicate some level of preparedness by the South African health care system for the first COVID-19 epidemic wave. However, the distribution and management of these resources is an important factor that needs to be assessed to develop adequate conclusions regarding the preparedness of the South African health care system in the first COVID-19 epidemic wave.

**Figure 14:**
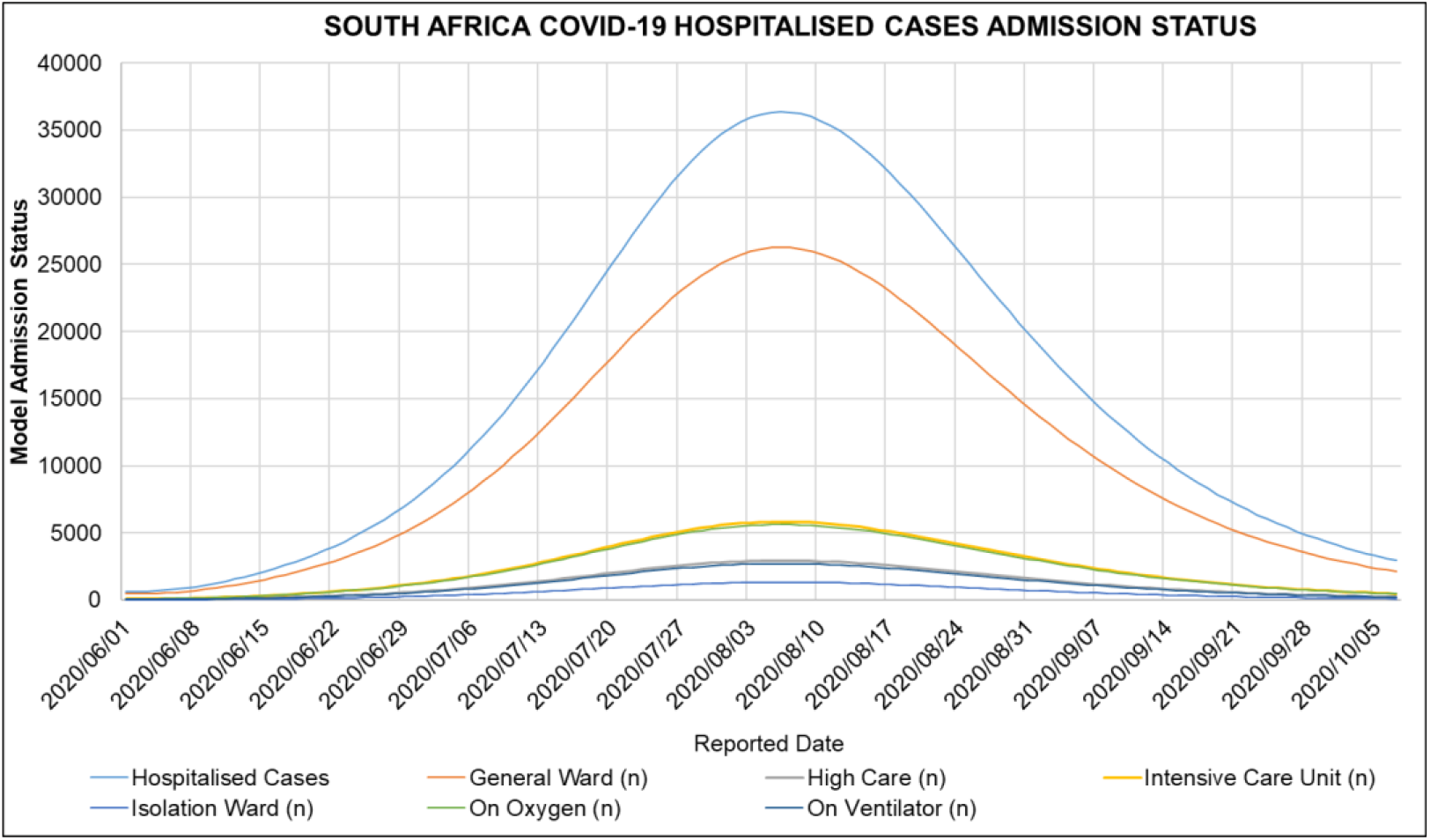
ARI COVID-19 SEIR Model Admission Status in the South African First COVID-19 Epidemic Wave

**Figure 15:**
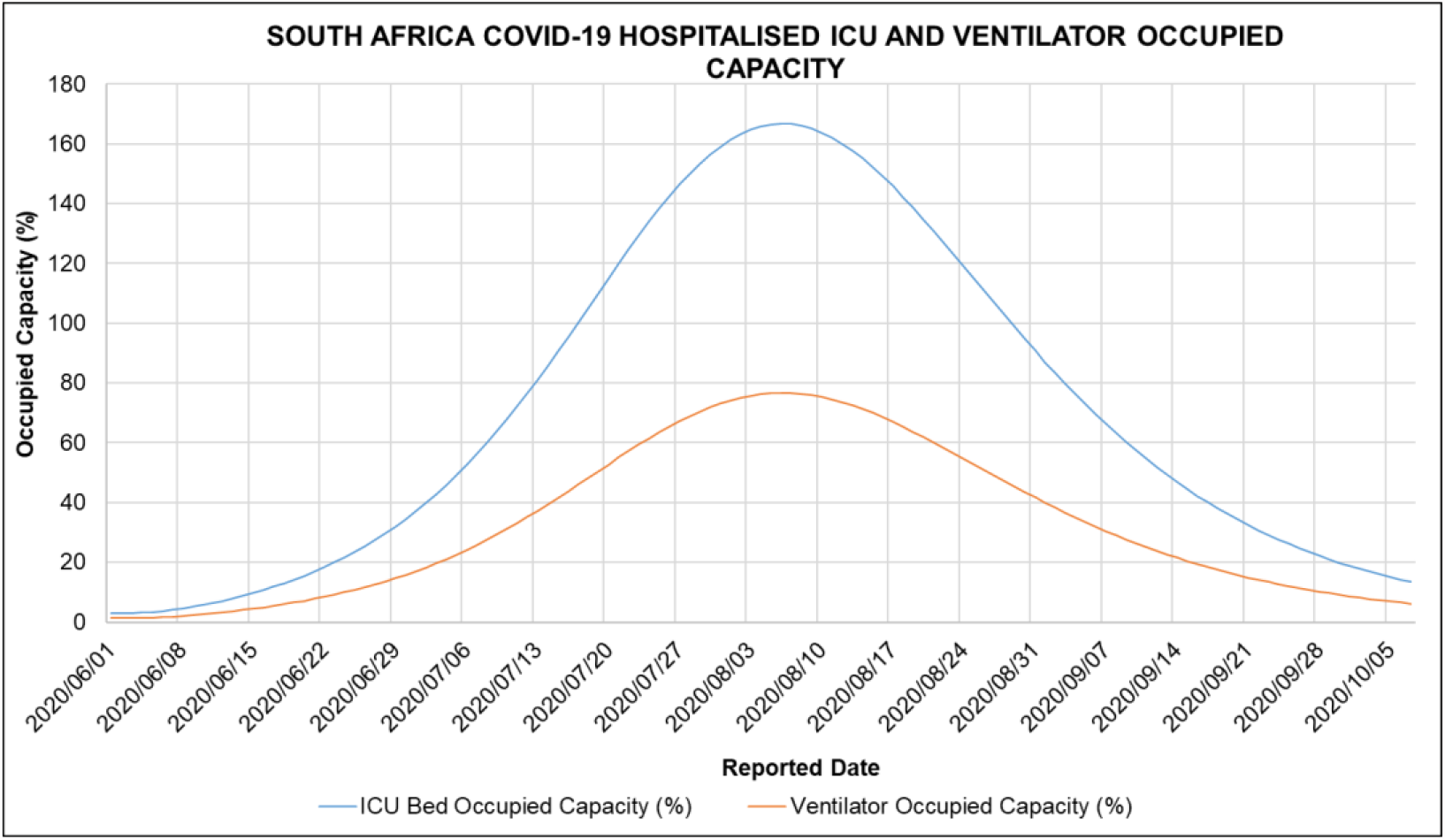
ARI COVID-19 SEIR Model ICU Bed and Ventilator Occupied Capacity in the South African First COVID-19 Epidemic Wave

### 3.9 Review of South Africa COVID-19 Modelling

COVID-19 has been widely modelled with variations of the SEIR model [26–28,30–32,92]. In this review, we explore COVID-19 models which had a significant influence on South Africa’s policy response to COVID-19 and the ARI COVID-19 Model. We also explore the models’ limitations and success.

One of the earliest COVID-19 transmission models to be published was the Imperial College London COVID-19 Model [28,33]. The Imperial College London COVID-19 Model had a great influence in the early policy response to COVID-19 in many countries including Africa [28]. The Imperial College London Model explored the use of Non-Pharmaceutical Interventions (NPI) in suppressing or mitigating COVID-19 using a mathematical transmission model. Ethical and economic factors were not explored in this model. The NPIs considered in this model were home isolation and quarantine, social distancing and closure of schools and universities. Interventions were modelled to reduce the effective contact rates thereby reducing the transmission of COVID-19. The transmission was explored in households, workplace, school, or random community. Mitigation strategies explored was the reduction of the COVID-19 Infections and protection of high-risk groups to exposure to COVID-19 whilst suppression explored the reduction of the Basic Reproductive Number (R_0_) to less than the critical disease-free equilibrium point (Less than 1). The model estimated that NPIs if implemented would result in a reduction in Health Care COVID-19 cases by two thirds and COVID-19 deaths by half. Whilst without interventions critical care beds would be exceeded over 30 times compared to capacity (in Great Britain and the United States of America). The NPIs would have to be maintained until the availability of a vaccine to immunise the population. If NPIs were not maintained, it was suggested a potential rebound of transmission could occur with an epidemic comparable scale to that of no interventions. Population-wide social distancing was observed to have the largest impact in suppressing COVID-19 whilst stopping mass gathering was predicted to have little impact because of the short contact time relative to household settings. The model explored pre-symptomatic infectiousness (12 hours prior symptoms), assumed that two-thirds of cases are symptomatic, 30 % of hospitalised cases will require critical care whilst 50 % of critical care cases will die [28]. In retrospect, the Imperial College London COVID-19 model did well to quantify the magnitude of the impact of NPIs in COVID-19 mitigation. The prediction of “Second Waves” of the COVID-19 pandemic after relaxation/lifting of some of the NPIs particularly movement restrictions were observed in Europe (August /September 2020) [93] and in Africa (November/December 2020) [94]. Although the initial Imperial College London COVID-19 model was successful in understanding the impact of NPIs on the COVID-19 pandemic, it overestimated the severity of COVID-19 [95] and was subsequently revised [96]. Asymptomatic cases of COVID-19 have been observed to account for more than 33 % [47–49]. Also, in retrospect, in adapting such a model to Africa’s policy response, the model did not consider the risk factor of disease comorbidity to COVID-19 severity particularly in Africa where there is a high disease burden. The model did not account for potential cases which are not hospitalised and the impact of the COVID-19 pandemic on Excess Natural Deaths in Africa.

Another COVID-19 Model of note was the model produced by the Center for Disease Dynamics, Economics & Policy (CDDEP) [27,29]. The CDDEP COVID-19 Models tried to understand the impact of Country-Wise Lockdowns [29] and the Health Care system preparedness of African countries [27]. The CDDEP COVID-19 Model had a more revised severity of COVID-19 particularly in the proportion of asymptomatic cases and the severe case fatality rate relative to the Imperial College London COVID-19 Model [29]. The CDDEP COVID-19 Model also attempted to account for the rate progression of COVID-19 due to Age, TB, and HIV/AIDS. The CDDEP COVID-19 Model predicted that 31 of 50 African countries will not have enough hospitals beds and even if 30 % of severely infected patients seek health services only 34 of 48 African countries have enough ICU Beds. Only five countries (Carbo Verde, Gabon, Egypt, and South Africa) would have enough ventilators. The CDDEP COVID-19 Model predicted the delay in peak due to lockdown measures and that implementation of large-scale mitigation measures may not be feasible or sustainable in Low and Middle-Income Countries (LMICs) in Africa [27]. The influence of COVID-19 Modelling on Southern Africa’s policy response to COVID-19 remains under-reported and there are limited published National COVID-19 Models with exception of South Africa. The Africa Center for Disease Control (Africa CDC) in response to the COVID-19 pandemic created a COVID-19 Modelling group in a bid to try to foster collaboration and sharing of information within COVID-19 modellers in Africa.

South Africa received much attention with regards to COVID-19 Modelling with several models being published and noted by the South African government [34]. Of note, is the National COVID-19 Epi Model (NCEM) and the National COVID-19 Cost Model (NCCM) by the South African COVID-19 Modelling Consortium, 2020. The NCEM is an SEIR stochastic compartmental transmission model that was developed to estimate the total and reported incidence of COVID-19 cases in South Africa up to November 2020. While the NCCM was a model developed to determine the COVID-19 response budget in South Africa. The NCEM and NCCM played a key role in South Africa’s policy and planning response to COVID-19. The NCEM assumed a relatively high proportion of asymptomatic cases (75 %) and symptomatic cases (95 %) and modelled an optimistic and pessimistic scenario. In the optimistic scenario, a Hard lockdown measure reduced COVID-19 transmissions by 60 %, Moderate Lockdown by 35 % and social distancing by 20 %. In the pessimistic scenario, a Hard lockdown measure reduced COVID-19 transmissions by 40 %, Moderate Lockdown by 25 % and social distancing by 10 %. The NCEM anticipated that lockdowns would flatten the epidemic curve and delay the COVID-19 peak in South Africa by 2 to 3 months. South Africa would observe peak demand for hospital care between August and September 2020. These factors were dependent on the response of the population’s social behaviour to measures. The NCCM estimated a total budget of 26 to 32 Billion Rands would be required for COVID-19 response in South Africa. The budget would cover Personal Protective Equipment (PPE), additional ICU, hospital beds and staff, additional PHC staff, ventilators, drugs, isolation facilities, testing and surveillance and port health budgets. The NCEM did not account for disease risk factors and location transmission risks rather assuming random mixing at provincial levels. For age-related risks, the NCEM used population-adjusted age-specific mortalities from the Chinese epidemic. The NCEM model did well in predicting the COVID-19 epidemic in South Africa. Particularly the expected peak (magnitude and progression). The NCEM took account and highlighted the impact of the COVID-19 epidemic at a provincial level with provincial variability noted in the difference in seeding and community contact behaviour. The NCEM also constantly revised its parameters ensuring more accurate modelling as the COVID-19 Epidemic in South Africa progressed [97].

Most COVID-19 Models have been proactive in attempting to predict and quantify the epidemic in Africa to advise on Africa’s early policy response to the epidemic. The ARI COVID-19 Model can be considered a semi-reactive model. The ARI COVID-19 Model was developed well within the epidemic in South Africa (July 2020). The model uses regression and sensitivity analysis of South African COVID-19 Reported Cases (Before Lockdown Measures), COVID-19 Deaths and Excess Natural Deaths (During Lockdown Measures) to quantify the impact of implemented NPIs in the suppression of COVID-19 in South Africa. This was done by adjusting the effective contact number during the regression analysis at the different National Lockdown Alert Levels (5, 4, 3) implemented in South Africa and seeding the model with observed COVID-19 Deaths (accounting for Excess Natural Deaths).**Error! Reference source not found**. Table 15 shows the Imperial College London COVID-19 Model (Ro=3.3, Mitigation-Social distancing the whole population and Suppression-1.6 deaths per 100,000 per week trigger), NCEM (Optimistic Scenario), ARI COVID-19 Model (With NPI interventions), CDDEP COVID-19 Model (Moderate Lockdown), observed total infections, active infections, total deaths, peak hospitalised cases, peak date with standard Deviation (StDev) and co-efficient of variance (CoV) of model outputs. The NCEM, CDDEP and ARI COVID-19 Models accurately predicted the dates of the first COVID-19 epidemic peak in South Africa. Table 15 shows that the predicted total infections, peak active infections, total deaths, peak hospitalised cases in the reviewed models had a standard deviation of 4 865 693, 2 362 685 cases, 48 303 deaths, 25780 cases with a coefficient of variance of 12.9 %, 102.7 %, 118.4 %, 84.2 % respectively. The relatively high coefficient of variance in model output or results shows the sensitivity in the model parameters used in modelling considering different model parameters were used in the reviewed models. The relatively high coefficient of variance also shows the uncertainty in the accuracy of the reviewed COVID-19 models. These results suggest the need for sensitivity analysis on model parameters when modelling COVID-19 using transmission models.

**Table 15:** Imperial College London COVID-19 Model (Ro=3.3, Mitigation-Social distancing the whole population and Suppression-1.6 deaths per 100,000 per week trigger), NCEM (Optimistic Scenario), ARI COVID-19 Model (With NPI interventions), CDDEP COVID-19 Model (Moderate Lockdown), Observed Total Infections, Active Infections, Total Deaths, Peak Hospitalised Cases, Peak Date with Standard Deviation (StDev) and Co-efficient of Variance (CoV) of Model Outputs

| Model Output | Imperial College London COVID-19 Model | NCEM | ARI COVID-19 Model | CDDEP COVID-19 Model | StDev | CoV (%) | Observed Peak Cases/Excess Deaths |
| --- | --- | --- | --- | --- | --- | --- | --- |
| Predicted Total Infections | 37762240 | 48658190 | 47393046 |  | 4865693 | 12.9 |  |
| Predicted Peak Active Infections |  | 4696334 | 8060344 | 2300000 | 2362685 | 102.7 | 173590 |
| Predicted Total Deaths | 153073 | 40784 | 64640 |  | 48303 | 118.4 |  |
| Predicted Peak Hospitalised Cases | 30600 | 93006 | 36373 | 34000 | 25780 | 84.2 | 47744 |
| Peak Date |  | 8/1/2020 | 7/30/2020 | 8/1/2020 | 1 | 0.0021 | 7/20/2020 |

From all models reviewed, the predicted peak of active cases was more than 27 to 46 times than the observed/reported. This observation highlights the need for more rigorous testing in South Africa especially with most COVID-19 cases estimated to be asymptomatic. Improvement in the testing protocol (inclusion of serological testing) is also needed to avoid false positives. A case in point in this argument is the notion that the epidemic in South Africa has not progressed to the extent to which the models have predicted. Factors to consider in this argument are the following:

1. COVID-19 Models conduct scenario analysis for the duration of the entire epidemic. Therefore, changes in NPIs during the epidemic cause a difference between model and observed results. The ARI COVID-19 Model as a semi-reactive model tried to account for changes in the NPIs used by Southern African governments and still obtained results that correlate with the pro-active models. Therefore, this point may not be valid in the argument.
2. COVID-19 Models assumed homogenous or random mixing. All models made this assumption. With a note of the NCEM which attempted to understand seeding and contact rates at a provincial level still assuming random mixing in provinces. The COVID-19 Epidemic in South Africa seems to be occurring in pockets within the population as clusters of cases [98]. South Africa has a high rural population (low density) with the urban population agglomerated in slums (high density). With the epidemic in most countries seeding in agglomerated cities, the rate of contacts from high density to low-density areas in the presence of lockdowns influences the rate of transmission. Clustering of the cases within the population suggests that populations are not homogenously mixing as assumed especially in the presence of NPIs. This could mean models are overestimating contact rates and the progression (magnitude) of the epidemic.

Implementation of NPIs results in the lowering of the required herd immunity to reach the Disease-Free Equilibrium. Therefore, once NPIs particularly movement restrictions/lockdowns are lifted there is likely to be a secondary wave, infecting the susceptible from the primary wave. COVID-19 models have been depicting the epidemic as a single occurrence, the implementation/relaxation/removal of NPIs can result in negative or positive damping of the epidemic curve resulting in the epidemic occurring in a series of waves as opposed to a single occurrence.

## 4. Conclusions

The following NPIs can be drawn from the South African National Lockdown Alert Level policy response implemented in the period of the country’s first COVID-19 epidemic wave: Entry and exit screening at borders, limitations of movements and gatherings, closure/limitations of the institution and business activities, ban/limiting of alcohol and tobacco industries, isolation, quarantine of potentially infected persons and contact tracing protocols, use of PPE and hygienic protocols. The COVID-19 policy response implemented in South Africa indicated a focus on restricting contact between individuals within the population. The South African COVID-19 NPI policies were effective in reducing the movement of communities in retail and recreation, grocery and pharmacy, parks, transit stations, workplaces locations in South Africa. The general trend in population movement in these locations shows that the National Lockdown Alert Level 5,4,3,2 were approximately 30% more effective in reducing population movement concerning each increase by 1 Alert Level. While the National Lockdown Alert Level 1 and 2 had a similar impact on the population movements in the South African communities. The effective SARS-CoV-2 daily contact number (β) in South Africa was estimated to be 0.498, 0144, 0.196, 0.208 day^-1^ for the no lockdown, National Lockdown Alert Level 5, 4 and 3 model scenarios respectively. These results translate into a reduction of 71.1 %, 60.6 %, 58.1 % in the effective SARS-CoV-2 daily contact number from having no lockdown in South Africa to the implementation of the National Lockdown Alert Level 5, 4 and 3 respectively. While adjusting the Alert Level by 1 was approximately 30 % more effective in reducing population movement in South Africa. The translated reduction in the effective SARS-CoV-2 daily contact number (β) was only 4.13 % to 14.6 % concerning increasing Alert Levels. Covid-19 and the policies formulated to help with reducing the spread of the SARS-CoV-2 had adverse effects on the South African economy, more especially on Micro Small and Medium Enterprises (MSMEs) and informal workers and their households.

The National Health Laboratory Service (NHLS) and National Institute for Communicable Diseases (NICD) were the national laboratories conducting the COVID-19 testing in South Africa and for other Southern African countries such as Lesotho, Namibia and Eswatini in their earlier COVID-19 epidemics. Major private laboratories involved in COVID-19 testing in South Africa included Abbott, Ampath, Pathcare and Lancet Laboratories. There were 5 383 078 cumulative COVID-19 tests conducted in South Africa in the period reported in 2020/02/07 to 2020/10/01 with an average daily testing capacity of 18 069±13760 COVID-19 tests. The correlation of COVID-19 testing and cases shown in this study indicates that testing has an impact on how the epidemic is observed/reported. Due to limitations in COVID-19 testing in South Africa, there is a probability that some COVID-19 cases in South Africa were not reported. There were 676 084 cumulative, 609 584 recovered COVID-19 cases and 16 866 COVID-19 deaths reported in the period of 2020/01/22 to 2020/10/01. 90.2 % of the COVID-19 cases reported in the respective reported period recovered. The first COVID-19 epidemic wave in South Africa lasted for 205 days from the first reported case. The total peak COVID-19 active cases in South Africa’s first COVID-19 epidemic wave was 173 587 COVID-19 cases observed on the 26^th^ of July 2020. The National Lockdowns Alert Level 5 and 4 were implemented at the start of the epidemic however the COVID-19 policy in South Africa was then eased to Lockdown Alert Level 3 where the majority of the positive exponential phase of the first epidemic wave was observed. The COVID-19 policy was further eased to the National Lockdown Alert Level 2 and 1 in the negative exponential phase of the first epidemic wave. District and provincial confinement of the South African population due to the National Alert Level Lockdowns, the difference in testing capacity, population, population distribution, residential settings and business activities in South African provinces resulted in different first COVID-19 epidemic amplitudes and periods being observed in provinces. At least 101 introductions of SARS-CoV-2 were estimated in South Africa, with the bulk of the important introductions occurring before lockdown from Europe. There were 42 different SARS-CoV-2 lineages with 16 South African specific lineages assigned to South African genomes in the period of 2019/12/24-2020/08/26. A review done in this study indicates that the SARS-CoV-2 C.1, B.1.1.54, B.1.1.56 lineage clusters were major drives of the first COVID-19 epidemic wave in South Africa.

During South Africa’s first COVID-19 epidemic wave, the estimated mean for the COVID-19 general ward, intensive care unit, on oxygen, high care, on ventilator, in isolation ward admission status in South African hospital was 58.5 %, 95% CI [58.1,59.0], 13.4 %, 95% CI [13.1,13.7], 13.3 %, 95% CI [12.6,14.0], 6.37 %, 95 % CI [6.23,6.51], 6.29 %, 95% CI [6.02,6.55], 2.13 %, 95% CI [1.87,2.43] respectively. Most COVID-19 patients reporting to South African hospitals were admitted into the general wards. The proportion reporting to intensive care units and on oxygen were similar regarding confidence intervals. A relatively low proportion of patients were admitted to the isolation ward. The estimated mean South African COVID-19 patient discharge rate was 11.9 days per patient. While the estimated mean of the South African COVID-19 patient case fatality rate (CFR) in hospital and outside the hospital was 2.06 %, 95% CI [1.86,2.25] (deaths per admitted patients) and 2.30 %, 95% CI [1.12,3.83](deaths per severe and critical cases) respectively. Linear regression analysis determined that the CFR and discharge rate in South African hospitals was constant in the first COVID-19 epidemic wave in South Africa. This indicates good clinical management in the face of adversity where the increasing number of cases towards the peak of the first epidemic wave did not influence the hospital COVID-19 discharge and death rate. The COVID-19 CFR outside the hospital was observed to be higher than in the hospital in the first COVID-19 epidemic wave in South Africa. People in the age groups over 40 years accounted for 78.9 % of the COVID-19 hospitalised cases in South African hospitals. While children under 19 years only accounted for 4.07 % of the COVID-19 hospitalised cases. Indicating low case incident in the severe and critical COVID-19 disease in children in South Africa.

COVID-19 deaths in South African hospitals increased with an increase in age groups up to the age group 60 to 69 years. COVID-19 deaths of children in South African hospitals was relatively low. People in the age groups of 50 to 59 years and 60 to 69 years had the highest proportion of COVID-19 deaths in South African hospitals accounting for 50.3 % of the hospitalised deaths. People in the age groups over 80 years had the highest risk of dying from COVID-19 in South African hospitals with a cumulative COVID-19 death risk ratio of 23.7, 95% CI [22.6,25.1]. The risk of dying from COVID-19 in South African hospitals for age groups over 20 years approximately doubled with an increase in age of 10 years. The weekly excess (natural) deaths, excess (natural) to natural deaths (%), excess deaths (natural) to COVID-19 death ratio for the period from 2019/12/29 to 2020/10/01 was estimated to be 2114, 95% CI [1239,2990], 16.7 %, 95% CI [10.9,22.5] and 1.12, 95% CI [0.55,1.68] respectively. COVID-deaths were under-reported during the epidemic wave with the accuracy decreasing in the positive exponential phase of the epidemic wave. The relatively low value of the excess (natural) to natural deaths in South Africa shows that the COVID-19 disease or epidemic did not account for the majority of the natural deaths occurring in South Africa in the respective period reported.

If no COVID-19 NPI policies were implemented in South Africa, 25 391 522 COVID-19 active cases would have occurred at the peak of South Africa’s first COVID-19 epidemic wave. This corresponds to almost 42.6 % of the South African population. The initial basic reproductive number would have been 4.73 with an estimated peak hospitalised cases at 100 653 and total deaths due to total COVID-19 deaths at 79 631 (conservative estimate). The adjustment to the National Lockdown Alert Level 3 policy resulted in a reduction in the peak total COVID-19 infections by 68.3 % to 8 060 344 cases. While peak COVID-19 hospitalised cases were reduced by 69 % to 31 003 cases and total COVID-19 deaths to 64 640 deaths. The peak of the first epidemic wave was delayed by 84 days to the 30th of July 2020. The estimated required herd immunity in South Africa’s first COVID-19 epidemic wave was 79 %. While the estimated, effective reproductive number in the first COVID epidemic wave. for the implemented NPI policies in South Africa up to the National, Lockdown Alert Level 3 was between 1.98 to 0.40. This study estimates that the ICU occupied capacity in South Africa was breached by the 17^th^ of July 2020 with the occupied capacity at the first COVID-19 epidemic peak at 167 %. While the ventilator capacity was not breached at 77 %. South Africa had sufficient ventilators and insufficient ICU beds in the first COVID-19 epidemic wave. The results obtained in this study indicate some level of preparedness by the South African health care system for the first COVID-19 epidemic wave. However, the distribution and management of these healthcare resources is an important factor that needs to be assessed to develop adequate conclusions regarding the preparedness of the South African health care system in the first COVID-19 epidemic wave. From all models reviewed, the predicted peak of active cases was more than 27 to 46 times than the observed/reported indicating the underreporting and need for more rigorous COVID-19 testing in South Africa.

The COVID-19 NPI policies implemented by the Government of South Africa in the form of national lockdown alert levels played a significant role in the reduction of COVID-19 active, hospitalised cases and deaths in South Africa’s first COVID-19 epidemic wave.

## Supporting information

Excess Deaths_South Africa

Community_Mobility_South Africa

Model_Review_South Africa

Model_Results_South Africa

NPIs_South Africa

Regression Analysis_South Africa

Reported Cases_Provincial_South Africa

Reported & Hospitalised Cases_South Africa

Testing_South Africa

## Data Availability

The data in this manuscript can be requested from the The Afrikan Research Initiative via the. All data is available and free.

https://www.afrikanresearchinitiative.com/

## 5. Acknowledgements

ARI would like to thank the members of the ADDRG and ACMRG for their voluntary commitment to working in the ARI COVID-19 Research Project. We would like to thank Professor Tom Moultrie and Dr Sheetal P. Silal from the University of Cape Town for providing consultation in the early modelling phase of the ARI COVID-19 Research project. We acknowledge the work by the National Institute for Communicable Diseases (NICD), Western Cape Department of Health Provincial Health Data Centre (PHDC) and South African Medical Research Council (SAMRC) in which the ARI COVID-19 Projects draws a lot of its data from. Lastly, we want to salute the scientific community, governments, health care workers, essential personnel in their response to the pandemic and we pay homage to those who have lost their lives due to the COVID-19 pandemic.

## 6. Funding and Conlfict of interests

This study falls under the ARI COVID-19 Research Project of which the project is currently not funded. Data used in this study was obtained from public sources.

## 8. Appendix

**Figure A. 3:**
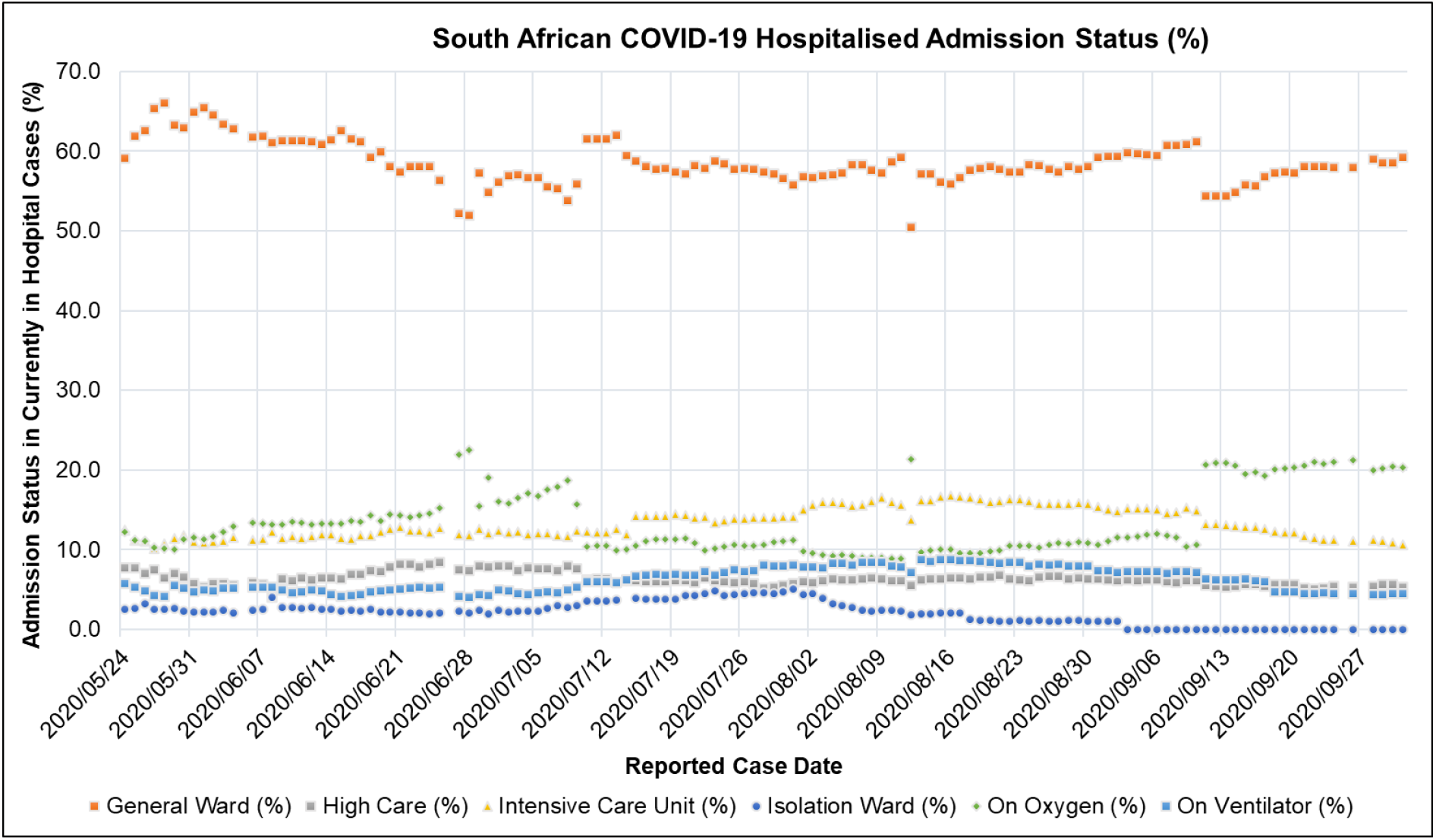
South African COVID-19 Hospitalised Admission Status (%): Currently in Hospital, General Ward, High Care, Intensive Care Unit, Isolation Ward, On Oxygen, On Ventilator for the period 2020/05/24 to 2020/10/01 [41]

**Figure A. 4:**
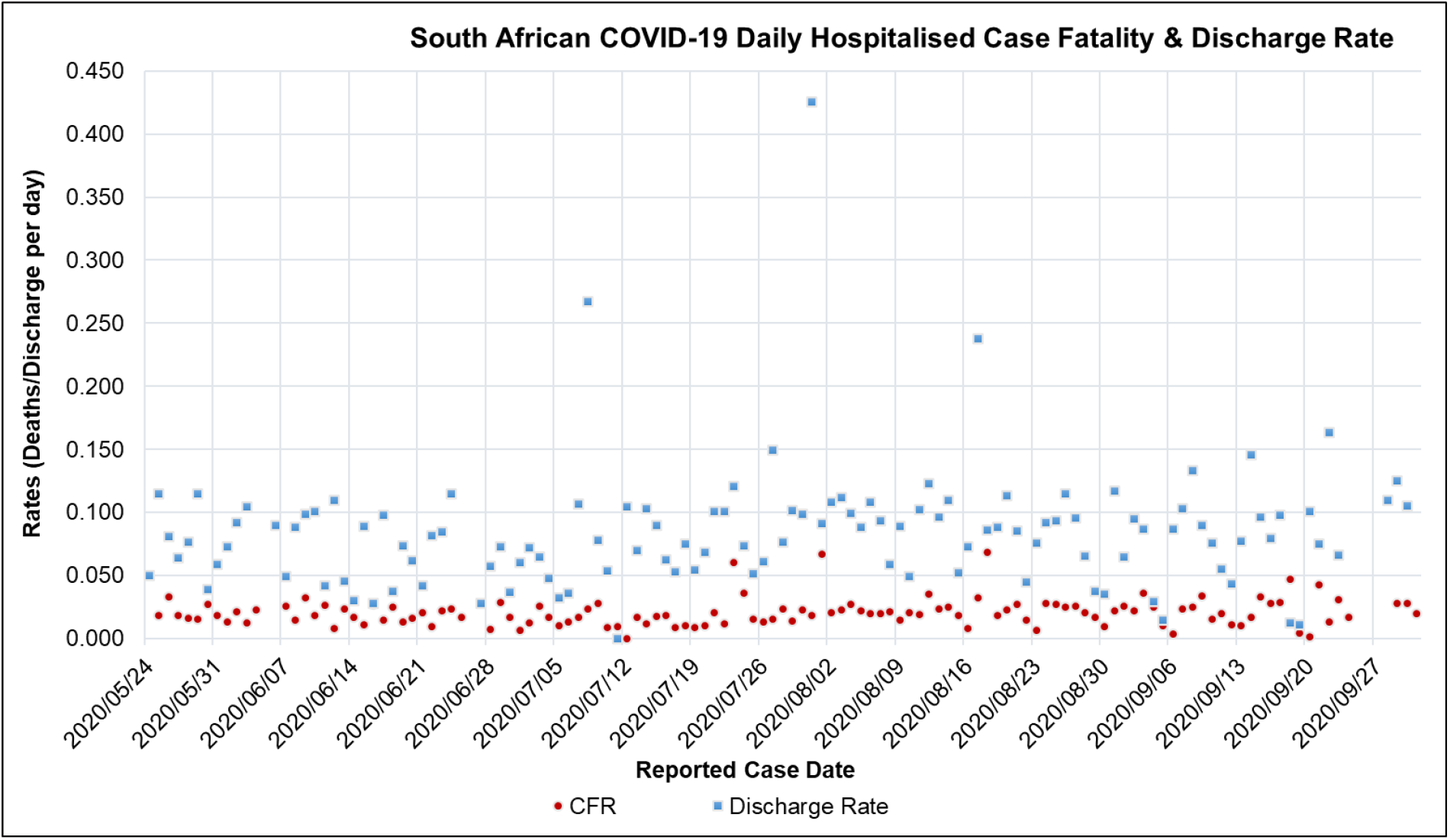
South African COVID-19 Hospitalised Case Fatality and Discharge Rate for the period of 2020/05/24 to 2020/10/01 [41]

**Figure A. 5:**
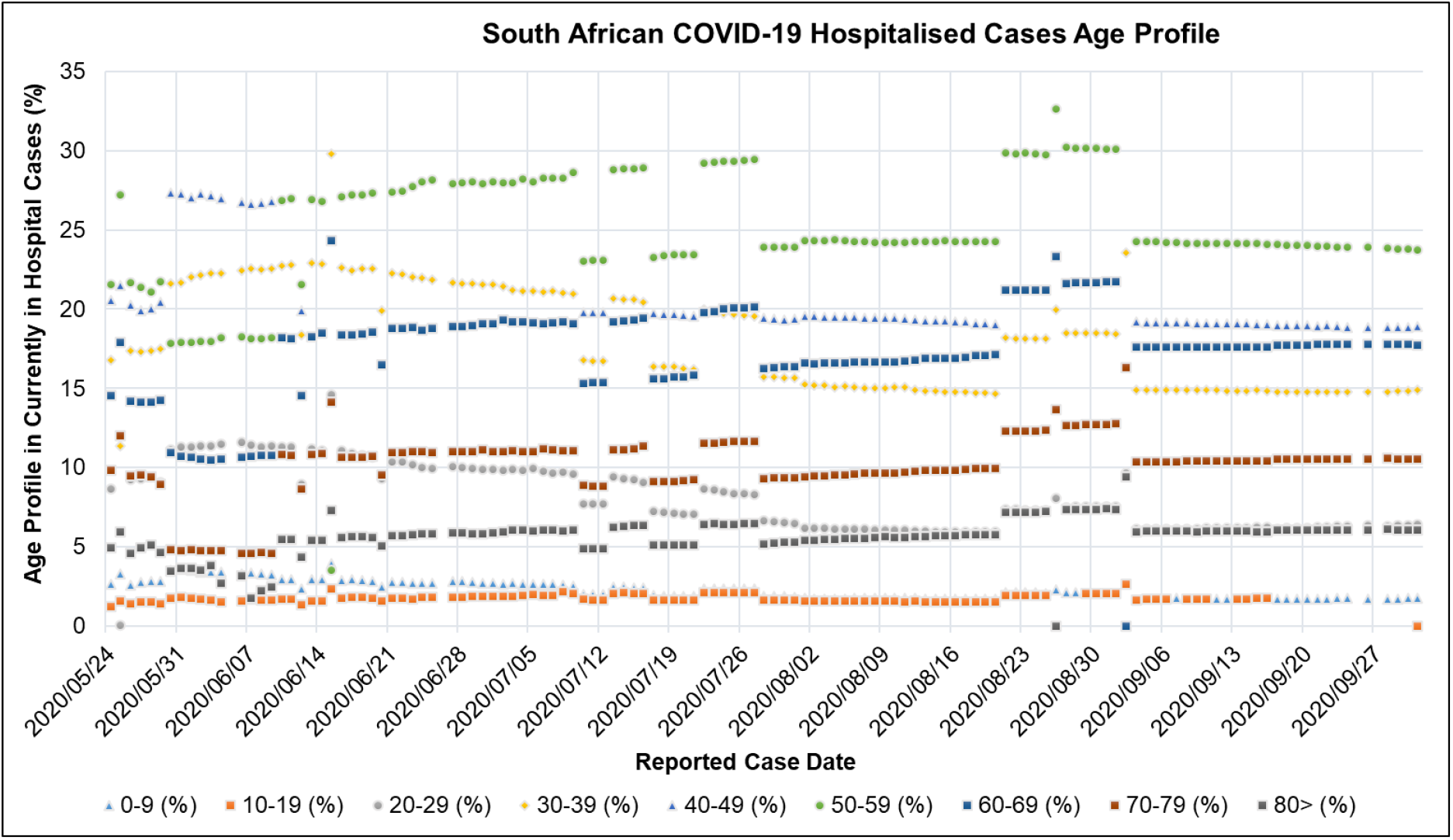
South Africa Hospitalised Case Age profile (%) for the periods of 2020/05/24 to 2020/10/01 [41]

**Figure A. 6:**
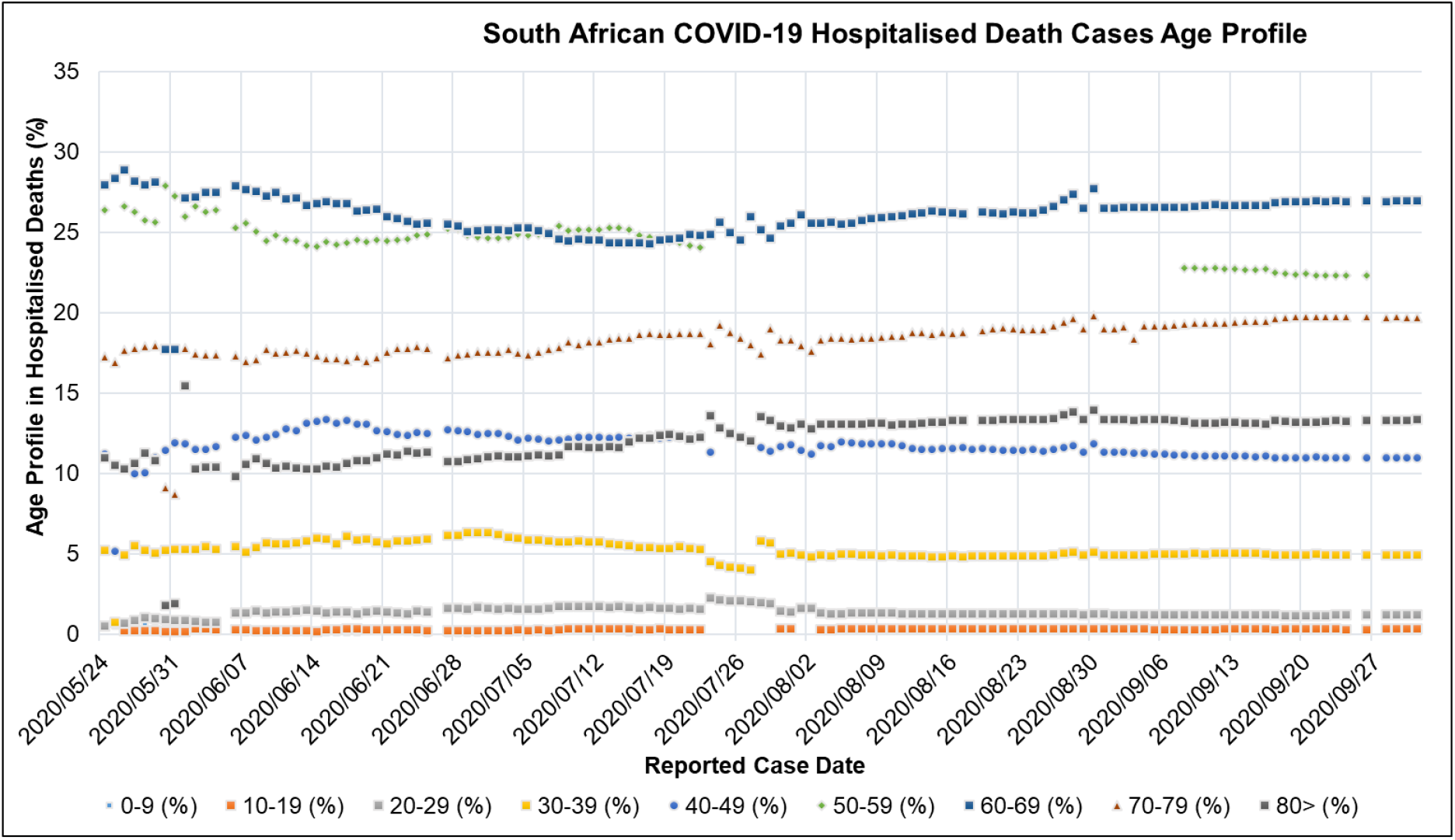
South Africa Hospitalised Deaths Age profile (%) for the periods of 2020/05/24 to 2020/10/01 [41]

**Figure A. 7:**
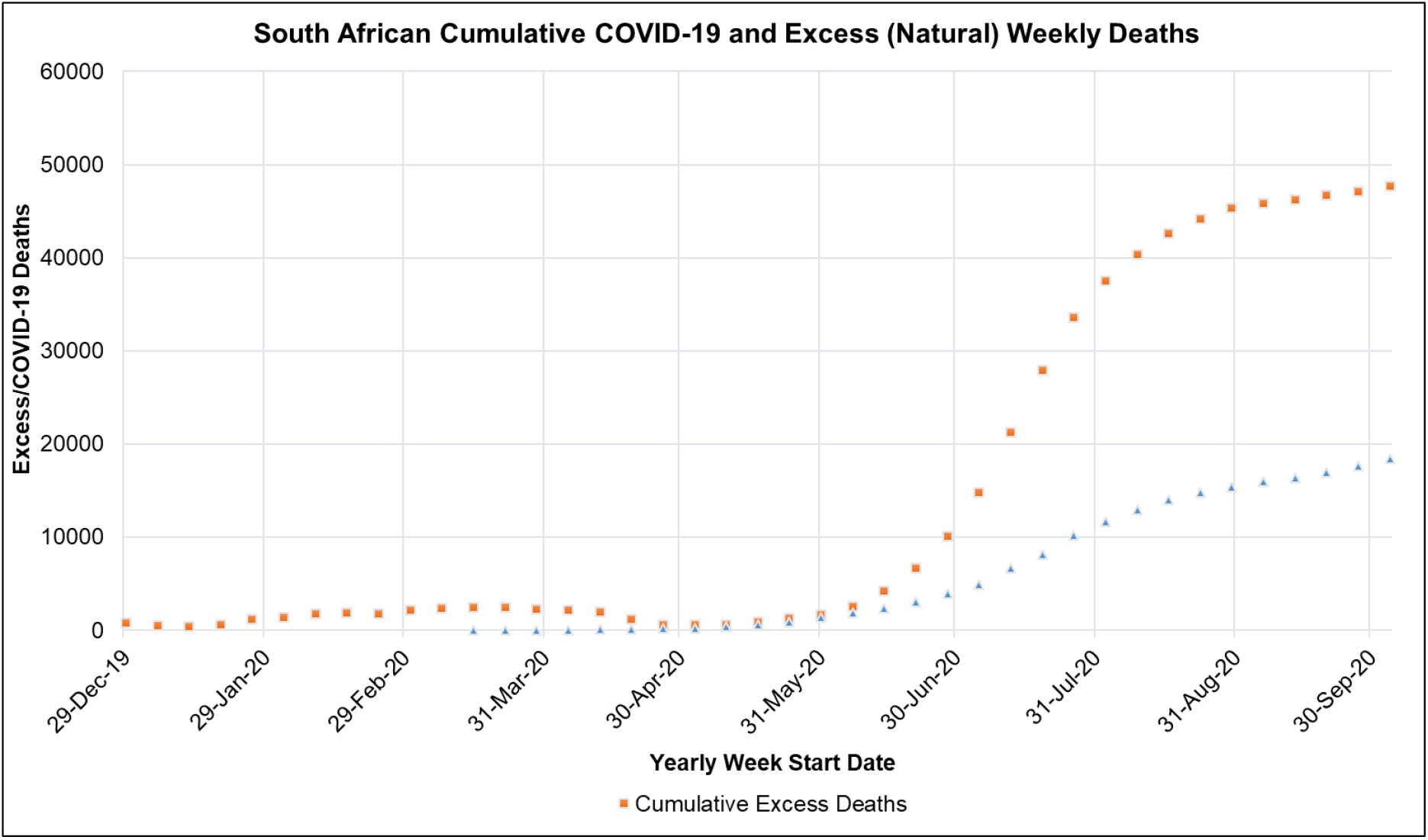
South African Cumulative Excess (Natural) Deaths and COVID-19 Reported Deaths for the period of 2019/12/29-2020/10/01 [44,45]

**Figure A. 8:**
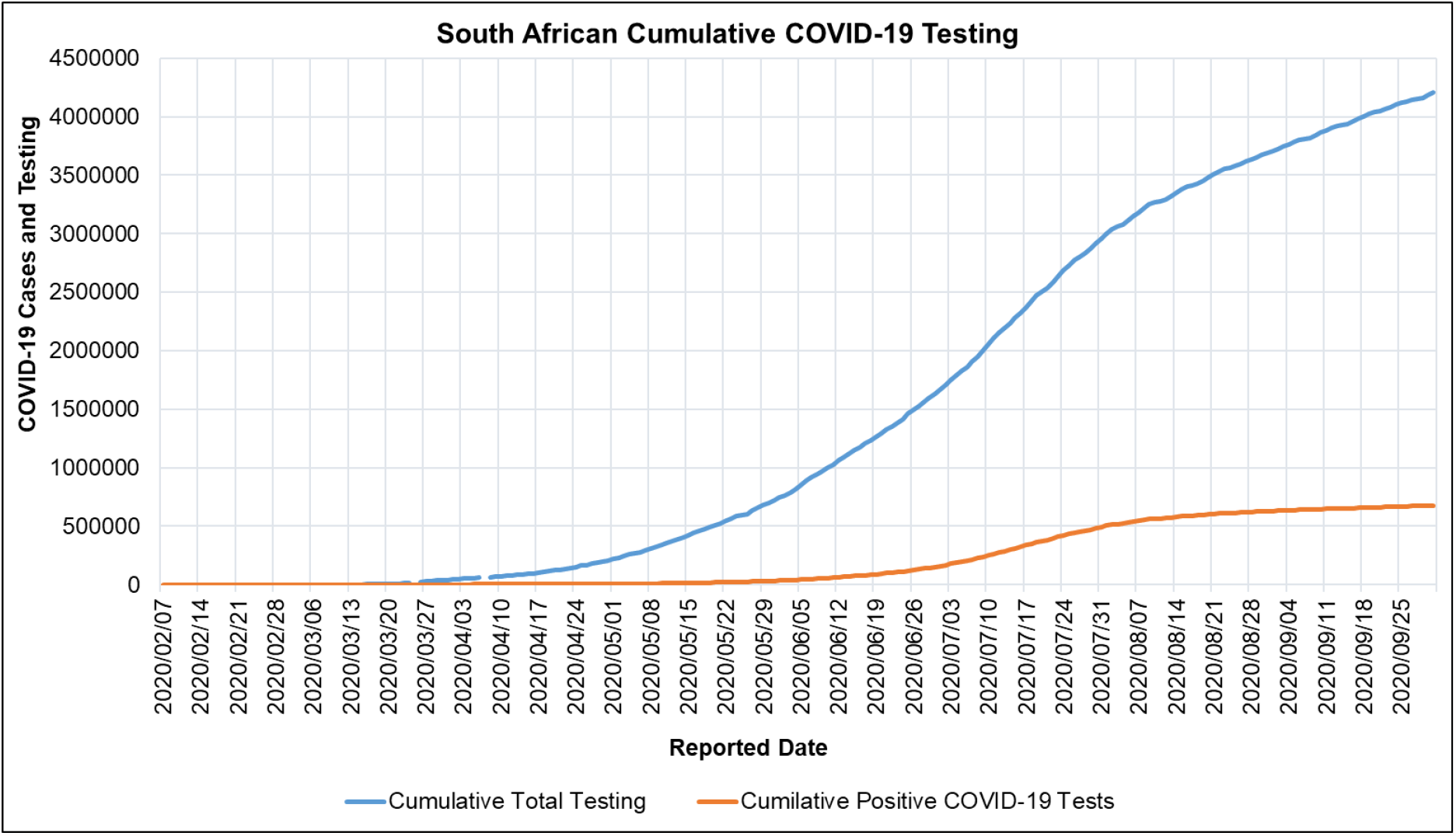
South African COVID-19 Cumulative Testing and Cases for the period 2020/02/07 to 2020/10/01 [80]

**Figure A. 9:**
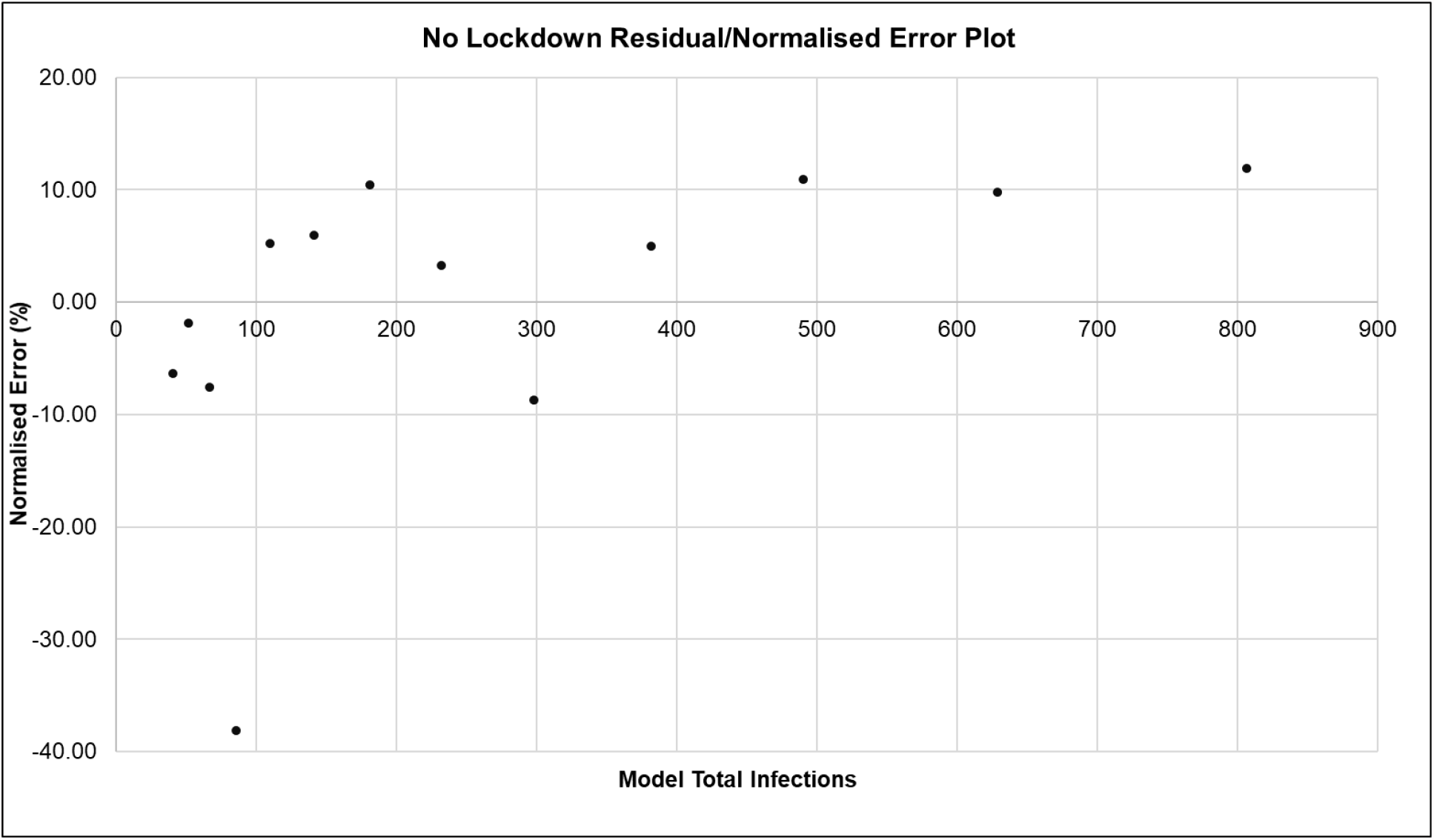
ARI COVID-19 SEIR Model Residual (Normalised Error) Plot between Model Data and Seeding Reported Case Data for the No Lockdown Scenario in the South African First Epidemic Wave.

**Figure A. 10:**
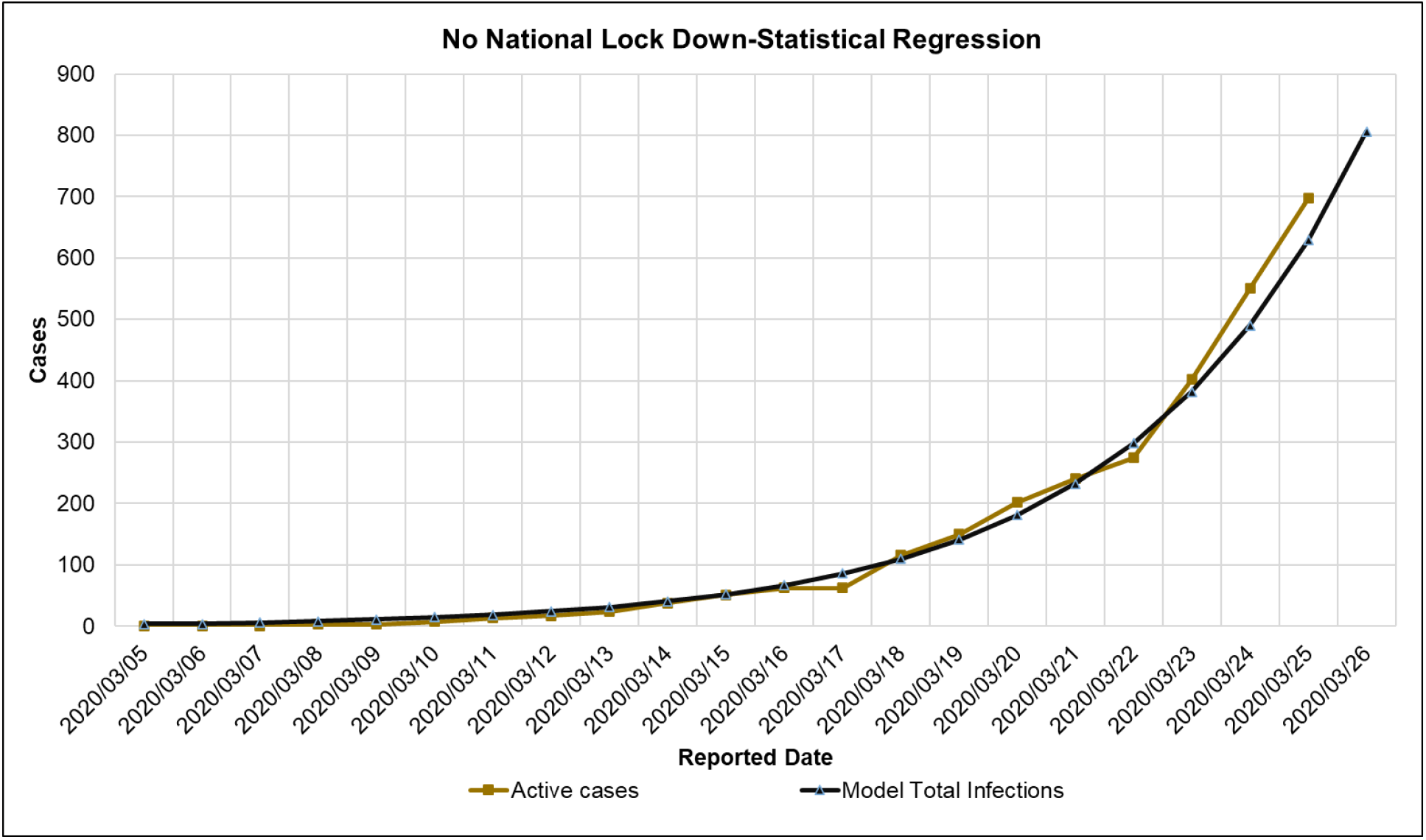
ARI COVID-19 SEIR Model Statistical Regression Plot between Model Data and Seeding Reported Case Data for the No Lockdown Scenario in the South African First Epidemic Wave.

**Figure A. 11:**
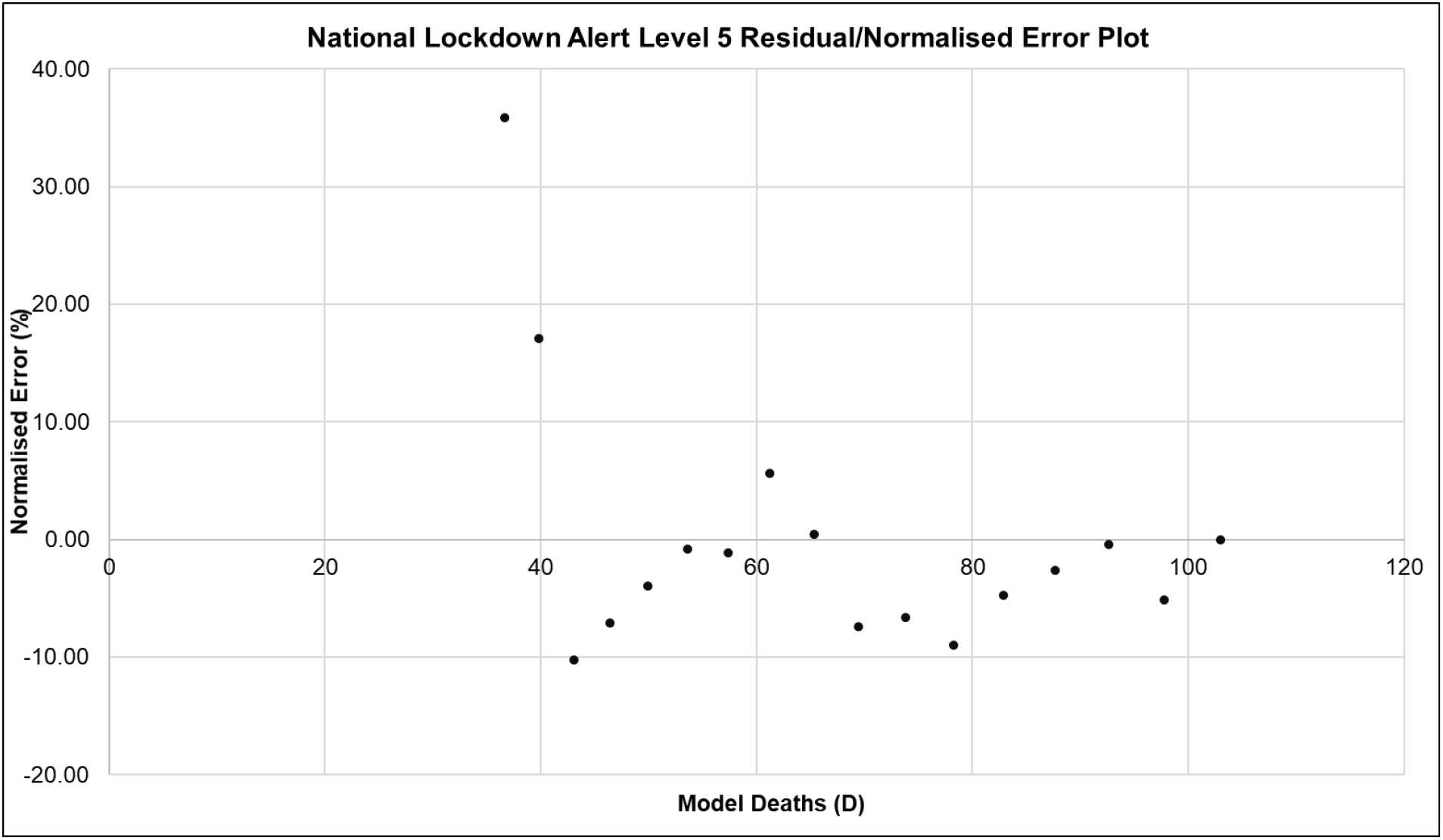
ARI COVID-19 SEIR Model Residual (Normalised Error) Plot between Model Data and Seeding Death Case Data for the National Alert Level 5 Scenario in the South African First Epidemic Wave.

**Figure A. 12:**
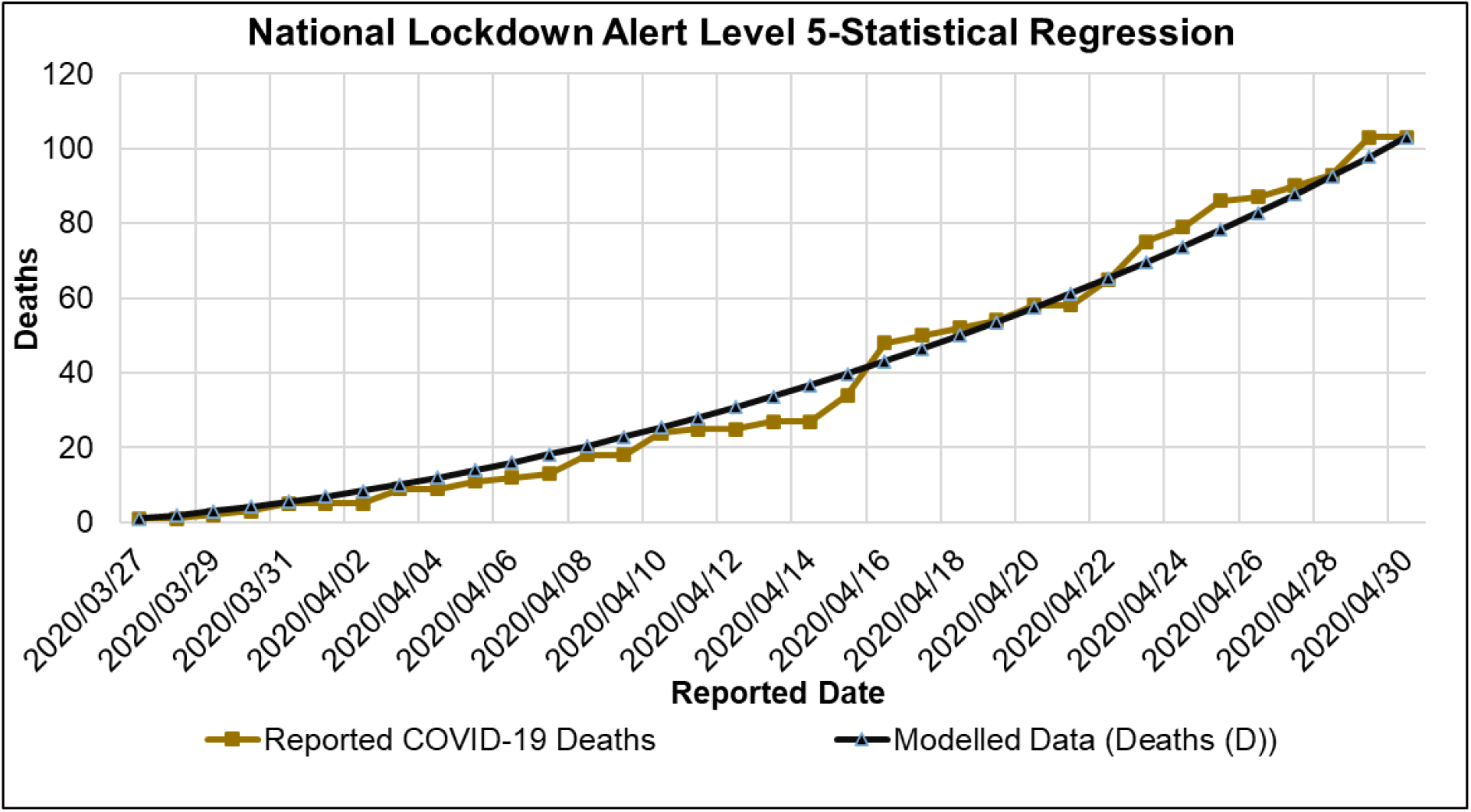
ARI COVID-19 SEIR Model Statistical Regression Plot between Model Data and Seeding Death Case Data for the National Alert Level 5 Scenario in the South African First Epidemic Wave.

**Figure A. 13:**
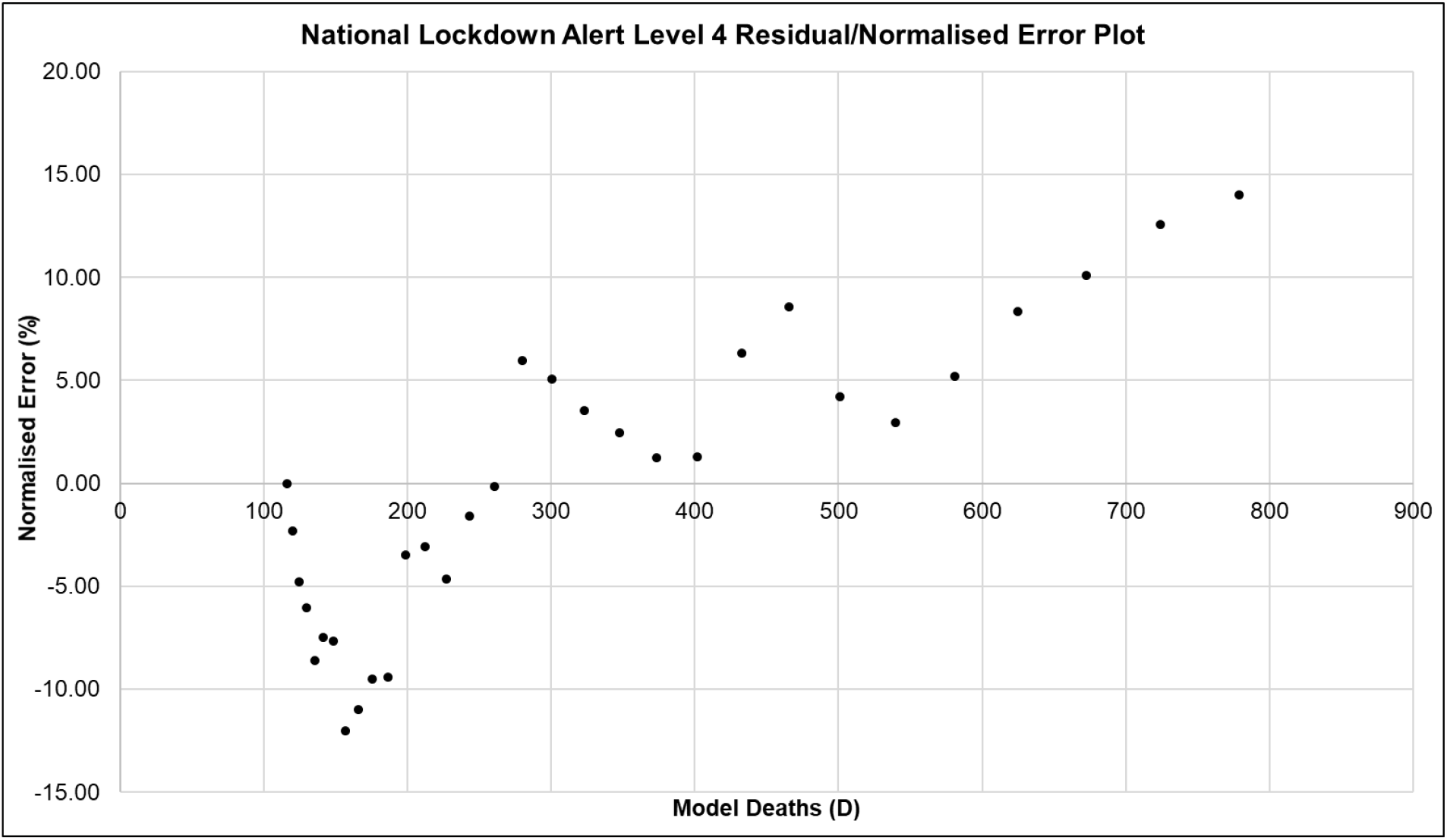
ARI COVID-19 SEIR Model Residual (Normalised Error) Plot between Model Data and Seeding Death Case Data for the National Alert Level 4 Scenario in the South African First Epidemic Wave.

**Figure A. 14:**
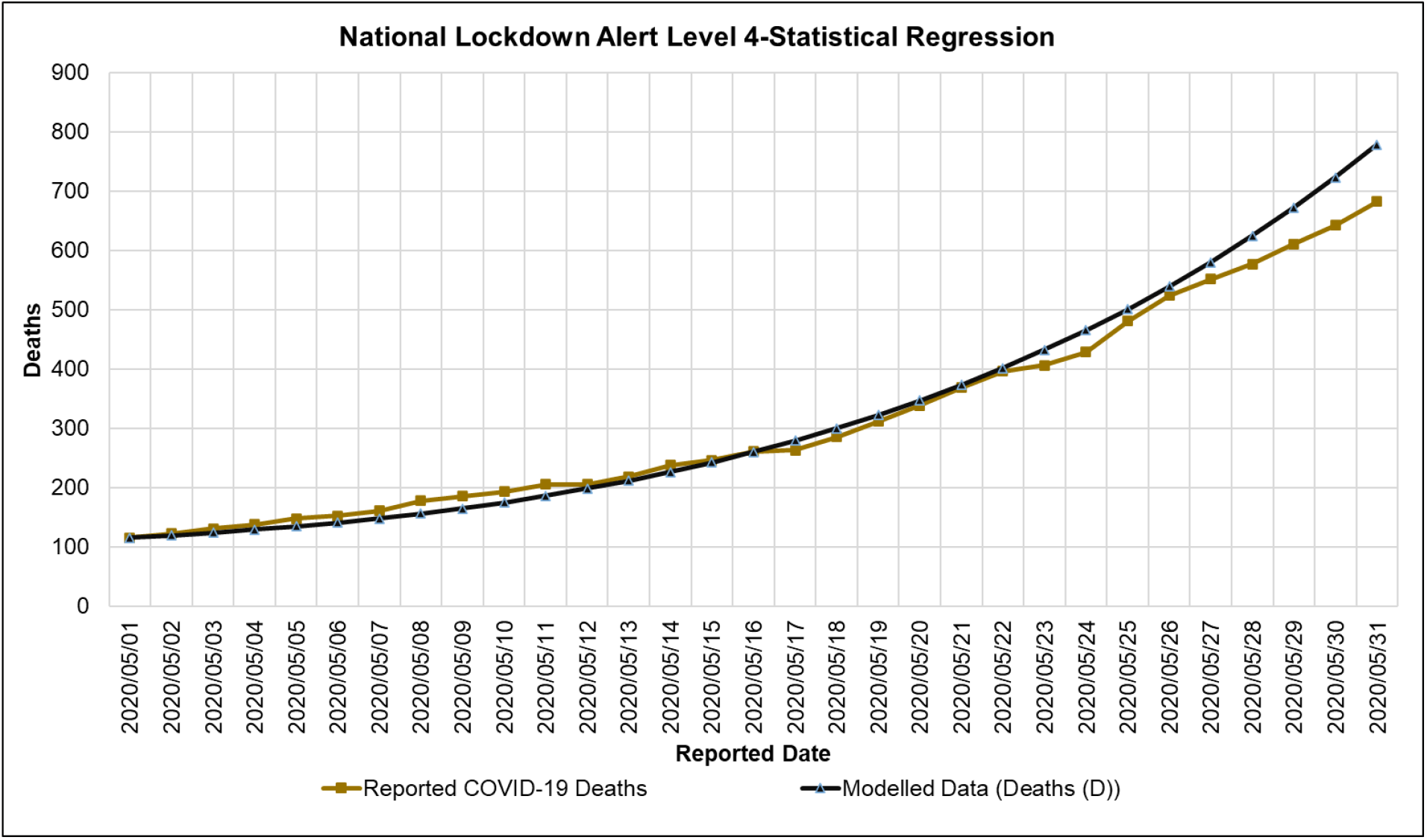
ARI COVID-19 SEIR Model Statistical Regression Plot between Model Data and Seeding Death Case Data for the National Alert Level 4 Scenario in the South African First Epidemic Wave.

**Figure A. 15:**
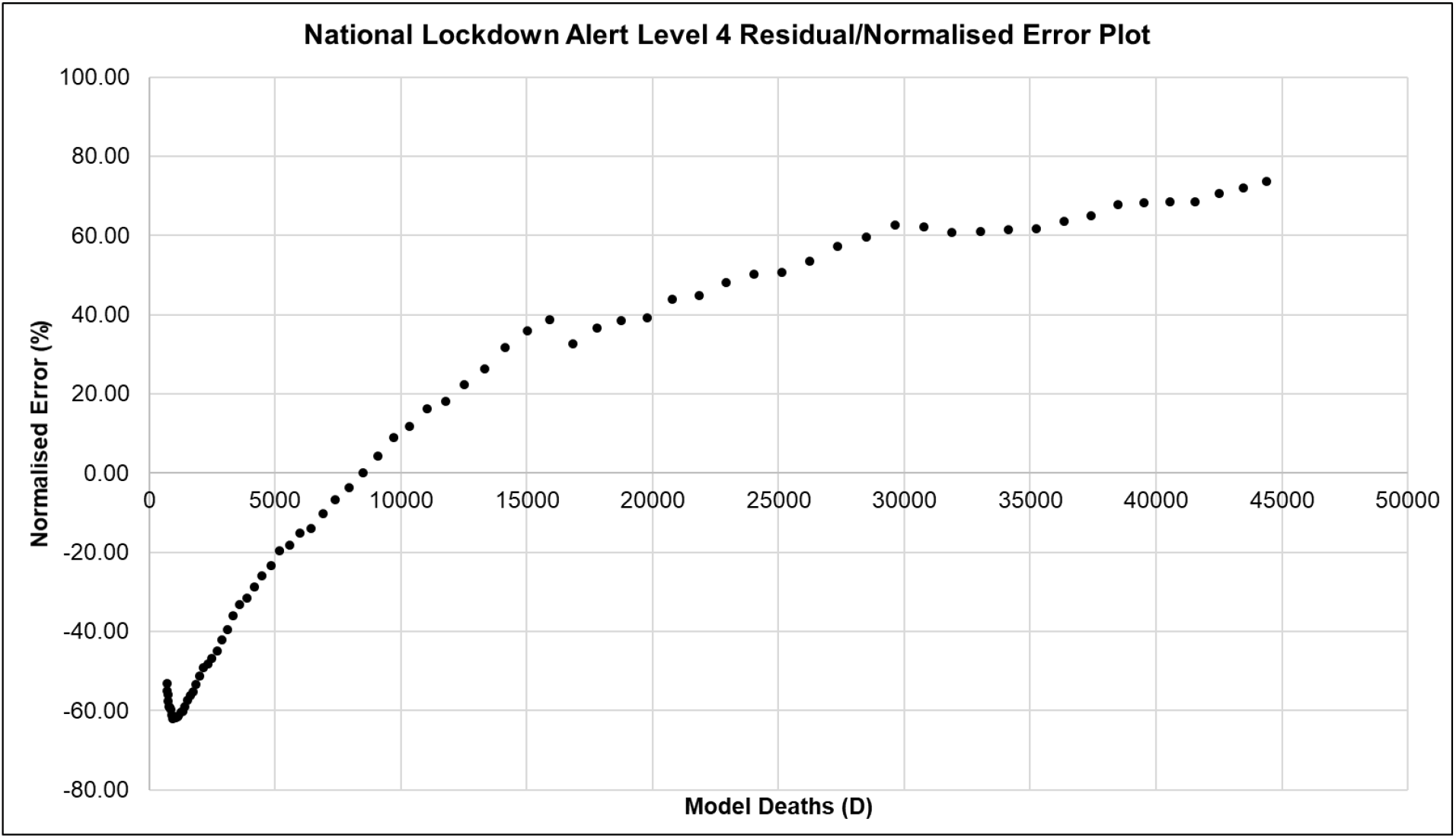
ARI COVID-19 SEIR Model Residual (Normalised Error) Plot between Model Data and Seeding Death Case Data for the National Alert Level 3 Scenario in the South African First Epidemic Wave.

**Figure A. 16:**
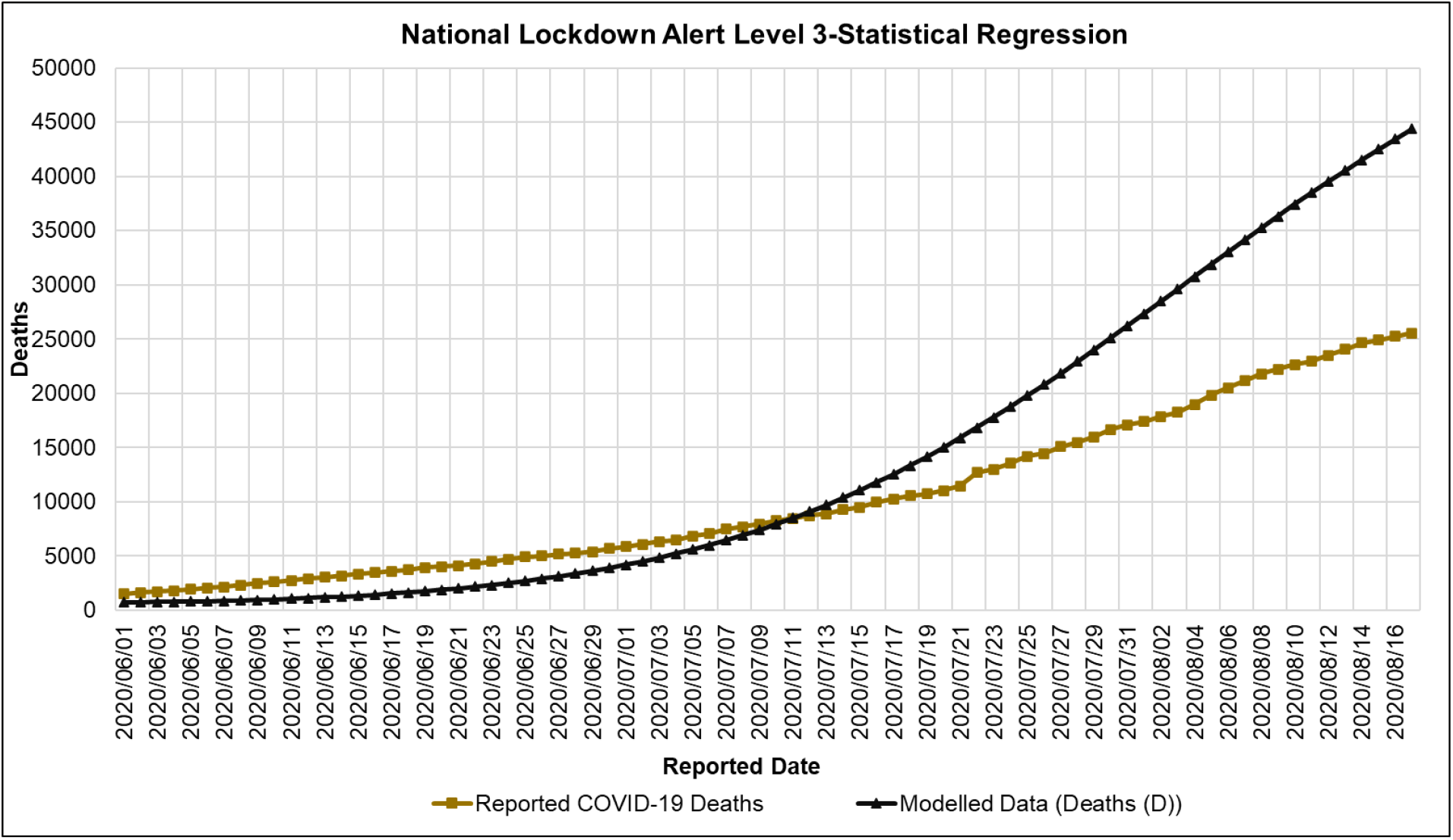
ARI COVID-19 SEIR Model Statistical Regression Plot between Model Data and Seeding Death Case Data for the National Alert Level 3 Scenario in the South African First Epidemic Wave.

**Figure A. 17:**
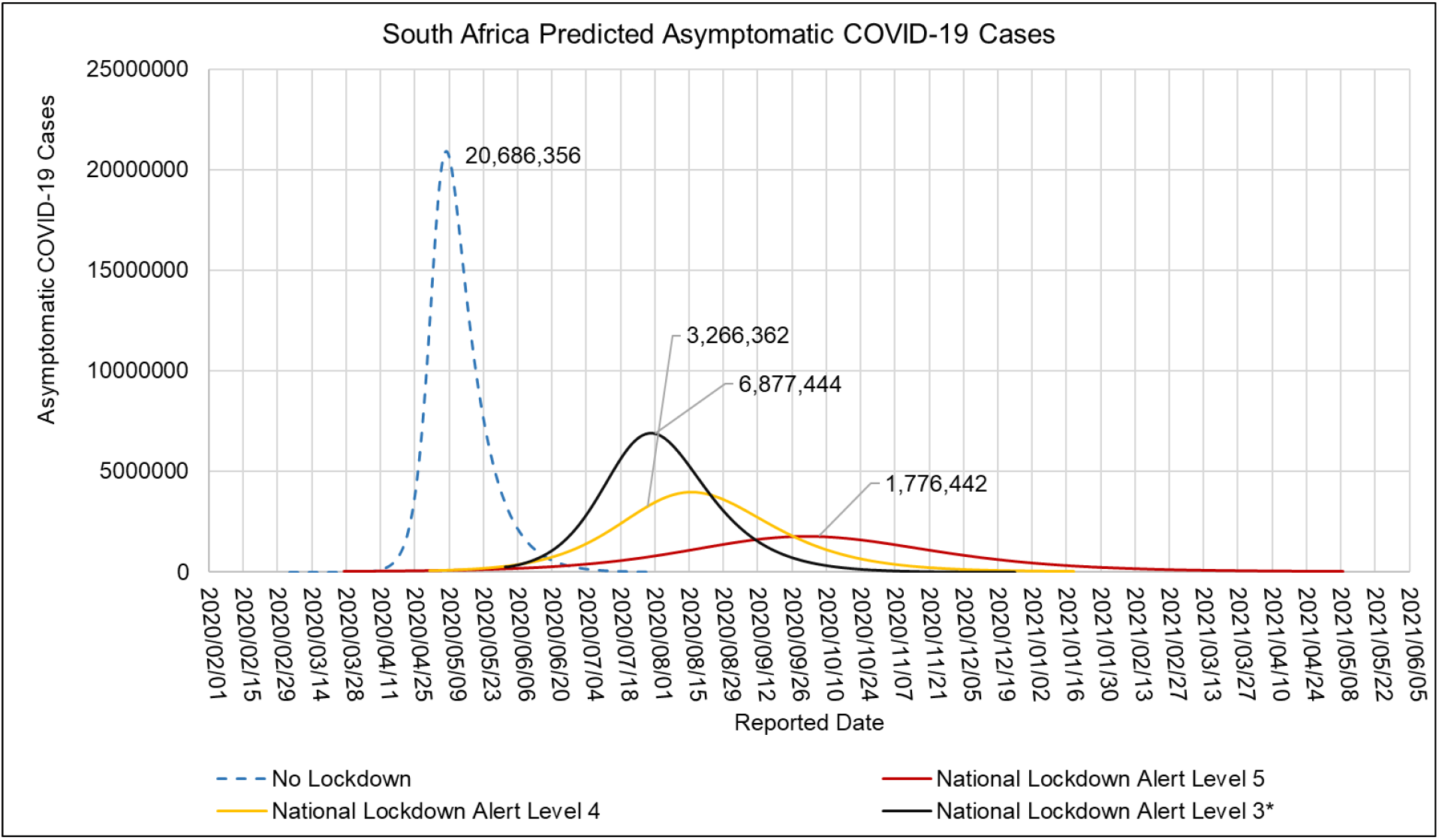
ARI COVID-19 SEIR Model Asymptomatic COVID-19 Cases in the South African First Epidemic Wave for the No Lockdown, Hard Lock down (National Lockdown Alert Level 5), Moderate Lock down (National Lockdown Alert Level 4) and Soft Lock down (National Lockdown Alert Level 3) scenarios

**Figure A. 18:**
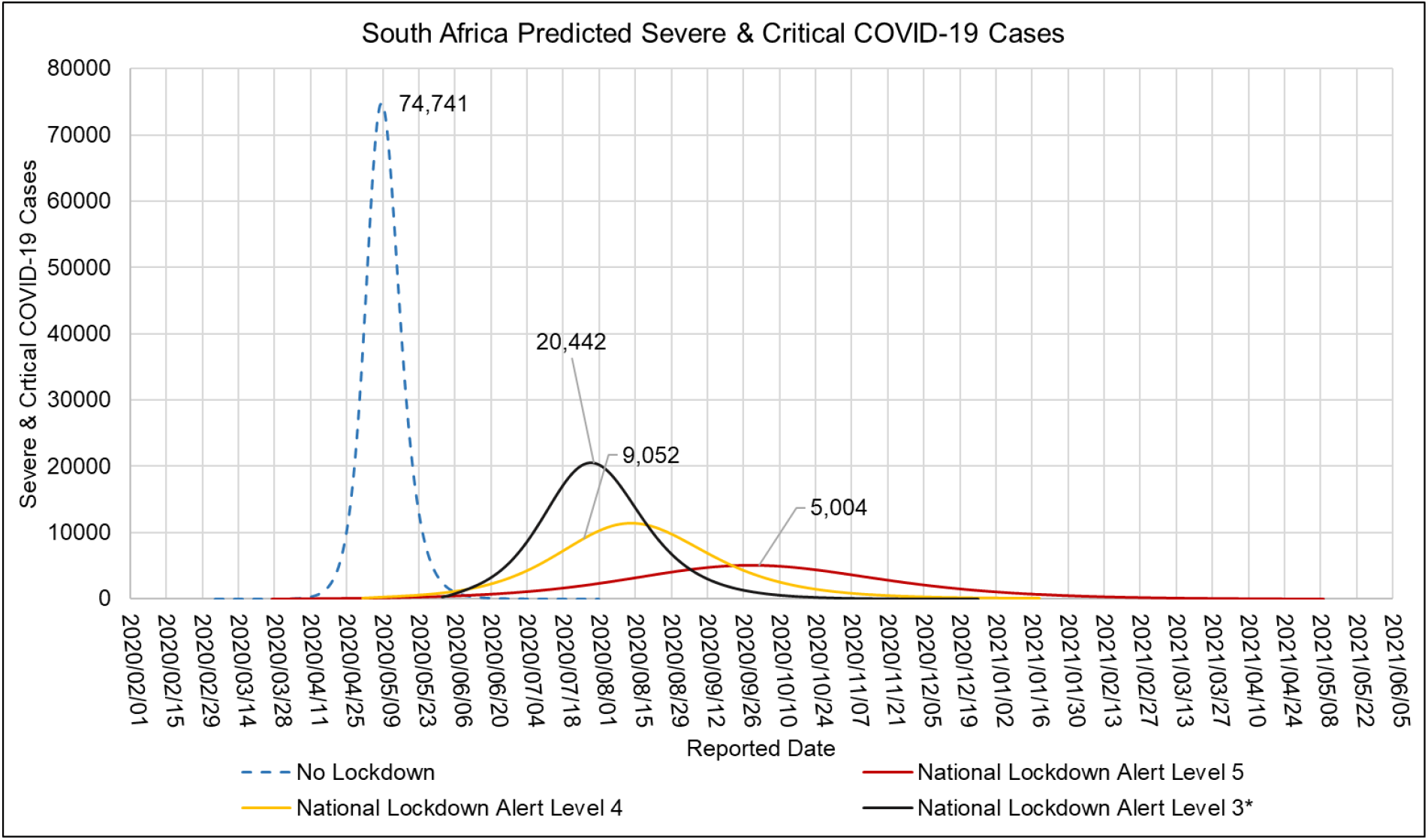
ARI COVID-19 SEIR Model Severe & Critical COVID-19 Cases in the South African First Epidemic Wave for the No Lockdown, Hard Lock down (National Lockdown Alert Level 5), Moderate Lock down (National Lockdown Alert Level 4) and Soft Lock down (National Lockdown Alert Level 3) scenarios

**Figure A. 19:**
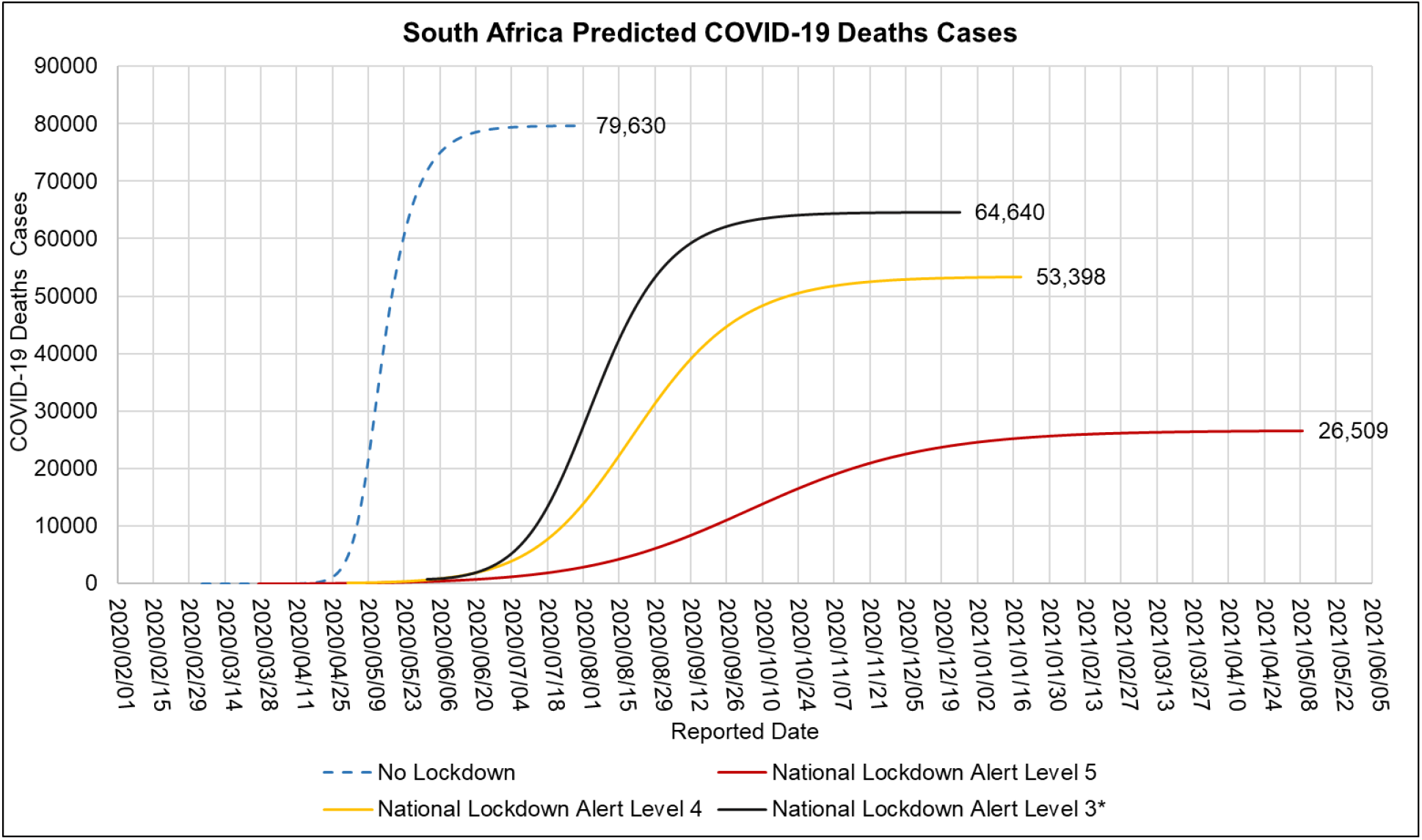
ARI COVID-19 SEIR Model COVID-19 Deaths in the South African First Epidemic Wave for the No Lockdown, Hard Lock down (National Lockdown Alert Level 5), Moderate Lock down (National Lockdown Alert Level 4) and Soft Lock down (National Lockdown Alert Level 3) scenarios

